# Mapping the association between environmental pollutants and steatotic liver disease: a systematic review and meta-analysis

**DOI:** 10.1101/2025.07.28.25332296

**Authors:** Xincheng Li, Jiajing Li, Jiangrong Zhou, Ibrahim Ayada, Qiuwei Pan, Luc J.W. van der Laan, Pengfei Li

## Abstract

**Introduction:** Environmental pollution poses increasing threats to public health, particularly in metabolic disorders. Steatotic liver disease (SLD) is characterized by metabolic dysfunctions of the liver and affects over one-third of the global population. However, whether exposure to environmental pollutants would increase the risk of SLD remains poorly understood.

**Aim:** To evaluate the association between exposure to environmental pollutants and the SLD risk.

**Methods:** A systematic search of Medline, Embase, and Web of Science databases, along with manually reviewed reference lists, was conducted from inception until May 30, 2025. Observational studies reporting quantitative effect estimates of environmental pollutants and SLD risk in adults were included, following PRISMA and MOOSE guidelines. Random-effect models were applied to pool the data.

**Results:** A total of 34 studies were included. Environmental pollutants in this study were categorized as air pollutants, endocrine-disrupting chemicals (EDCs), and heavy metals. Significant associations with increased SLD risk were observed for particulate matter (PM2.5: OR = 1.23; PM10: OR = 1.07; PM1: OR = 1.45) and nitrogen dioxide (NO_2_: OR = 1.19) per 10 μg/m³ increase. Among EDCs, exposure to bisphenol A (BPA: OR = 1.42), MECPP (OR = 1.43), MEHHP (OR = 1.54), MEOHP (OR = 1.38), and PFOA (OR = 1.23) was correlated with elevated SLD risk. Exposure to heavy metals, including lead (Pb: OR = 1.61), cadmium (Cd: OR = 2.32), mercury (Hg: OR = 2.41), barium (Ba: OR = 1.16), arsenic (As: OR = 1.09), and cobalt (Co: OR = 1.18) was significantly associated with increased SLD risk.

**Conclusions:** Exposure to a wide range of environmental pollutants significantly increased SLD risk, underscoring the need for public health interventions to mitigate pollutant exposure and its contribution to SLD development.

## 1. Introduction

Environmental pollution has emerged as one of the most pressing challenges, posing significant threats to human and overall well-being on the globe. The World Health Organization (WHO) estimates that approximately nine million deaths annually are attributable to environmental pollution(Fuller et al., 2022). Environmental pollutants encompass substances that contaminate air, soil, or water, which originate from both natural sources and human activities such as industrial discharge and agricultural practices(Fuller et al., 2022; Lee et al., 2024). Alarmingly, rapid urbanization and industrialization in recent decades have substantially accelerated the release of diverse environmental pollutants, heightening concerns about the risk of pollutants to public health(Silva et al., 2024).

Among various types of pollution, air pollution is a major public health concern that is driven by the accumulation of particulate matter (PM), nitrogen dioxide (NO_2_), carbon monoxide (CO), and other gaseous pollutants in the atmosphere(Orellano et al., 2020). Apart from air pollutants, endocrine-disrupting chemicals (EDCs) and heavy metals represent two other important categories of environmental toxicants that adversely affect human health(Ismanto et al., 2022). These pollutants are capable of infiltrating the human body through inhalation, ingestion, or dermal contact, contributing to organ damage and immune system dysfunction(Chia et al., 2025). Notably, environmental pollutants are peculiarly associated with the development of chronic diseases such as metabolic disorders(Y. Zhang et al., 2024). Emerging evidence suggests that certain pollutants may interfere with hepatic lipid metabolism by inducing oxidative stress or inflammatory responses, both of which are potentially involved in the pathogenesis of steatotic liver disease (SLD)(Jiang et al., 2025; Xu et al., 2019). However, the relationship between environmental pollutant exposure and SLD remains largely underexplored.

Steatotic liver disease (SLD) refers to the condition where excessive lipids accumulate in the liver. SLD affects more than one-third of the global population and is mainly driven by obesity, type 2 diabetes, and alcohol intake(Israelsen et al., 2024). In the EU and USA, SLD has become the leading cause of liver cirrhosis, which results in millions of deaths each year(Huang et al., 2023). Since the liver plays a central role in taking up toxic substances and exerting metabolic functions, we hypothesize that environmental pollutants may contribute to hepatic injury and disease progression, thereby elevating the development or progression of SLD. Supporting this hypothesis, recent evidence showed that subacute exposure to PM2.5 elicited hepatocellular oxidative stress and inflammation, and aberrantly activated the PI3K/AKT signaling pathway, leading to hepatic insulin resistance and metabolic dysfunction(Lu et al., 2025). Similarly, a population-based longitudinal study has reported the association between air pollution exposure and elevated liver enzyme levels, suggesting that air pollutants may induce hepatic injury(Li et al., 2022). Moreover, a recent large cohort study found that all chronic liver diseases were caused by over 70% of the environmental risk factors (exposome) rather than age, sex, or genetic factors(Argentieri et al., 2025). Nevertheless, further studies are warranted to determine the specific relationship between different types of environmental pollutants and the risk of SLD.

In this systematic review and meta-analysis, we aim to comprehensively assess the association between exposure to environmental pollutants, including air pollutants, endocrine-disrupting chemicals, and heavy metals, and the risk of SLD.

## 2. Methods

### 2.1. Study Design

This study followed the Preferred Reporting Items for Systematic Reviews and Meta-Analyses (PRISMA) guidelines(Page et al., 2021) and the Meta-Analysis of Observational Studies in Epidemiology (MOOSE) reporting guideline(Stroup et al., 2000) and was registered in the International Prospective Register of Systematic Reviews(PROSPERO). The registration number is CRD42024608282 (www.crd.york.ac.uk/PROSPERO).

### 2.2. Data sources and searches

We conducted a systematic search across three databases, including Medline, Embase, and Web of Science, and manually reviewed reference lists for studies on the association of environmental exposure with SLD. These databases were searched with restrictions to only English language and studies from inception until May 30, 2025, using search terms relating to metabolic liver disease and environmental exposure. The search was performed in collaboration with the medical library of Erasmus University Rotterdam and the full search strategies are provided in eAppendix 1 in the Supplement.

### 2.3. Study Selection and Data Extraction

To identify relevant articles, the titles and abstracts of the retrieved papers were exported to EndNote version X9 (Clarivate), where duplicates were removed. Then, three reviewers (X.L., J.L., and J.Z.) worked independently to determine whether a study met the inclusion criteria (eAppendix 2 in the Supplement). Reviewers first screened titles and abstracts, and subsequently the full text of potentially eligible articles. The following data were extracted from eligible studies: the first author, publication year, study design, geographic location, age, sample size and sample type, exposures, exposure assessment methods, SLD diagnostic criteria, and adjusted covariates. Among included studies, distinct measuring methods were used for different categories of environmental pollutants, and effect estimates were reported either as continuous variables or categorical comparisons. For studies presenting results as continuous variables, corresponding effect estimates were collected as continuous measures in our meta-analysis. In contrast, for studies reporting results as categorical comparisons, effect estimates were extracted based on the lowest and highest gradients (e.g., lowest and highest tertile or quartile). To reduce potential variability, effect estimates derived from the same type of comparison were prioritized wherever possible. If multiple models with varying levels of covariate adjustment were reported, estimates from the model with the most comprehensive adjustment were selected to minimize the influence of confounding factors.

### 2.4. Study Quality Assessment, Risk of Bias, and Evidence Level Assessment

In this study, the Newcastle–Ottawa Scale (NOS) was employed to assess the quality of case-control and cohort studies(Stang, 2010), while the Joanna Briggs Institute (JBI) checklist was utilized for evaluating the quality of cross-sectional studies(Aromataris E, 2024) (eTable 1-3 in the Supplement). In addition, the potential risk of bias in included literature was assessed according to the OHAT Risk of Bias Rating Tool(“National Toxicology Program. Protocol for systematic review of effects of fluoride exposure on neurodevelopment. US Dept of Health & Human Services, Public Health Service, National Institutes of Health;,”) (eTable 4 in the Supplement). Confidence in the body of evidence and related level of evidence was evaluated using the NTP/OHAT framework(Rooney et al., 2014), based on the GRADE approach(Morgan et al., 2016; Schünemann et al., 2011) (eTable 5 in the Supplement).

### 2.5. Statistical Analysis

All statistical analyses were performed in R version 4.3.1 using the “meta” package to ensure a rigorous and transparent data synthesis process. Odds ratios (ORs) were pooled across studies using the inverse-variance method to calculate the overall effect estimates. To assess heterogeneity among the included studies, τ² and the P value were used as qualitative indicators, where a larger τ² and a smaller P value suggest greater heterogeneity. In addition, I² was used to quantitatively evaluate heterogeneity across studies. Heterogeneity was considered substantial when I² ≥ 50%, and a random-effects model was applied to pool the effect sizes accordingly. Meta-analysis was conducted only if at least two studies were available for each pollutant. The association between environmental pollutants and SLD was estimated using pooled ORs and 95% confidence intervals (CIs). Subgroup analyses were performed to explore potential sources of heterogeneity based on study characteristics or methodological differences. Sensitivity analyses were carried out using the leave-one-out method, where each study was sequentially excluded to evaluate its impact on the overall results, ensuring the robustness of the findings. Publication bias was assessed using Egger’s test, funnel plots, and the trim-and-fill method to provide a comprehensive evaluation.

## 3. Results

### 3.1. Study selection and characteristics

A total of 2,119 records were identified after deduplication through systematic searches of literature databases from Medline (839), Embase (564), and Web of Science (708), with 8 additional records retrieved manually. Of these, 34 studies met the inclusion criteria for analyzing the association between environmental pollutants and SLD (Figure 1). These studies comprised 31 cross-sectional studies, 2 case-control studies, and 1 cohort study, encompassing a total of 48,887,089 adult participants with publication years spanning from 2019 to 2025 (Table 1).

**Table 1.**
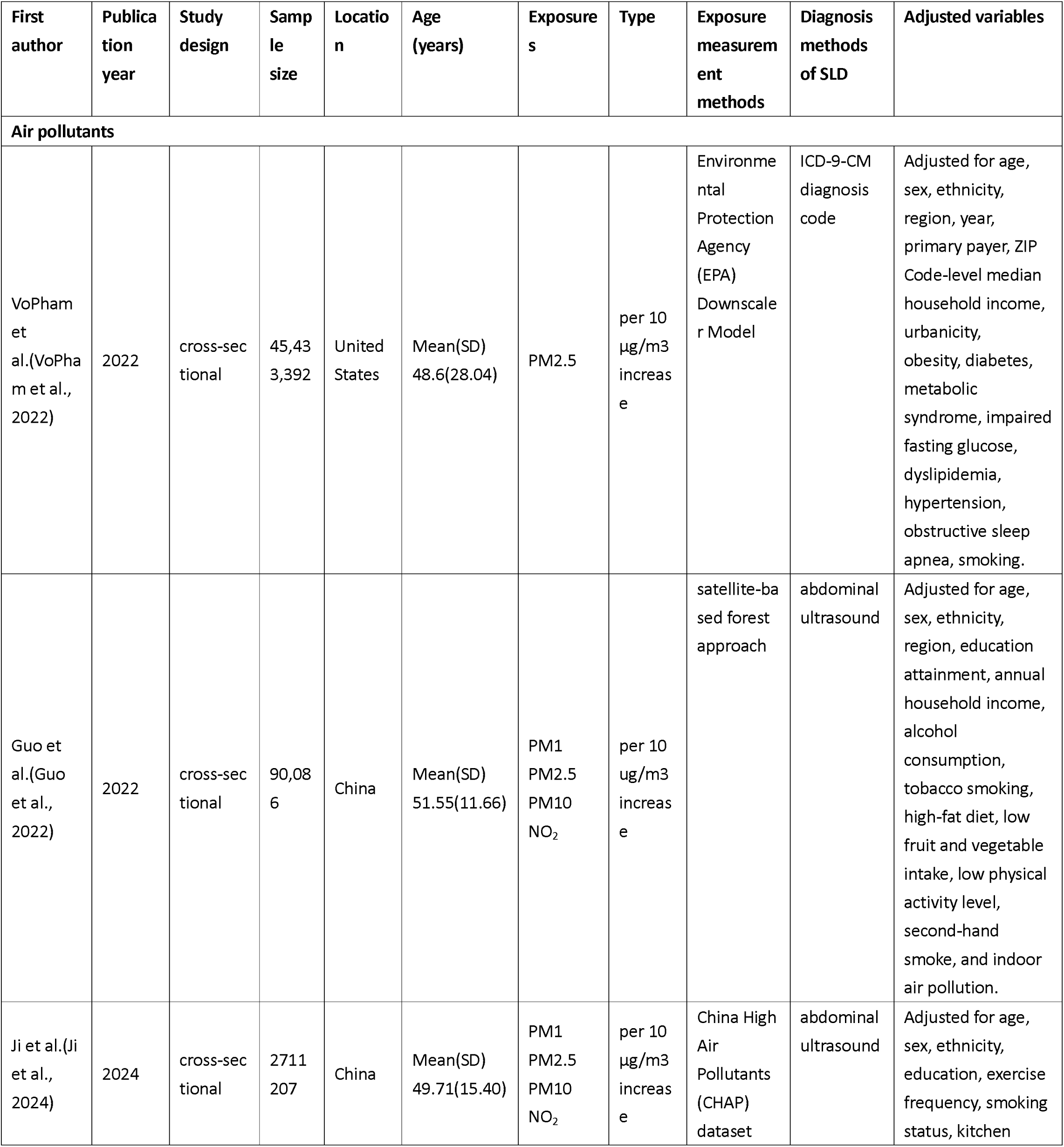
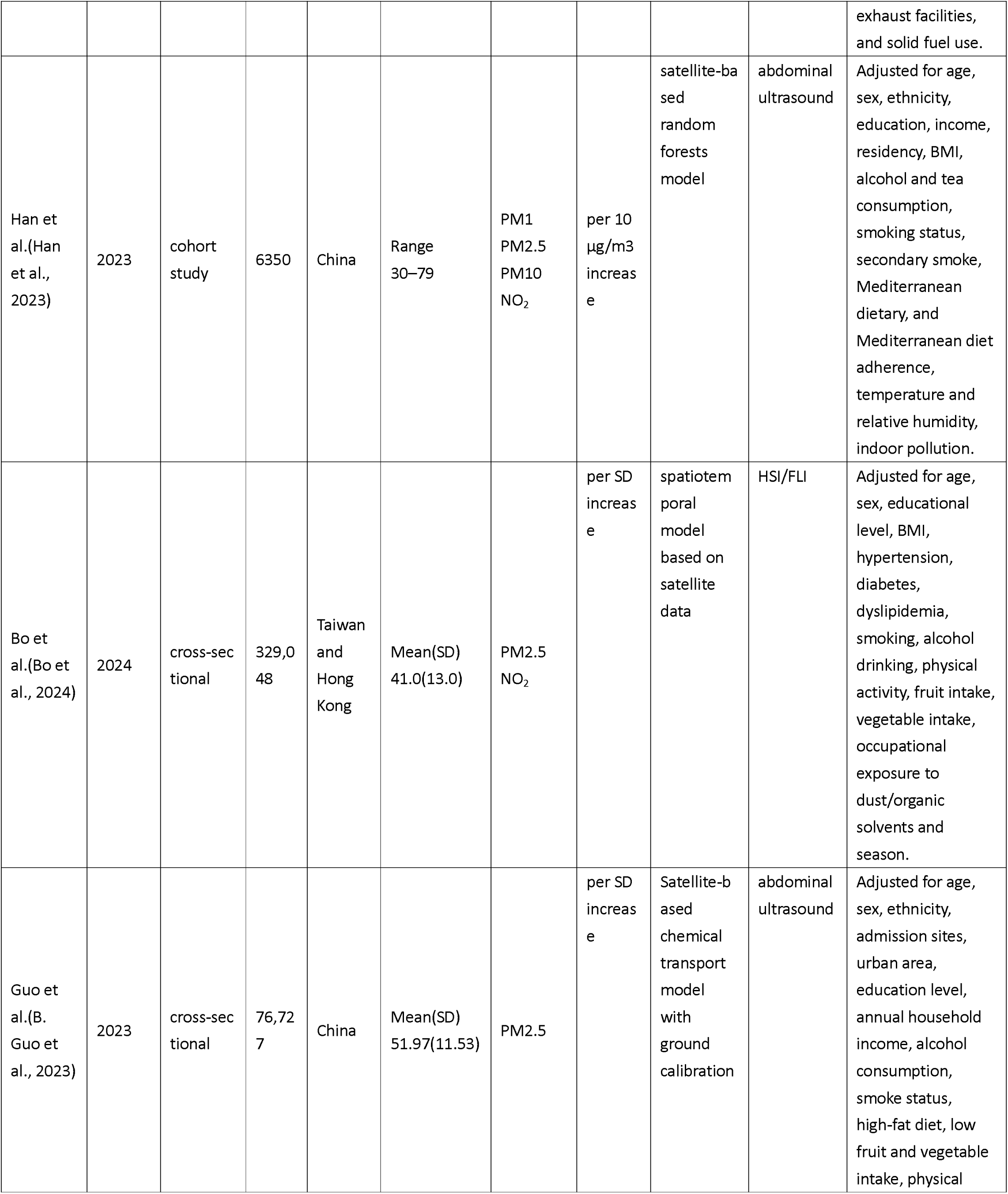
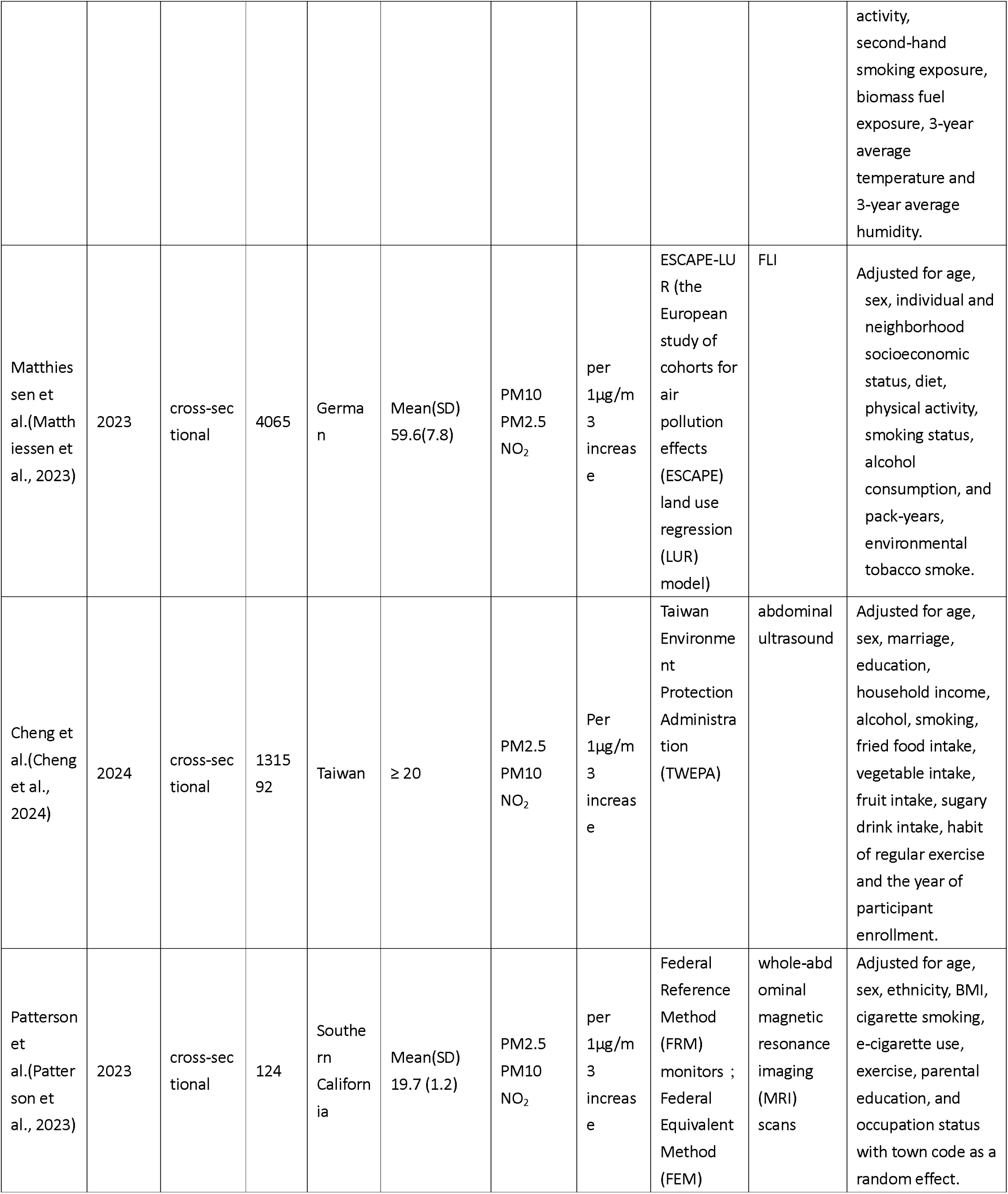
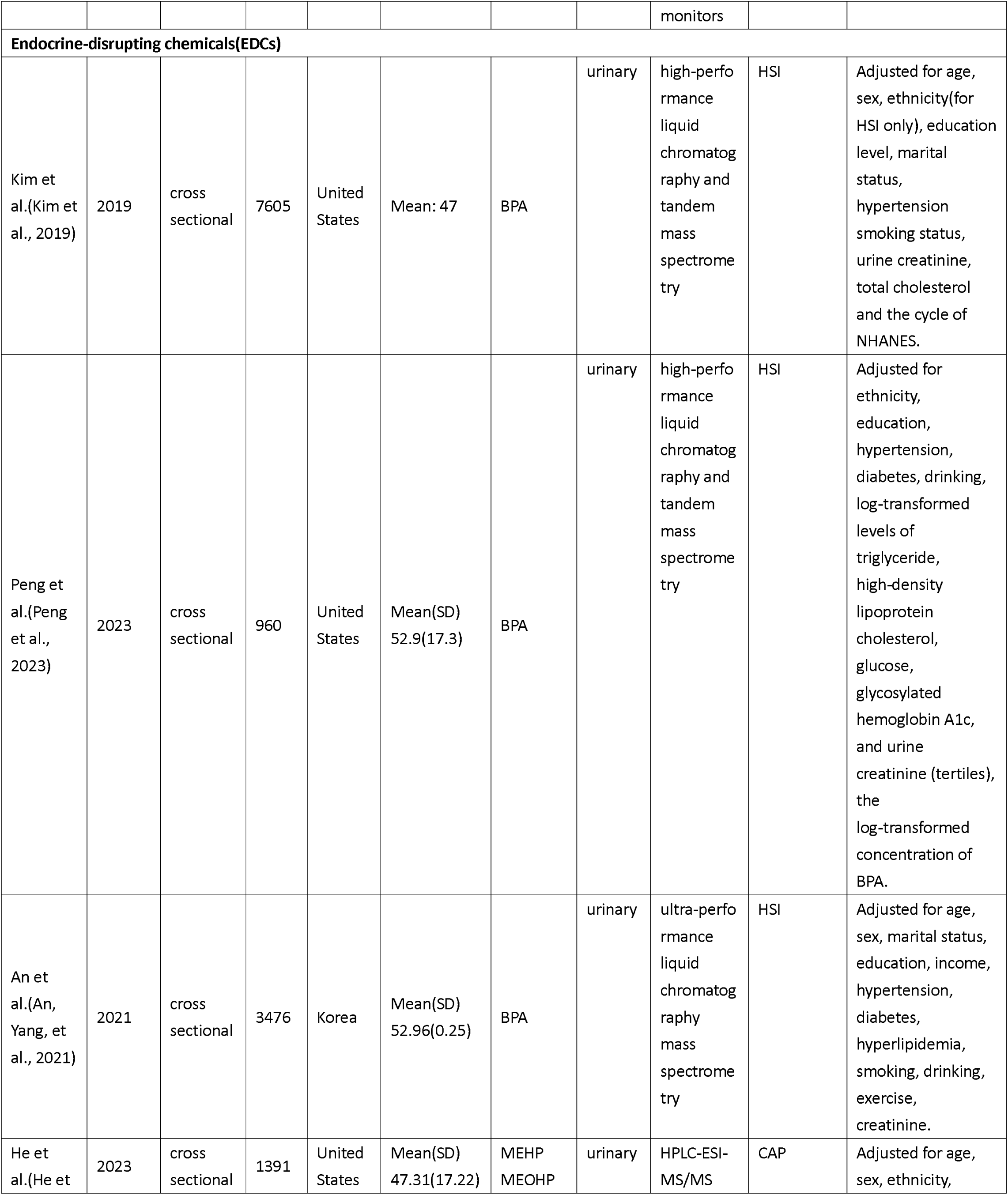
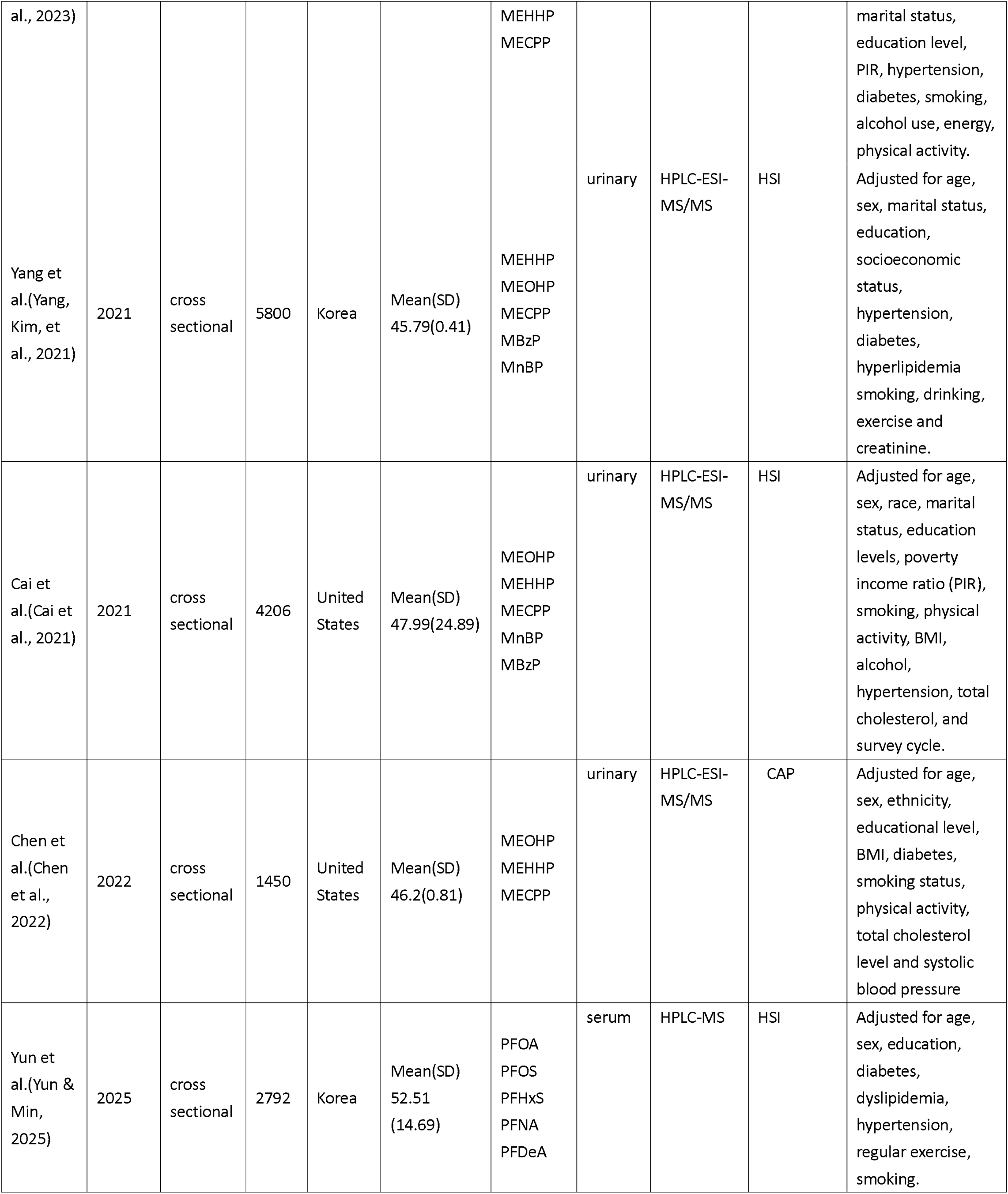
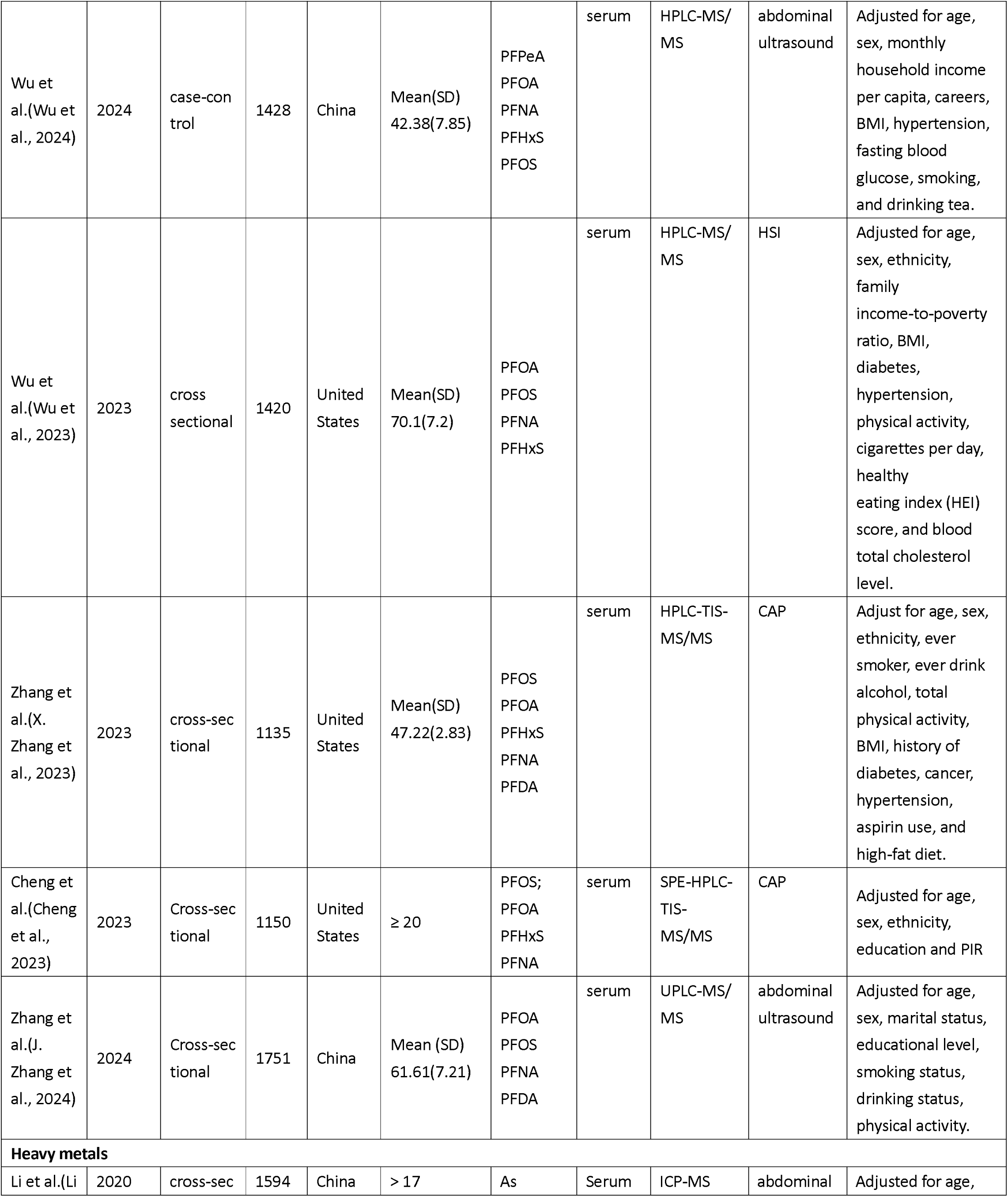
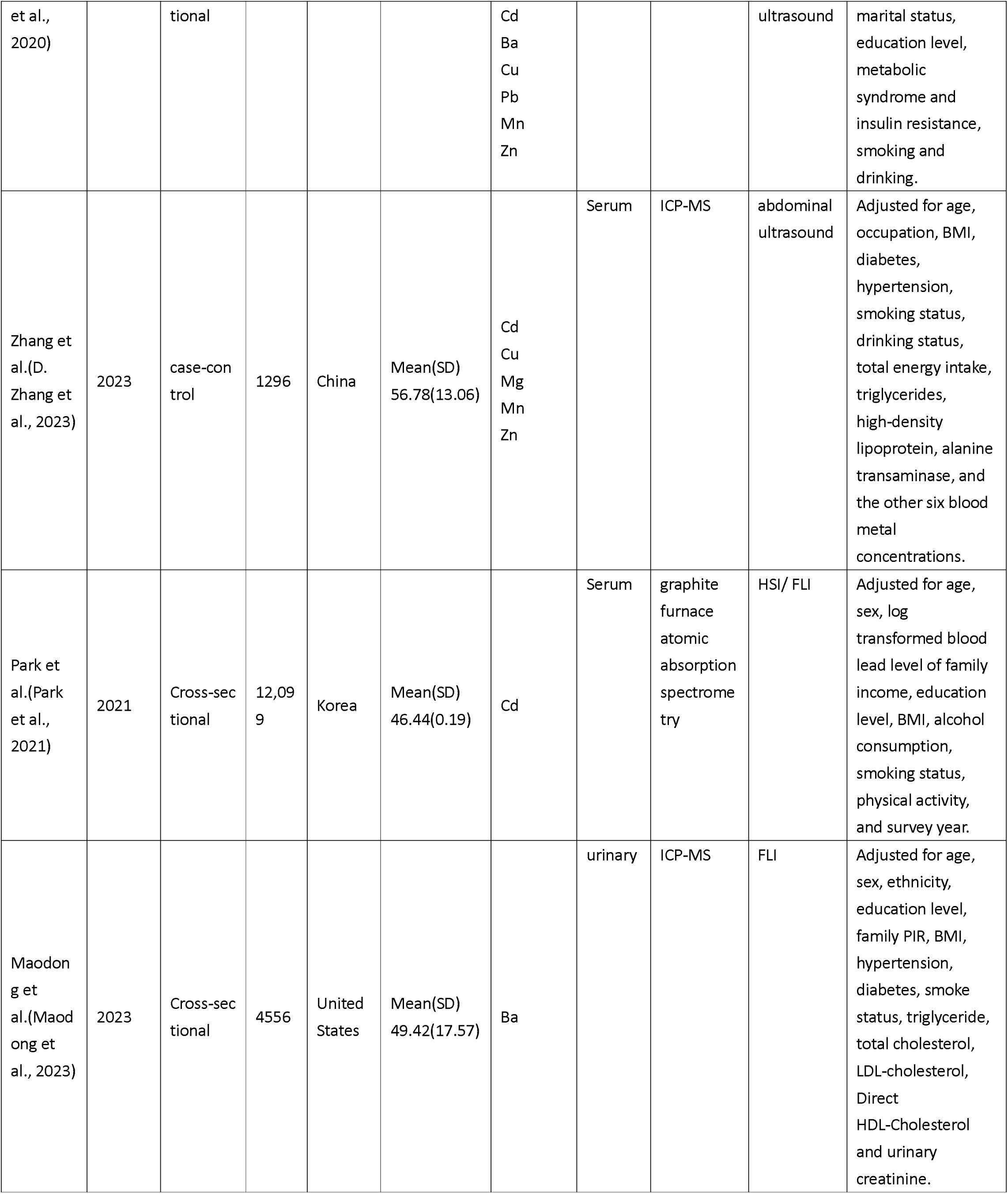
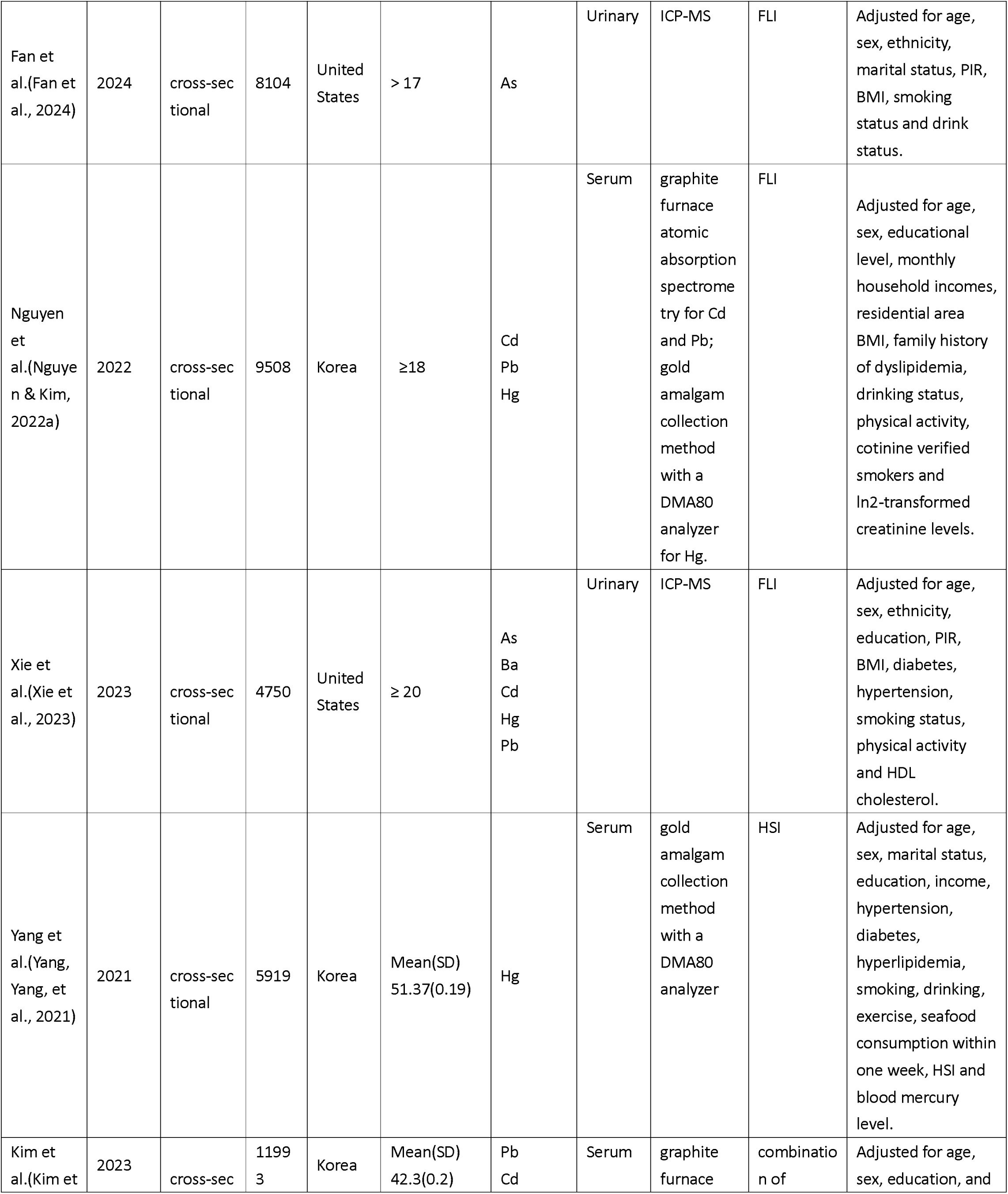
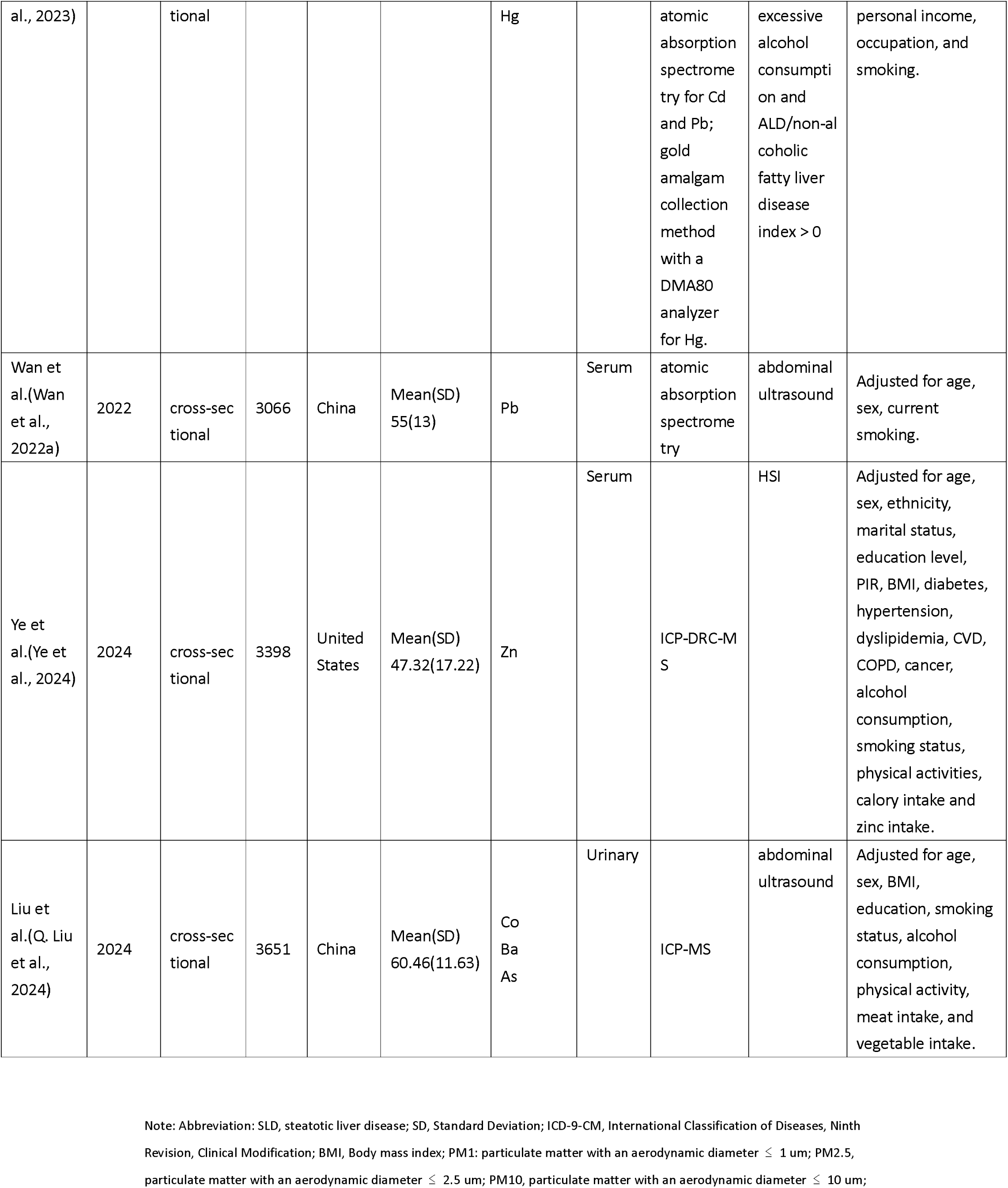
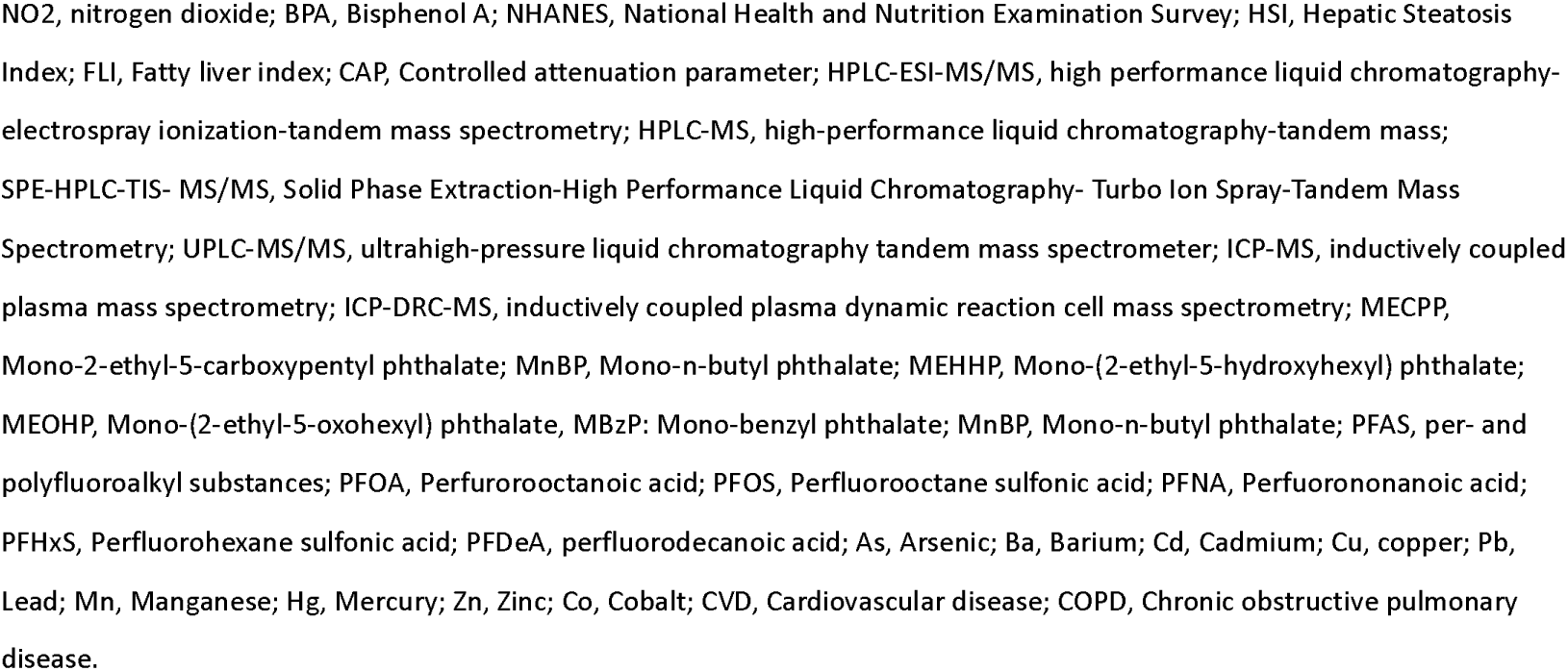
Characteristics of the Included Studies.

### 3.2. Categorization of Included Pollutants

In this study, we evaluated three major categories of environmental pollutants: air pollutants, endocrine-disrupting chemicals (EDCs), and heavy metals. Air pollutants included particulate matter (PM10, PM2.5, PM1) and nitrogen dioxide (NO_2_). EDCs comprised bisphenol A (BPA), phthalate acid esters (PAEs), and Per- and poly-fluoroalkyl substances (PFAS). PAEs were further subdivided into Mono-2-ethyl-5-carboxypentyl phthalate (MECPP), Mono-(2-ethyl-5-hydroxyhexyl) phthalate (MEHHP), Mono-(2-ethyl-5-oxohexyl) phthalate (MEOHP), Mono-n-butyl phthalate (MnBP), and Mono-benzyl phthalate (MBzP). PFAS were categorized as perfluorooctanoic acid (PFOA), perfluorooctane sulfonic acid (PFOS), perfluorononanoic acid (PFNA), perfluorodecanoic acid (PFDeA), and perfluorohexane sulfonic acid (PFHxS). Heavy metals analyzed in our study included barium (Ba), cadmium (Cd), arsenic (As), copper (Cu), mercury (Hg), manganese (Mn), lead (Pb), and cobalt (Co). While Hg, Cd, and Pb are also recognized as EDCs, they were here categorized as heavy metals for consistency and clarity.

### 3.3. Association between exposure to different pollutants and SLD risk

First, we analyzed nine studies(Bo et al., 2024; Cheng et al., 2024; Guo et al., 2022; B. Guo et al., 2023; Han et al., 2023; Ji et al., 2024; Matthiessen et al., 2023; Patterson et al., 2023; VoPham et al., 2022) that reported adjusted effect sizes assessing the association between air pollution exposure and SLD risk. Air pollutant concentrations were reported using three metrics in included studies, including per 10 μg/m³, per 1 μg/m³, and per standard deviation (SD) increase. Among the different exposure units, effect sizes reported per 10 μg/m³ increase were the most frequently used across studies. Therefore, to ensure consistency, maximize study inclusion, and enhance statistical power, we primarily analyzed effect sizes quantified per 10 μg/m³ increase in the main meta-analysis. Notably, the pooled analysis of all included air pollutants revealed a significant association between air pollutant exposure per 10 μg/m³ increase and elevated SLD risk (OR = 1.24, 95% CI: 1.12–1.36) (Figure 2). Specifically, exposure to PM2.5 (OR = 1.23, 95% CI: 1.14–1.32), PM10 (OR = 1.07, 95% CI: –1.13), PM1 (OR = 1.45, 95% CI: 1.04–2.03), and NO_2_ (OR = 1.19, 95% CI: 1.08–1.32) appeared to be associated with an increased SLD risk (Figure 2). In addition, exposure to PM2.5 per 1ug/m^3^ increase and per SD increase was also correlated to the increased SLD risk (eFigure 1 in the Supplement). These results suggest a possible link between air pollution and SLD development.

We next assessed the relationship between EDCs exposure and SLD risk. A pooled analysis of 13 studies revealed a potential association between overall EDCs exposure and increased SLD risk (OR = 1.20; 95%CI: 1.08-1.35) (Figure 3). We further conducted subgroup analysis for BPA(An, Yang, et al., 2021; Kim et al., 2019; Peng et al., 2023), PFAS(Cheng et al., 2023; Wu et al., 2024; Wu et al., 2023; Yun & Min, 2025; J. Zhang et al., 2024; X. Zhang et al., 2023), and PAEs(Cai et al., 2021; Chen et al., 2022; He et al., 2023; Yang, Kim, et al., 2021). Notably, we observed that exposure to BPA (OR: 1.42; 95% CI: 1.23–1.64) and PAEs (OR = 1.29, 95% CI: 1.13–1.47) were respectively associated with increased SLD risk (Figure 3, eFigure 2-4 in the Supplement). For PFAS, only exposure to PFOA (OR = 1.23; 95% CI: 1.14-1.34) showed a possible association. Among different PAEs subtypes, exposure to MECPP (OR = 1.43, 95% CI: 1.20–1.71), MEHHP (OR = 1.54, 95% CI: 1.29–1.84), and MEOHP (OR = 1.38, 95% CI: 1.11–1.73) demonstrated significant associations, whereas MnBP and MBzP did not show statistically significant correlations.

Next, we examined the relationship between heavy metal exposure and SLD risk. The pooled analysis of 12 studies(Fan et al., 2024; Kim et al., 2023; Li et al., 2020; Q. Liu et al., 2024; Maodong et al., 2023; Nguyen & Kim, 2022b; Park et al., 2021; Wan et al., 2022b; Xie et al., 2023; Yang, Yang, et al., 2021; Ye et al., 2024; D. Zhang et al., 2023)identified a significant association between overall heavy metal exposure and increased SLD risk (OR = 1.36, 95% CI: 1.13–1.63) (Figure 4). Specifically, exposure to Pb (OR = 1.61, 95% CI: 1.34–1.93), Cd (OR = 2.32, 95% CI: 1.68–3.20), Hg (OR = 2.41, 95% CI: 1.86–3.11), Ba (OR = 1.16, 95% CI: 1.04–1.30), As (OR = 1.09, 95% CI: –1.16), and Co (OR = 1.18, 95% CI: 1.05–1.34) appeared to be associated with higher risk of SLD. In contrast, no significant associations were observed for Cu (OR = 0.61, 95% CI: 0.30–1.24), Zn (OR = 1.41, 95% CI: 0.93–2.12), and Mn (OR = 0.99, 95% CI: 0.32–3.09) (Figure 4).

### 3.4. Study Quality Assessment, Risk of Bias, and Evidence Level Assessment

The included studies were evaluated as high quality using the JBI and NOS scales (eTables S1–S3 in the Supplement). In addition, the assessment by the OHAT Risk of Bias Rating Tool suggested a low risk of bias among included studies (eAppendix 3-4, eTable S4 in the Supplement). To enhance transparency and rigor, we provided study-specific justifications and domain-level evaluation criteria for each included study (eAppendix 5 in the Supplement). We further evaluated the evidence level of our study according to the NTP/OHAT framework, which assigns an initial “high confidence” rating only to experimental and controlled studies, as they reduce the risk of randomization bias and clearly determine whether exposure precedes the outcome. Since all studies included in our review were observational, including cross-sectional (31), case-control (2), and cohort studies (1), the initial confidence rating was set at “low to moderate.”( Specific details and instructions were listed in the eAppendix 3). After assessing the upgrading and downgrading factors, the final overall confidence in the body of evidence was determined to be “low to moderate” (eTable 5 in the Supplement).

### 3.5. Subgroup Analysis, Sensitivity Analysis, and Publication Bias

Subgroup analyses indicated that differences in SLD diagnostic methods, including the hepatic steatosis index (HSI), abdominal ultrasound, and the controlled attenuation parameter (CAP), as well as covariate adjustments for body mass index (BMI), diabetes, and alcohol consumption, may partially contribute to the observed heterogeneity. For MEOHP, which showed moderate heterogeneity (I^²^ = 47%, τ² = 0.0227), adjustment for BMI reduced heterogeneity to zero (eFigure 5 in the Supplement), highlighting the potential confounding role of BMI in this specific association. Regarding diagnostic methods, studies using CAP for SLD assessment showed substantially lower heterogeneity across PFOA, PFOS, PFNA, and PFDeA, compared to those using HSI or abdominal ultrasound (eFigure 6 in the Supplement), likely reflecting its relatively higher diagnostic sensitivity. In terms of covariate adjustment, models that adjusted for diabetes showed lower heterogeneity compared to those without adjustment (eFigure 7 in the Supplement). However, studies that did not adjust for BMI showed lower heterogeneity (eFigure 8 in the Supplement). This counterintuitive result may be explained by their larger sample sizes, which tend to produce narrower confidence intervals and more stable effect estimates, thereby reducing between-study variability. Furthermore, for PM2.5, PFOS, and PFNA, all of which exhibited high heterogeneity(I² > 90%), adjustment for alcohol consumption markedly reduced heterogeneity to zero. In contrast, for PFOA, which initially showed moderate heterogeneity (I² = 40%, τ² = 0.0002), heterogeneity increased in the alcohol-adjusted subgroup (eFigure 9-10 in the Supplement). Nevertheless, these observations should be interpreted with caution. The limited number of studies may restrict the ability to accurately assess heterogeneity, and the apparent absence of heterogeneity could be due to chance. This underscores the need for larger and more consistently adjusted datasets in future analyses to robustly identify sources of heterogeneity. Sensitivity analysis using the leave-one-out method demonstrated that the overall effect estimates were generally robust across most pollutants, except for PFNA, PFHxS, and Zn(eFigure 11-13 in the Supplement). For PFNA, excluding Wu et al. (2024) reduced heterogeneity from I² = 95% and τ² = 0.2913 to I² = 51% and τ² = 0.0390, resulting in a statistically significant association with SLD (OR = 1.44, 95% CI: 1.11–1.87). For PFHxS, exclusion of either Yun et al. (2024) or Wu et al. (2024) reduced heterogeneity to zero. However, only removal of Wu et al.(2024) resulted in a narrower confidence interval and a positive association (OR = 1.12, 95% CI: 1.04–1.22), suggesting a more stable and precise estimate compared to the inverse association observed when excluding Yun et al.(2024) (OR = 0.83, 95% CI: 0.73–0.95, eFigure 12 in the Supplement). Notably, Wu et al. (2024) was the only case–control study among predominantly cross-sectional designs, suggesting that its study design may have exerted a disproportionate influence on the pooled results. Similarly, for Zn, excluding Zhang et al. (2023), also a case–control study, reversed the direction of association, indicating a potential positive link with SLD risk (OR = 1.64, 95% CI: 1.24–2.17, eFigure 13 in the Supplement). These findings suggest that study design may contribute substantially to heterogeneity and result stability. However, due to the limited number of included studies, formal subgroup analyses by study design were not feasible. Overall, the sensitivity of pooled estimates for PFNA, PFHxS, and Zn warrants cautious interpretation and highlights the need for further validation. To assess publication bias, Egger’s test was first performed and revealed no significant evidence of bias (eTable 6 in the Supplement). Given the small number of studies (n < 3 for some pollutants), funnel plots and the trim-and-fill method were also applied. Additional studies were imputed for most pollutants, except for PFOA, PFOS, PFDeA, MECPP, Ba, and Zn, whose funnel plots appeared symmetrical (eFigure 14–19 in the Supplement). These results suggest potential publication bias in certain subgroups, although the limited number of included studies constrains the reliability of this assessment.

## 4. Discussion

Through systematic review and meta-analysis, this study mapped the association between major environmental pollutants and the risk of SLD. Our findings revealed that exposure to air pollutants (PM1, PM2.5, PM10, NO₂), EDCs (BPA, PFOA, MECPP, MEHHP, and MEOHP), and metals (Pb, Cd, Hg, Ba, As, and Co), whether considered individually or in combination, appears to be associated with an elevated risk of SLD.

According to the WHO, nearly 99% of the global population breathes air containing high levels of pollutants(World Health Organization. WHO air pollution-overview. https://www.who.int/health-topics/air-pollution#tab=tab_1). Air pollution is primarily composed of particulate matter (PM) and gaseous pollutants. A cohort study demonstrated a gradual increase in the odds ratios of chronic liver disease (CLD) with exposure to smaller PM sizes (PM1, PM2.5, PM10)(Chen et al., 2024). Similarly, our analysis identified a gradually elevated SLD risk when exposed to PM1 (OR: 1.45), PM2.5 (OR: 1.23), and PM10 (OR: 1.07) accordingly. Smaller PM is known to have a higher capability of penetrating the bloodstream through alveolar gas exchange, which accumulates in organs, particularly in the liver(Zhang et al., 2016). This may partially explain our findings that smaller PM induced higher SLD risk. Additionally, previous studies suggest that higher spatial resolution and shorter temporal averaging intervals in exposure assessment may help reduce exposure misclassification(Korhonen et al., 2024; Sun & Xia, 2025). In our included studies, long-term exposure was consistently assessed using annual average concentrations across all air pollutants. For PM, the spatial resolution varied among studies (1 km², 10 km², and 12 km²), whereas all NO₂-related studies employed a consistent resolution of 10 km². The pooled estimates for PM2.5 (I² = 93%, τ² = 0.0042), PM10 (I² = 94%, τ² = 0.0017), and PM1 (I² = 99%, τ² = 0.0855) showed substantial heterogeneity, while NO₂ (I² = 68%, τ² = 0.0039) exhibited relatively lower heterogeneity. This pattern reasonably suggests that inconsistent spatial resolution may partially contribute to the observed heterogeneity. However, due to the limited number of studies, we were unable to conduct subgroup analyses to formally test this hypothesis. Notably, the PM1 subgroup also presented a wide confidence interval (1.04–2.03) despite showing a statistically significant association. These findings, while suggestive of a potential dose–response relationship, should be interpreted with caution and confirmed by future studies with larger sample sizes and standardized exposure assessment methodologies. Intriguingly, exposure to PM2.5 has been linked to hepatic lipid accumulation through oxidative stress and inflammatory responses, which disrupt lipid metabolism and contribute to the development of SLD(Xu et al., 2019). Although direct evidence remains limited, several studies suggest that PM2.5 may also impair gut microbiota composition and intestinal barrier integrity, indicating a potential role in disturbing the gut–liver axis and promoting liver injury(Liu et al., 2021; Ran et al., 2024). In addition, our analysis also showed that exposure to gaseous pollutant NO_2_ was associated with an increased risk of SLD. From mechanism-of-action, NO_2_ exposure has been shown to disrupt galactose metabolism and promote hepatic lipogenesis, contributing to hepatic fat accumulation and liver injury(Y. Guo et al., 2023). Moreover, frequent exposure to cooking oil fumes and secondhand smoke were also associated with increased risk of SLD(Lin et al., 2020; E. Liu et al., 2024). Beyond disease onset, air pollution has been implicated in SLD progression, for example, contributing to advanced liver fibrosis and cirrhosis (Li et al., 2023; Xing et al., 2023). Collectively, these findings highlight the multifactorial impact of air pollution on liver health.

Endocrine-disrupting chemicals (EDCs) are widely present in pesticides, metals, food, and personal care products(Monneret, 2017). They are capable of interfering with the hormone actions of the human endocrine system. Exposure to EDCs has been linked to various health conditions, including reproductive impairment, metabolic disorders, and cancers(La Merrill et al., 2020). Our analysis further demonstrated that exposure to EDCs, specifically BPA, PFAS, and PAEs, is associated with an elevated SLD risk. BPA, primarily used in the manufacture of polycarbonate plastics and epoxy resins, has been identified to dysregulate hepatic lipid homeostasis, contributing to metabolic dysfunction–associated SLD (MASLD)(Dallio et al., 2019). A recent experimental study demonstrated that lowZIdose BPA exposure suppresses hepatic PPARγ activity and significantly downregulates Cpt1a expression in mouse liver, disrupting fatty acid oxidation and promoting lipid accumulation in hepatocytes(Zhu et al., 2025). In our analysis, BPA exposure was associated with an increased risk of SLD (OR = 1.42), with no observed heterogeneity (I² = 0%, τ² = 0%), suggesting a relatively consistent association across the included studies.

PFAS, a group of highly persistent synthetic chemicals, have been reported to alter lipid metabolism and de novo lipogenesis in human hepatocytes(Kashobwe et al., 2024). Recent evidence further revealed that PFOA may disrupt hepatic immunometabolic homeostasis through modulation of PPARα and PPARγ signaling pathways, leading to suppressed M2 macrophage activation and subsequent promotion of inflammation and lipid dysregulation(You et al., 2025). In animal models, PFAS exposure has been shown to induce hepatic steatosis through gut microbiota dysbiosis and compromised intestinal barrier integrity, resulting in systemic inflammation. Additionally, PFAS interfere with metabolic signaling and bile acid homeostasis, characterized by upregulation of lipogenic genes and downregulation of genes involved in fatty acid oxidation(Chen et al., 2025; Gu et al., 2024). This proposed mechanism may collectively provide a biological explanation for the observed association between PFOA exposure and increased risk of SLD in our meta-analysis. In contrast, other PFAS compounds did not show significant associations, which may be attributable to non-linear dose–response effects.

PAEs are abused as plasticizers in the industrial production of plastics or other related products(Wang et al., 2023). Importantly, three types of PAEs— di-(2-ethylhexyl) phthalate (DEHP), dibutyl phthalate (DBP), and butyl benzyl phthalate (BBzP), have been shown to adversely affect liver metabolic functions(An, Lee, et al., 2021), (Theodoropoulou et al., 2024), (Vacy et al., 2025). After entering the body, PAEs undergo hydrolysis into monoester phthalates (mPAEs) metabolites by liver microsomes(Hanioka et al., 2012). In our study, DEHP metabolites MEHHP, MECPP, and MEOHP were found to be significantly associated with increased SLD risk, aligning with a previous study based on NHANES(Zou et al., 2024). However, no significant associations were observed for the DBP metabolite MnBP or the BBzP metabolite MBzP. Although these findings underscore the role of specific PAEs in SLD risk, large-scale population-based studies are warranted to further elucidate these associations.

Heavy metals have increasingly been released into the environment primarily by anthropogenic activities such as mining, industrial production, and agriculture(Briffa et al., 2020). Notably, these metals do not degrade and can bioaccumulate in the human body along the food chain(Dar et al., 2017). Emerging evidence suggests that heavy metals contribute to multi-organ toxicity, including hepatotoxicity, cardiovascular toxicity, and neurotoxicity(Garza-Lombó et al., 2019; Hyder et al., 2013; Zefferino et al., 2017). In our study, we observed positive associations between exposure to Pb, Cd, Hg, Ba, As, and Co and an increased risk of SLD. Although no similar association was observed for Zn, Cu, and Mn, this is seemingly due to limited studies with high heterogeneity. Heavy metals are speculated to preferentially accumulate in the liver, leading to hepatic injury through complex mechanistic steps. For instance, heavy metals such as As, Pb, Cd, and Ba have been associated with increased ROS generation, leading to lipid peroxidation, mitochondrial dysfunction, and hepatocyte apoptosis(Elwej et al., 2017; Teschke, 2022). Co has been reported to disrupt fatty acid metabolism by altering fatty acid composition and interfering with key metabolic pathways such as desaturation and elongation(Ghribi et al., 2025). In addition, heavy metals such as Cd and Pb have been shown to markedly disturb intestinal microbial composition and damage epithelial integrity, leading to increased translocation of bacterial endotoxins, which in turn activate hepatic immune responses and promote steatosis and inflammation(Ghosh et al., 2024). Furthermore, a recent cross-sectional study further demonstrated an association between heavy metal exposure and early liver dysfunction among rural residents in China(Yang et al., 2022).

In addition to the pollutants analyzed in this study, numerous other environmental pollutants also pose threats to public health. Among them, microplastics and nanoplastics, which originate from the widespread plastic use, have emerged as a major concern. These plastic particles are ubiquitous and can infiltrate human tissues through food, water, and air(van der Laan et al., 2023). Emerging evidence from a recent in vivo murine study suggests that nanoplastics may pose hepatotoxic risks by disrupting hepatic lipid metabolism through inhibition of PPARγ, thereby inducing oxidative stress, inflammation, and lipid accumulation(Jiang et al., 2025).

Collectively, our synthesis of the evidence indicates that environmental pollutants promote the development of SLD primarily through three biological mechanisms: oxidative stress and inflammation, lipid metabolism disruption, and gut–liver axis dysregulation. However, these mechanistic pathways are not mutually exclusive but rather interconnected. A single pollutant may act through multiple biological routes to promote hepatic steatosis, emphasizing the complex and multifactorial nature of pollutant-induced liver injury. Furthermore, mechanistic studies remain limited for many environmental pollutants, highlighting the need for continued investigation to elucidate their roles in the pathogenesis of SLD.

## 5. Public Health and Regulatory Implications

Finally, coordinated public health interventions are essential to address the health risks associated with environmental pollutants. The 2021 WHO Air Quality Guidelines provide evidence-based threshold values for six key air pollutants, including PM2.5, PM10, NO_2_, SO_2_, O_3_, and CO(World Health Organization, 2021). For PM2.5, the recommended annual mean concentration is 5 μg/m³, a level frequently exceeded in many urban and industrial areas. Our meta-analysis showed that each 10 μg/m³ increment in PM2.5 was associated with a 23% increased risk of SLD (OR = 1.23), and even a 1 μg/m³ increase corresponded to a 2% rise in risk (OR = 1.02), highlighting the substantial public health impact of PM2.5 exposure. For NO₂ and PM10, every 10 μg/m³ increase was associated with 19% and 7% higher SLD risks, respectively, although 1 μg/m³ increments did not yield statistically significant associations. These findings suggest that maintaining PM10 and NO₂ concentrations below guideline thresholds (15 μg/m³ and 10 μg/m³, respectively) may also help reduce SLD burden. Although the WHO air quality guidelines are not legally binding, they provide a vital framework for national air quality management, emphasizing the importance of regular monitoring, emission control, and interregional coordination. In contrast, comparable international consensus thresholds for chemical contaminants remain limited due to their diverse structures and toxicological behaviors. For example, the U.S. Environmental Protection Agency (EPA) recommends several achievable steps to reduce PFAS exposure, including using certified water filters capable of removing PFAS, avoiding stain-resistant textiles and non-stick cookware, and limiting intake of highly processed and packaged foods(U.S. Environmental Protection Agency, 2024). Other chemicals, including BPA and PAEs, and certain metals, recommended measures include phasing out high-risk substances, strengthening the regulation of food contact materials, promoting safer alternatives, and encouraging reduced plastic use and minimally processed diets(Muncke et al., 2025). Collectively, these efforts are integral to a broader strategy aimed at reducing environmental exposures and improving population health.

## 6. Limitation

This study has several limitations. Firstly, high heterogeneity was observed in some analyses, likely due to the limited studies available for each pollutant. This also restricted our ability to perform subgroup and sensitivity analyses to better investigate potential sources of heterogeneity. Therefore, although our study identified several statistically significant associations, these should be regarded as preliminary signals rather than definitive evidence and should be interpreted with appropriate caution. Secondly, most included studies were cross-sectional, limiting causal inference due to the lack of temporality. As these studies typically assess disease prevalence rather than incidence, they are also more susceptible to confounding and require adjustment for multiple potential covariates. Thirdly, variations in SLD diagnostic methods, pollutant exposure assessments, population characteristics, and adjusted covariates may induce bias and limit the robustness of our conclusions. Fourthly, our analysis focused on three major pollutant categories, while data on other environmental pollutants remain insufficient. Finally, only English-language studies were included, potentially leading to the exclusion of relevant research published in other languages.

## 7. Conclusion

This study demonstrated that exposure to environmental pollutants, including air pollutants (PM1, PM2.5, PM10, NO_2_), endocrine-disrupting chemicals (BPA, PFOA, MECPP, MEHHP, and MEOHP), and heavy metals (Pb, Cd, Hg, Ba, As and, Co), appeared to be associated with an increased risk of SLD. However, the predominance of cross-sectional study designs substantially limits the ability to infer causality, thereby weakening the overall strength of the evidence. High-quality prospective studies are urgently needed to confirm these associations and clarify temporal relationships.

## Data Availability

All data produced in the present work are contained in the manuscript

## CRediT authorship contribution statement

Xincheng Li: Concept and design, acquisition, analysis, or interpretation of data, Drafting of the manuscript; Jiajing Li: Concept and design, acquisition, analysis, or interpretation of data, writing-reviewing and editing; Jiangrong Zhou: Concept and design, acquisition, analysis, or interpretation of data, writing-reviewing and editing; Ibrahim Ayada: Concept and design, writing-reviewing and editing; Qiuwei Pan: Concept and design, writing-reviewing and editing; Luc J.W. van der Laan: Supervision, Concept and design, writing-reviewing and editing; Pengfei Li: Supervision, Concept and design, writing-reviewing and editing.

---

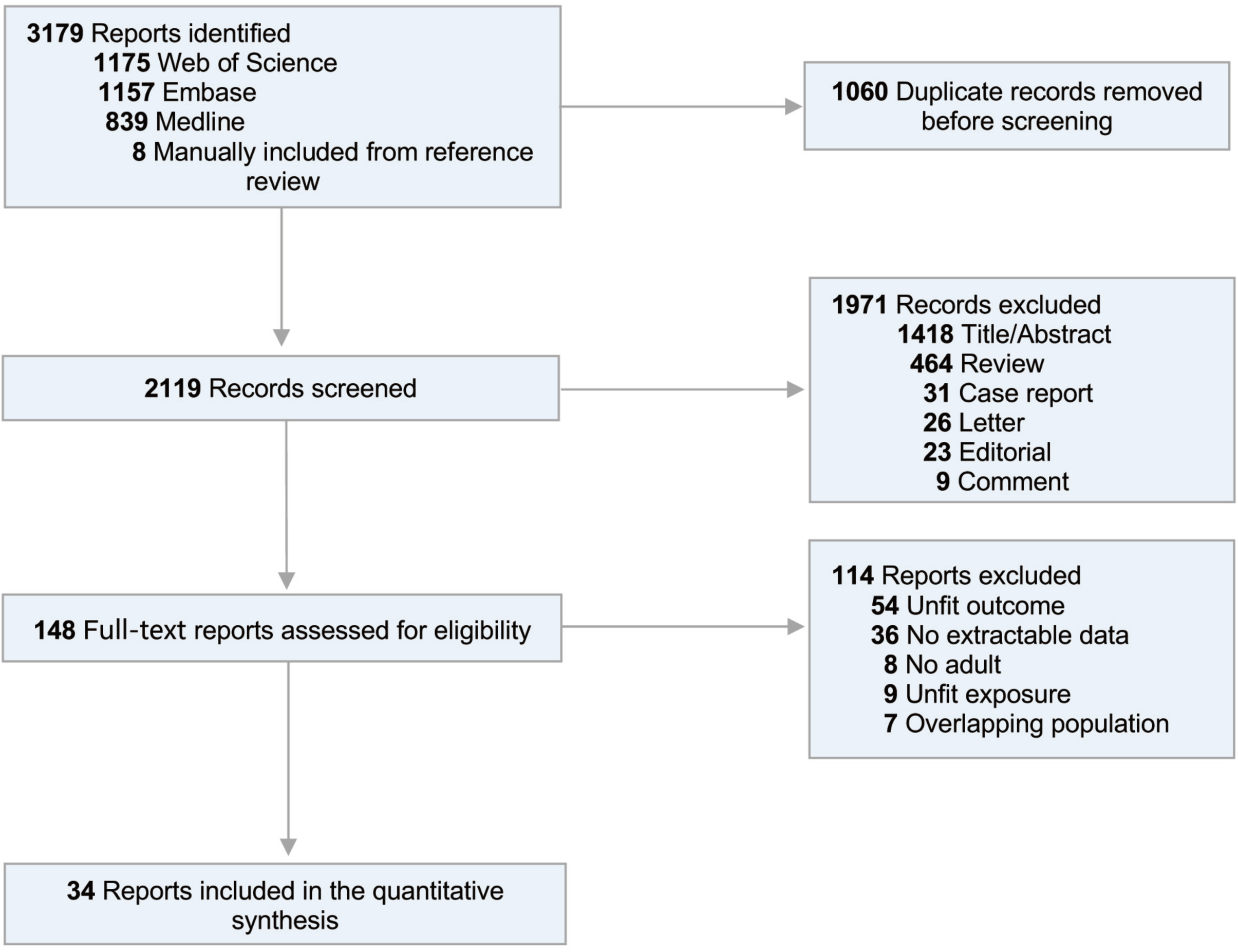

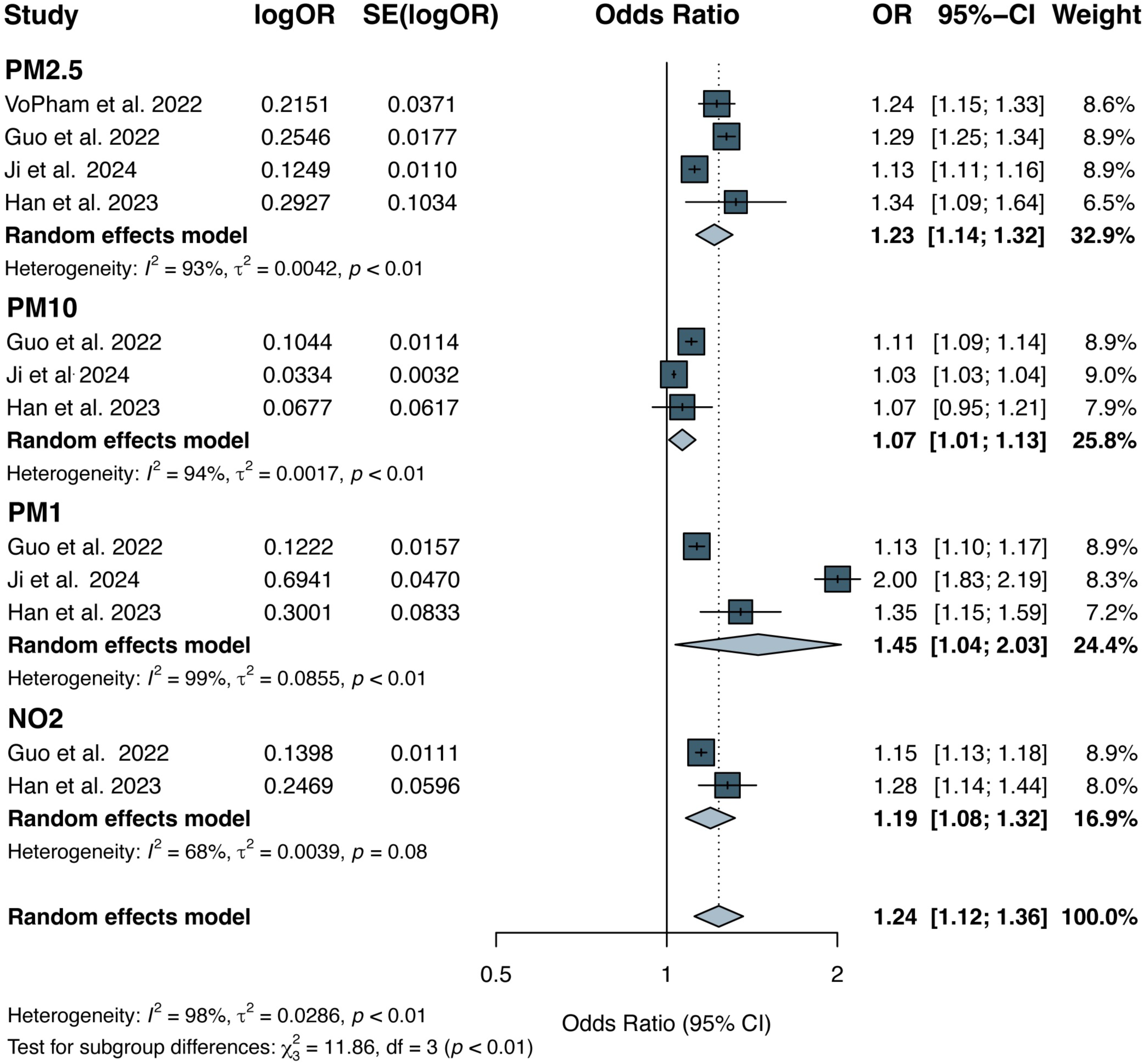

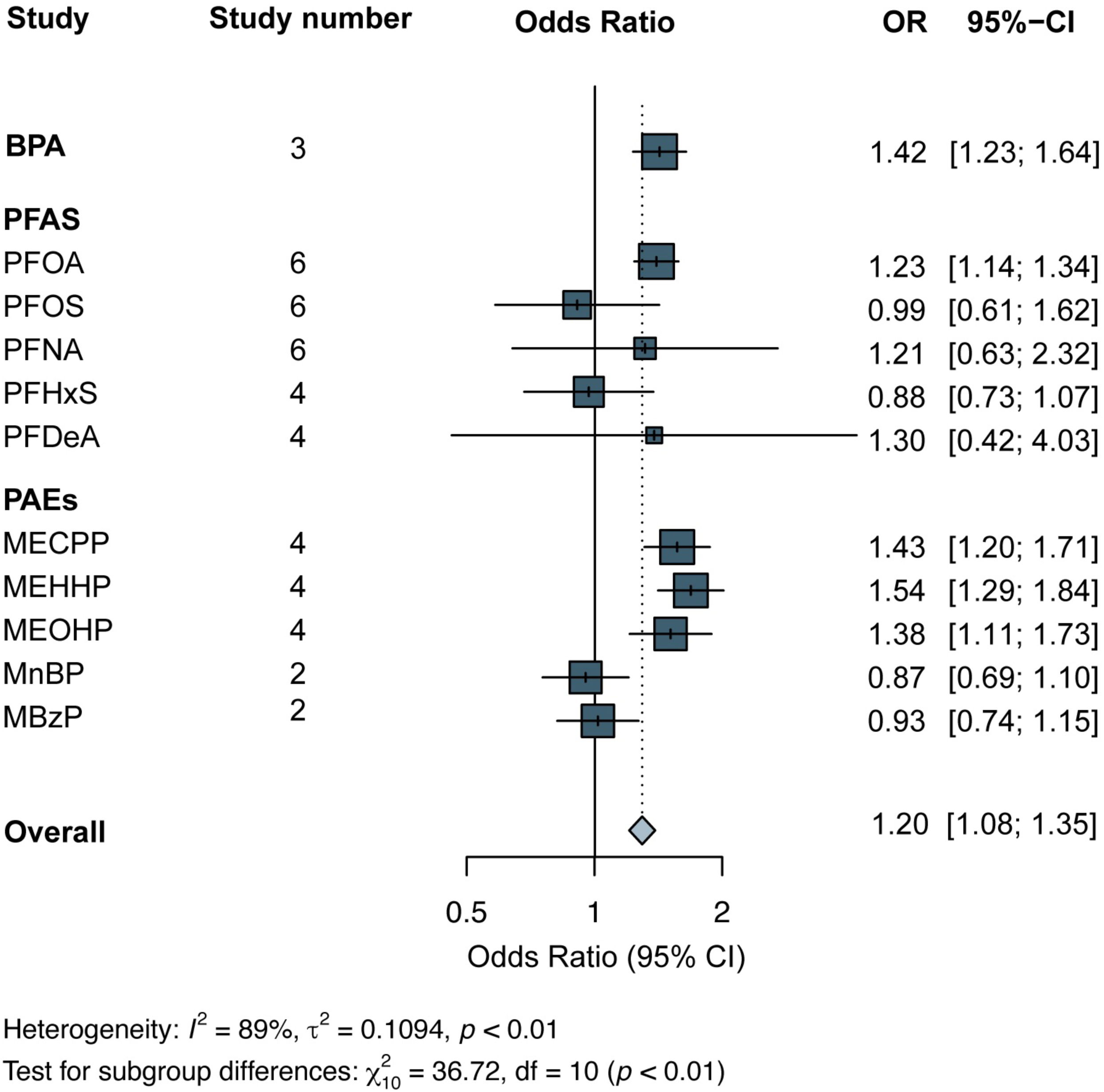

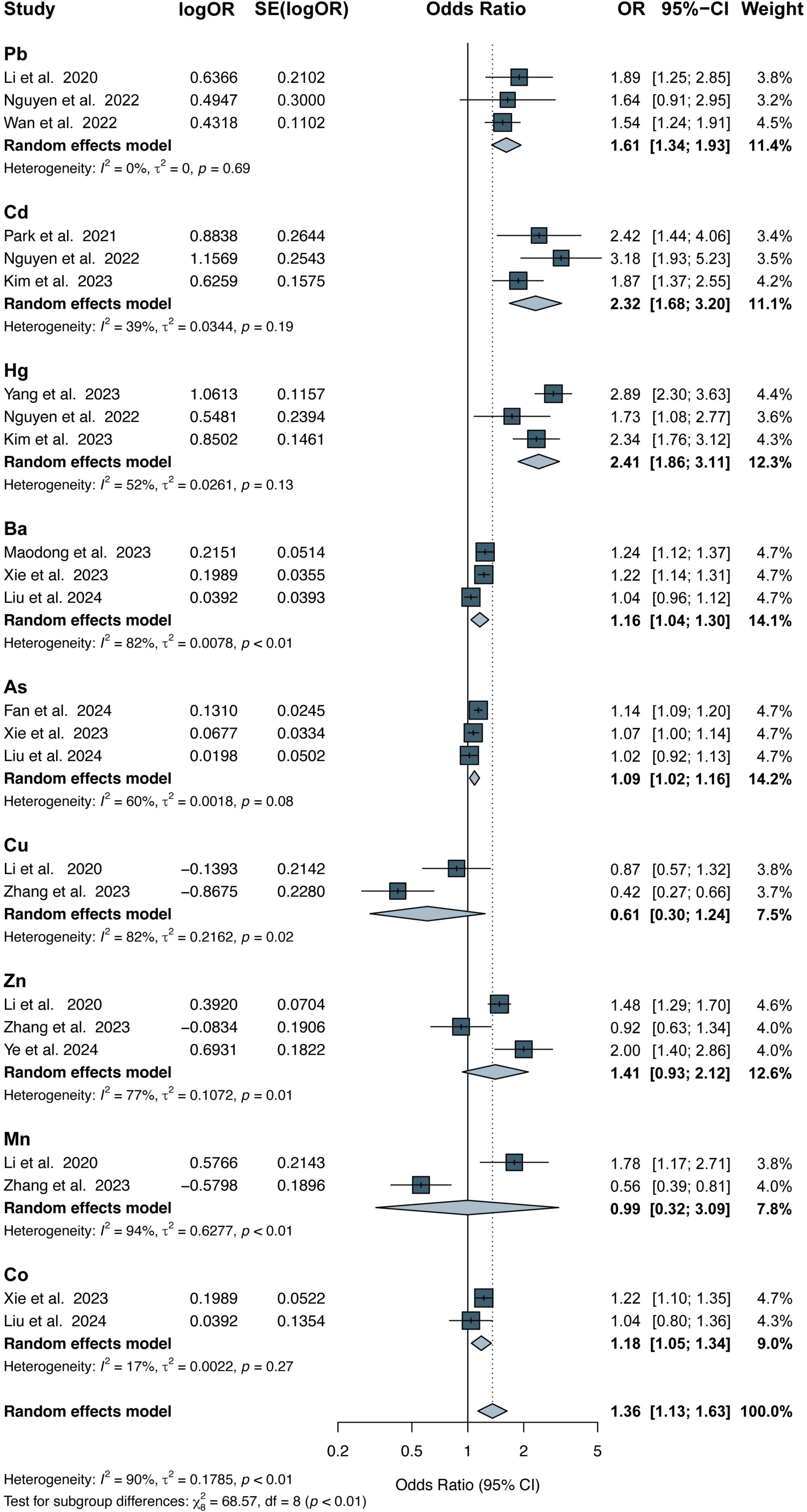

