## Supplementary file for "Mapping the association between environmental pollutants and steatotic liver disease: a systematic review and meta-analysis"

Li et al.

**eAppendix 1.** Literature search strategies
**eAppendix 2.** Inclusion and exclusion criteria
**eAppendix 3.** OHAT Risk-of-Bias Criteria
**eAppendix 4.** Confidence in the body of evidence and the level of evidence Criteria
**eAppendix 5.** Detailed risk of bias results of the included studies.
**eTable 1.** Joanna Briggs Institute (JBI) checklist (applied to cross-sectional studies)
**eTable 2.** The Newcastle–Ottawa Scale (NOS) checklist (applied to case-control studies)
**eTable 3.** The Newcastle–Ottawa Scale (NOS) checklist (applied to cohort studies)
**eTable 4.** Risk of bias for included studies
**eTable 5.** Confidence in the level of evidence
**eTable 6.** Estimation of publication bias
**eFigure 1.** Forest plot ORs (95% CI) for the association between PM2.5 per 1ug/m3 increase and per SD increase and SLD
**eFigure 2.** Forest plot ORs (95% CI) for the association between Bisphenol A (BPA) and SLD.
**eFigure 3.** Forest plot ORs (95% CI) for the association between perfluoroalkyl substance (PFAS) exposure and SLD
**eFigure 4.** Forest plot ORs (95% CI) for the association between phthalate esters (PAEs)exposure and SLD
**eFigure 5.** Subgroup Analysis of MEOHP and SLD Risk Based on Adjustment for BMI
**eFigure 6.** Subgroup Analysis of PFAS Exposure and SLD Risk Based on Diagnostic Methods
**eFigure 7.** Subgroup Analysis of PFAS Exposure and SLD Risk Based on Adjustment for diabetes
**eFigure 8.** Subgroup Analysis of PFAS Exposure and SLD Risk Based on Adjustment for BMI
**eFigure 9.** Subgroup Analysis of PM2.5 and SLD Risk Based on Adjustment for Alcohol Consumption
**eFigure 10.** Subgroup Analysis of PFAS and SLD Risk Based on Adjustment for Alcohol Consumption
**eFigure 11.** Sensitivity analysis of air pollutants (per 10ug/m3 increase)
**eFigure 12.** Sensitivity analysis of PFAS
**eFigure 13.** Sensitivity analysis of heavy metals
**eFigure 14.** Funnel Plots of Air Pollutants Before and After Trim-and-Fill Adjustment
**eFigure 15.** Funnel Plots of PFAS Before and After Trim-and-Fill Adjustment
**eFigure 16.** Funnel Plots of PAEs Before and After Trim-and-Fill Adjustment
**eFigure 17.** Funnel Plot of BPA Before and After Trim-and-Fill Adjustment
**eFigure 18.** Funnel Plots of Metal Pollutants (n < 3)
**eFigure 19.** Funnel Plots of Metal Pollutants (n ≥ 3)

#### eAppendix 1. Literature search strategies

| Database searched | Platform | Years of coverage | Records | Records after duplicates removed |
|---|---|---|---|---|
| Medline ALL | Ovid | 1946 - Present | 839 | 839 |
| Embase | Embase.com | 1971 - Present | 1157 | 564 |
| Web of Science Core Collection* | Web of Knowledge | 1975 - Present | 1175 | 708 |
| Manually screened | | | 8 | |
| **Total** | | | **3179** | **2119** |

*Science Citation Index Expanded (1975-present); Social Sciences Citation Index (1975-present); Arts & Humanities Citation Index (1975-present); Conference Proceedings Citation Index- Science (1990-present); Conference Proceedings Citation Index- Social Science & Humanities (1990-present); Emerging Sources Citation Index (2005-present)

No other database limits were used than those specified in the search strategies

The research date ended on May 30th, 2025.

**medline 839**

(Non-alcoholic Fatty Liver Disease / OR Fatty Liver / OR (((metabol* OR steato*) ADJ6 (liver* OR hepatic*) ADJ6 (disease* OR dysfunction*)) OR ((nonalcohol* OR non-alcohol*) ADJ3 (fatty-liver*)) OR steatohepatit* OR (steato* ADJ3 (hepat* OR liver*)) OR nafld OR (lipid* ADJ6 (liver* OR hepatic*) ADJ6 (level* OR concentrat*))).ab,ti,kf. OR (((metabol* OR steato* OR fatty*) AND (liver* OR hepatic*) AND (disease* OR dysfunction*)) OR fatty-liver*).ti.) AND (Environmental Pollutants/ OR Environmental Exposure/ OR Microplastics/ OR Environmental Pollution/ OR exp Air Pollution/ OR Particulate Matter/ OR Food Contamination/ OR Pesticides/ OR Fungicides, Industrial/ OR Herbicides/ OR Insecticides/ OR Rodenticides/ OR exp * Pesticides/ OR Water Quality/ OR (((environment* OR air OR water* OR soil* OR chemical* OR industr* OR food* OR exposure* OR agriculture* OR plastic*) ADJ3 (pollut* OR contamin* OR residue*)) OR ((environment* OR traffic* OR exhaust* OR fumes* OR emission* OR chemical*) ADJ3 (exposure* OR risk* OR toxic*)) OR ((environment*) ADJ3 (traffic* OR exhaust* OR fumes* OR emission* OR chemical*)) OR microplastic* OR micro-plastic* OR nanoplastic* OR nano-plastic* OR (particulate* ADJ3 matter*) OR pm1 OR pm10 OR pm2-5 OR pm-1 OR pm-10 OR pm-2-5 OR toxicant* OR fluorocarbon* OR fluoro-carbon* OR pesticide* OR fungicide* OR herbicide* OR insecticide* OR rodenticide* OR Perfluoroalkyl* OR polyfluoroalkyl* OR Perfluoro-alkyl* OR polyfluoro-alkyl* OR pfas OR organohalogen* OR (exhaust ADJ (gas* OR fumes*)) OR glyphosate* OR ((ambient* OR indoor* OR quality) ADJ3 air) OR ((quality) ADJ3 (water*)) OR Perchlorate* OR Bisphenol* OR Heterocyclic* OR Plastic-compound*).ab,ti,kf. OR (pollut* OR contamin*).ti.) NOT (exp animals/ NOT humans/) NOT (news OR congres* OR abstract* OR book* OR chapter* OR dissertation abstract*).pt. AND english.la.

**Embase 1157**

('metabolic liver disease'/de OR 'nonalcoholic fatty liver'/exp OR 'fatty liver'/de OR 'lipid liver level'/exp OR (((metabol* OR steato*) NEAR/6 (liver* OR hepatic*) NEAR/6 (disease* OR dysfunction*)) OR ((nonalcohol* OR non-alcohol*) NEAR/3 (fatty-liver*)) OR steatohepatit* OR (steato* NEAR/3 (hepat*

OR liver*)) OR nafld OR (lipid* NEAR/6 (liver* OR hepatic*) NEAR/6 (level* OR concentrat*))):ab,ti,kw OR (((metabol* OR steato* OR fatty*) AND (liver* OR hepatic*) AND (disease* OR dysfunction*)) OR fatty-liver*):ti) AND ('environmental pollutant'/de OR 'environmental exposure'/de OR microplastic/exp OR 'pollution and pollution related phenomena'/exp OR 'particulate matter'/exp OR 'food contamination'/exp OR pesticide/de OR fungicide/de OR herbicide/de OR insecticide/de OR rodenticide/de OR pesticide/exp/mj OR 'organohalogen derivative'/de OR 'ambient air'/de OR 'air quality'/de OR 'water quality'/de OR 'environmental parameters'/mj OR (((environment* OR air OR water* OR soil* OR chemical* OR industr* OR food* OR exposure* OR agriculture* OR plastic*) NEAR/3 (pollut* OR contamin* OR residue*)) OR ((environment* OR traffic* OR exhaust* OR fumes* OR emission* OR chemical*) NEAR/3 (exposure* OR risk* OR toxic*)) OR ((environment*) NEAR/3 (traffic* OR exhaust* OR fumes* OR emission* OR chemical*)) OR microplastic* OR micro-plastic* OR nanoplastic* OR nano-plastic* OR (particulate* NEAR/3 matter*) OR pm1 OR pm10 OR pm2-5 OR pm-1 OR pm-10 OR pm-2-5 OR toxicant* OR fluorocarbon* OR fluoro-carbon* OR pesticide* OR fungicide* OR herbicide* OR insecticide* OR rodenticide* OR Perfluoroalkyl* OR polyfluoroalkyl* OR Perfluoro-alkyl* OR polyfluoro-alkyl* OR pfas OR organohalogen* OR (exhaust NEXT/1 (gas* OR fumes*)) OR glyphosate* OR ((ambient* OR indoor* OR quality) NEAR/3 air) OR ((quality) NEAR/3 (water*)) OR Perchlorate* OR Bisphenol* OR Heterocyclic* OR Plastic-compound*):ab,ti,kw OR (pollut* OR contamin*):ti) NOT ([animals]/lim NOT [humans]/lim) NOT ([conference abstract]/lim) AND [english]/lim

**Web of science 1175**

(TS=(((metabol* OR steato*) NEAR/5 (liver* OR hepatic*) NEAR/5 (disease* OR dysfunction*)) OR ((nonalcohol* OR non-alcohol*) NEAR/2 (fatty-liver*)) OR steatohepatit* OR (steato* NEAR/2 (hepat* OR liver*)) OR nafld OR (lipid* NEAR/5 (liver* OR hepatic*) NEAR/5 (level* OR concentrat*))) OR TI=(((metabol* OR steato* OR fatty*) AND (liver* OR hepatic*) AND (disease* OR dysfunction*)) OR fatty-liver*)) AND (TS=(((environment* OR air OR water* OR soil* OR chemical* OR industr* OR food* OR exposure* OR agriculture* OR plastic*) NEAR/2 (pollut* OR contamin* OR residue*)) OR ((environment* OR traffic* OR exhaust* OR fumes* OR emission* OR chemical*) NEAR/2 (exposure* OR risk* OR toxic*)) OR ((environment*) NEAR/2 (traffic* OR exhaust* OR fumes* OR emission* OR chemical*)) OR microplastic* OR micro-plastic* OR nanoplastic* OR nano-plastic* OR (particulate* NEAR/2 matter*) OR pm1 OR pm10 OR pm2-5 OR pm-1 OR pm-10 OR pm-2-5 OR toxicant* OR fluorocarbon* OR fluoro-carbon* OR pesticide* OR fungicide* OR herbicide* OR insecticide* OR rodenticide* OR Perfluoroalkyl* OR polyfluoroalkyl* OR Perfluoro-alkyl* OR polyfluoro-alkyl* OR pfas OR organohalogen* OR (exhaust NEAR/1 (gas* OR fumes*)) OR glyphosate* OR ((ambient* OR indoor* OR quality) NEAR/2 air) OR ((quality) NEAR/2 (water*)) OR Perchlorate* OR Bisphenol* OR Heterocyclic* OR Plastic-compound*) OR TI=(pollut* OR contamin*)) AND DT=(article) AND LA=(english)

#### eAppendix 2. Inclusion and exclusion criteria

| |
|---|
| **Inclusion criteria** |
| 1. Observational studies (e.g., cohort, case-control, cross-sectional, longitude) reporting quantitative effect estimates (e.g., odds ratios [OR], relative risks [RR], hazard ratios [HR]) that examine the association between various environmental pollutants and the incidence or prevalence of steatotic liver disease (SLD) |
| 2. Studies conducted on adult populations |
| 3. Studies in which participants experienced environmental exposure before disease onset |
| **Exclusion criteria** |
| 1. Publications such as case reports, letters, reviews, case series, editorials, and commentaries |
| 2. Studies focusing on animal models or in vitro experiments |
| 3. Studies that do not report quantitative effect estimates (e.g., OR, RR, HR values, and 95% confidence intervals) |
| 4. Studies where the outcome is not SLD |
| 5. Studies where the exposure substance is not relevant to the topic |
| 6. Non-English publications |

#### eAppendix3. OHAT Risk-of-Bias Criteria

The NTP/OHAT risk of bias framework uses a tiered system that emphasizes key domains considered most relevant for assessing individual studies. In observational human research, these critical domains commonly include how exposures and outcomes are measured, as well as how confounding and selection biases are handled.

##### 1. This tool evaluates each study based on a set of 7 risk-of-bias questions:

| |
|---|
| 1. **Confounding bias [Key item]:** Did the study design or analysis (stratification, regression, matching, inverse probability weighing, etc.) account for important confounding and modifying variables? |
| 2. **Selection bias:** Did selection of study participants result in appropriate comparison groups? |
| 3. **Attrition/exclusion bias:** Were outcome data incomplete due to attrition or exclusion from analysis? |
| 4. **Detection bias [Key item]:** Can we be confident in the exposure characterization? |
| 5. **Detection bias [Key item]:** Can we be confident in the outcome assessment? |
| 6. **Selective reporting bias:** Were all measured outcomes reported? |
| 7**. Conflict of interest:** Was the study free of support from a company, study author, or other entity having a financial interest in any of the treatments studied? |

| **Selection bias** | Did selection of study participants result in appropriate comparison groups? |
|---|---|
| Definitely low risk of bias | There is direct evidence that subjects (both exposed and non-exposed) were similar (e.g., recruited from the same eligible population, recruited with the same method of ascertainment using the same inclusion and exclusion criteria, and were of similar age and health status), recruited within the same time frame, and had similar participation/response rates.<br>There is direct evidence that cases and controls were similar (e.g., recruited from the same eligible population including being of similar age, gender, ethnicity, and eligibility criteria other than outcome of interest as appropriate), recruited within the same time frame, and controls are described as having no history of the outcome.<br>A study will be considered low risk of bias if baseline characteristics of groups differed, but these differences were considered as potential confounding or stratification variables. |
| Probably low risk of bias | There is indirect evidence that subjects (both exposed and non-exposed) were similar (e.g., recruited from the same eligible population, recruited with the same method of ascertainment using the same inclusion and exclusion criteria, and were of similar age and health status), recruited within the same time frame, and had similar participation/response rates, OR differences between groups would not appreciably bias results. |
| Probably high risk of bias | There is indirect evidence that subjects (both exposed and non-exposed) were not similar, recruited within very different time frames, or had very different participation/response rates, OR there is insufficient information provided about the comparison group including a different rate of non-response without an explanation (record "NR" as basis for answer). |

| | |
|---|---|
| Definitely high risk of bias | There is direct evidence that subjects (both exposed and non-exposed) were not similar, recruited within very different time frames, or had very different participation/response rates |
| **Attrition/exclusion bias** | Were outcome data complete without attrition or exclusion from analysis? |
| Definitely low risk of bias | There is direct evidence that loss of subjects (i.e., incomplete outcome data) was adequately addressed and reasons were documented when human subjects were removed from a study. Acceptable handling of subject attrition includes: very little missing outcome data; reasons for missing subjects unlikely to be related to outcome (for survival data, censoring unlikely to be introducing bias); missing outcome data balanced in numbers across study groups, with similar reasons for missing data across groups, OR missing data have been imputed using appropriate methods and characteristics of subjects lost to follow up or with unavailable records are described in identical way and are not significantly different from those of the study participants. |
| Probably low risk of bias | There is indirect evidence that loss of subjects (i.e., incomplete outcome data) was adequately addressed and reasons were documented when human subjects were removed from a study, OR it is deemed that the proportion lost to follow-up would not appreciably bias results. This would include reports of no statistical differences in characteristics of subjects lost to follow up or with unavailable records from those of the study participants. Generally, the higher the ratio of participants with missing data to participants with events, the greater potential there is for bias. For studies with a long duration of follow-up, some withdrawals for such reasons are inevitable. |
| Probably high risk of bias | There is indirect evidence that loss of subjects (i.e., incomplete outcome data) was unacceptably large and not adequately addressed, OR there is insufficient information provided about numbers of subjects lost to follow-up (record “NR” as basis for answer). |
| Definitely high risk of bias | There is direct evidence that loss of subjects (i.e., incomplete outcome data) was unacceptably large and not adequately addressed. Unacceptable handling of subject attrition includes reason for missing outcome data likely to be related to true outcome, with either imbalance in numbers or reasons for missing data across study groups; or potentially inappropriate application of imputation. |
| **Detection bias** | Can we be confident in the exposure characterization? |
| Definitely low risk of bias | There is direct evidence that exposure was consistently assessed (i.e., under the same method and timeframe) using well-established methods that directly measure exposure (e.g., measurement of the chemical in air or measurement of the chemical in blood, plasma, urine, etc.), OR exposure was assessed using less-established methods that directly measure exposure and are validated against well-established methods. |
| Probably low risk of bias | There is indirect evidence that the exposure was consistently assessed using well-established methods that directly measure exposure, OR exposure |

| | |
|---|---|
| | was assessed using indirect measures (e.g., questionnaire or occupational exposure assessment by a certified industrial hygienist) that have been validated or empirically shown to be consistent with methods that directly measure exposure (i.e., inter-methods validation: one method vs. another). |
| Probably high risk of bias | There is indirect evidence that the exposure was assessed using poorly validated methods that directly measure exposure, OR there is direct evidence that the exposure was assessed using indirect measures that have not been validated or empirically shown to be consistent with methods that directly measure exposure (e.g., a job-exposure matrix or self-report without validation)(record "NR" as basis for answer),OR there is insufficient information provided about the exposure assessment, including validity and reliability, but no evidence for concern about the method used (record "NR" as basis for answer) |
| Definitely high risk of bias | There is direct evidence that the exposure was assessed using methods with poor validity, OR evidence of exposure misclassification (e.g., differential recall of self-reported exposure) |
| **Outcome assessment** | Can we be confident in the outcome assessment? |
| Definitely low risk of bias | There is direct evidence that the outcome was assessed using well-established methods (e.g., the "gold standard" with validity and reliability >0.70), AND subjects had been followed for the same length of time in all study groups. Acceptable assessment methods will depend on the outcome, but examples of such methods may include: objectively measured with diagnostic methods, measured by trained interviewers, obtained from registries AND there is direct evidence that the outcome assessors (including study subjects, if outcomes were self-reported) were adequately blinded to the study group, and it is unlikely that they could have broken the blinding prior to reporting outcomes. |
| Probably low risk of bias | There is indirect evidence that the outcome was assessed using acceptable methods (i.e., deemed valid and reliable but not the gold standard) (e.g., validity and reliability ≥0.40), AND subjects had been followed for the same length of time in all study groups [Acceptable, but not ideal assessment methods will depend on the outcome, but examples of such methods may include proxy reporting of outcomes and mining of data collected for other purposes],OR it is deemed that the outcome assessment methods used would not appreciably bias results, AND there is indirect evidence that the outcome assessors (including study subjects, if outcomes were self-reported) were adequately blinded to the study group, and it is unlikely that they could have broken the blinding prior to reporting outcomes, OR it is deemed that lack of adequate blinding of outcome assessors would not appreciably bias results, which is more likely to apply to objective outcome measures. |
| Probably high risk of bias | There is indirect evidence that the outcome assessment method is an insensitive instrument (e.g., a questionnaire used to assess outcomes with no information on validation), OR the length of follow up differed by study group, |

| | |
|---|---|
| | OR there is indirect evidence that it was possible for outcome assessors (including study subjects if outcomes were self-reported) to infer the study group prior to reporting outcomes, OR there is insufficient information provided about blinding of outcome assessors (record “NR” as basis for answer) |
| Definitely high risk of bias | There is direct evidence that the outcome assessment method is an insensitive instrument, OR the length of follow up differed by study group, OR there is direct evidence for lack of adequate blinding of outcome assessors (including study subjects if outcomes were self-reported), including no blinding or incomplete blinding. |
| **Selective reporting of results** | Were all measured outcomes reported? |
| Definitely low risk of bias | There is direct evidence that all of the study’s measured outcomes (primary and secondary) outlined in the protocol methods, abstract, and/or introduction (that are relevant for the evaluation) have been reported. This would include outcomes reported with sufficient detail to be included in meta-analysis or fully tabulated during data extraction and analyses had been planned in advance. |
| Probably low risk of bias | There is indirect evidence that all of the study’s measured outcomes (primary and secondary) outlined in the protocol, methods, abstract, and/or introduction (that are relevant for the evaluation) have been reported, OR analyses that had not been planned in advance (i.e., retrospective unplanned subgroup analyses) are clearly indicated as such and it is deemed that the unplanned analyses were appropriate and selective reporting would not appreciably bias results (e.g., appropriate analyses of an unexpected effect). This would include outcomes reported with insufficient detail such as only reporting that results were statistically significant (or not). |
| Probably high risk of bias | There is indirect evidence that all of the study’s measured outcomes (primary and secondary) outlined in the protocol, methods, abstract, and/or introduction (that are relevant for the evaluation) have been reported, OR and there is indirect evidence that unplanned analyses were included that may appreciably bias results, OR there is insufficient information provided about selective outcome reporting (record “NR” as basis for answer) |
| Definitely high risk of bias | There is direct evidence that all of the study’s measured outcomes (primary and secondary) outlined in the protocol, methods, abstract, and/or introduction (that are relevant for the evaluation) have not been reported. In addition to not reporting outcomes, this would include reporting outcomes based on composite score without individual outcome components or outcomes reported using measurements, analysis methods or subsets of the data (e.g., subscales) that were not pre-specified or reporting outcomes not pre-specified, or that unplanned analyses were included that would appreciably bias results. |

#### 2. Approach for Determining Tiers of Study Quality for Individual Observational Studies

| |
|---|
| Tier 1: A study must be rated as "definitely low" or "probably low" risk of bias for key elements AND have most other applicable items answered "definitely low" or "probably low" risk of bias. |
| Tier 2: Study meets neither the criteria for Tier1 or Tier3. |
| Tier 3: A study must be rated as "definitely high" or "probably high" risk of bias for key elements AND have most other applicable items answered "definitely high" or "probably high" risk of bias. |

#### 3. Guidance on When to Downgrade for Risk of Bias Across Studies

| Downgrade | Interpretation | Guidance |
|---|---|---|
| Not likely | Plausible bias unlikely to seriously alter the results | Most information is from Tier 1 studies (low risk of bias for all key domains). |
| Serious | Plausible bias that raises some doubt about the results | Most information is from Tier 1 and 2 studies. |
| Very serious | Plausible bias that seriously weakens confidence in the results. | The proportion of information from Tier 3 studies at high risk of bias for all key domains is sufficient to affect the interpretation of results. |

#### eAppendix 4. Confidence in the body of evidence and the level of evidence Criteria

#### 1. Initial rating of confidence based on study design

**The initial confidence rating will be determined by four key study design features**

| |
|---|
| • The exposure to the substance is experimentally controlled |
| • The exposure assessment demonstrates that exposures occurred prior to the development of the outcome (or concurrent with aggravation/amplification of an existing condition) |
| • The outcome is assessed on the individual level (i.e., not through population aggregate data) |
| • An appropriate comparison group is included in the study |

**Study Design Features for Initial Confidence Rating**

| Study Design | Controlled Exposure | Exposure Prior to Outcome | Individual Outcome Data | Comparison Group Used | Initial Confidence Rating |
|---|---|---|---|---|---|
| Human controlled trial | likely | likely | likely | likely | high |
| Cohort | unlikely | may or may not | likely | likely | low to moderate |
| Case-control | unlikely | may or may not | likely | likely | low to moderate |
| Cross-sectional | unlikely | unlikely | likely | likely | low |
| Ecologic | unlikely | may or may not | may or may not | likely | Very low to moderate |
| Case series/report | unlikely | may or may not | likely | unlikely | Very low to low |

#### 2. Factors that can downgrade confidence

The initial rating is downgraded for five factors that decrease confidence in the results (risk of bias, unexplained inconsistency, indirectness or lack of applicability, imprecision, and publication bias)

##### 2.1 Risk of bias across studies

| | |
|---|---|
| "Not likely" | Most information is from Tier 1 studies (low risk of bias for all key domains).<br>Plausible bias unlikely to seriously alter the results |
| "Serious" | Most information is from Tier 1 and 2 studies.<br>Plausible bias that raises some doubt about the results |
| "Very serious" | The proportion of information from Tier 3 studies at high risk of bias for all key domains is sufficient to affect the interpretation of results.<br>Plausible bias that seriously weakens confidence in the results. |

##### Imprecision

| | |
|---|---|
| Not serious | • No or minimal indications of large standard deviations (i.e., SD > mean)<br>• For ratio measures (e.g., odds ratio, OR) the ratio of the upper to lower 95% |

| | |
|---|---|
| | CI for most studies (or meta-estimate) is < 10; or for absolute measures (e.g., percent control response) the absolute difference between the upper and lower 95% CI for most studies (or meta-estimate) is < 100 |
| Serious | Does not clearly meet guidance for "not serious" or "very serious" |
| Very serious | • Large standard deviations (i.e., SD > mean)<br>• For ratio measures (e.g., OR) the ratio of the upper to lower 95% CI for most studies (or meta-estimate) is ≥ 10; or for absolute measures (e.g., percent control response) the absolute difference between the upper and lower 95% CI for most studies (or meta-estimate) is ≥ 100 |

#### 2.2 Unexplained Inconsistency

| | |
|---|---|
| "Not serious" | • Point estimates similar<br>• Confidence intervals overlap<br>• Statistical heterogeneity is non-significant<br>• $I^2$ of ≤ 50% |
| "Serious" | • Point estimates vary<br>• Confidence intervals show minimal overlap<br>• Statistical heterogeneity has low p-value (p ≤ 0.1)<br>• I2 of > 50% to 75% |
| "Very serious" | • Point estimates vary widely<br>• Confidence intervals show minimal or no overlap<br>• Statistical heterogeneity has low p-value (p ≤ 0.1)<br>• I2 of > 75% |

#### 2.3 Publication bias

| |
|---|
| • Early positive studies, particularly if small in size, are suspect. |
| • Publication bias should be suspected when studies are uniformly small, particularly when sponsored by industries, non-government organizations, or authors with conflicts of interest. |
| • Funnel plots, Egger's regression, and trim and fill techniques can be used to visualize asymmetrical or symmetrical patterns of study results to help assess publication bias when adequate data for a specific outcome are available. Funnel plots and other approaches are less reliable when there are only a few studies. |
| • The identification of abstracts or other types of grey literature that do not appear as full-length articles within a reasonable time frame (around 3 to 4 years) can be another indication of publication bias. |

#### 2.4 Indirectness or lack of applicability

OHAT considers downgrading confidence for indirectness by evaluating: (1) relevance of the animal model to outcome of concern, (2) directness of the endpoints to the primary health outcome(s), (3) nature of the exposure in human studies and route of administration in animal studies, and (4) duration of treatment in animal studies and length of time between exposure and outcome assessment in animal and prospective human studies.

| | |
|---|---|
| relevance of the animal model to outcome of concern | Studies conducted in mammalian model systems are assumed relevant for humans (i.e., not downgraded) unless compelling evidence to the contrary |

| | |
|---|---|
| | is identified during the course of the evaluation. |
| endpoint directness for the health outcome(s) | The applicability of specific health outcomes or biological processes in non-human animal models is outlined in the PECO-based inclusion and exclusion criteria, with the most accepted relevant/interpretable outcomes considered "primary" and less direct measures, biomarkers of effect, or upstream measures of health outcome considered "secondary." |
| alignment of human exposure | • Human studies are not downgraded for directness regardless of the exposure level or setting (e.g., general population, occupational settings, etc.).<br>• Dose levels used in animal studies: There is no downgrading for dose level used in experimental animal studies because it is not considered as a factor under directness for the purposes of reaching confidence ratings for evidence of health effects. OHAT recognizes that the level of dose or exposure is an important factor when considering the relevance of study findings. In OHAT's process, consideration of dose occurs after hazard identification as part of reaching a "level of concern" conclusion when the health effect is interpreted in the context of what is known regarding the extent and nature of human exposure.<br>• Route of administration in animal studies: External dose comparisons used to reach level of concern conclusions need to consider internal dosimetry in animal models, which can vary based on route of administration, species, age, diet, and other cofactors. The most commonly used routes of administration (i.e., oral, dermal, inhalation, subcutaneous) are generally considered direct for the purposes of establishing confidence ratings. Pharmacokinetic data are also considered. Other routes of administration are more likely to be considered indirect (e.g., intraperitoneal, water for aquatic species, or culture media for culture media for cells, ex vivo preparations, or invertebrates). |
| adequacy of treatment duration and exposure-outcome timing in animal and prospective human studies. | Studies that assess health outcomes following longer periods of exposure and follow-up are generally anticipated to be more informative than studies of shorter duration, e.g., acute toxicity studies lasting from hours to several days. |

#### 2.5 Unexplained Inconsistency

| | |
|---|---|
| "Not serious" | • Point estimates similar<br>• Confidence intervals overlap<br>• Statistical heterogeneity is non-significant<br>• $I^2$ of ≤ 50% |
| "Serious" | • Point estimates vary<br>• Confidence intervals show minimal overlap<br>• Statistical heterogeneity has low p-value (p ≤ 0.1)<br>• I2 of > 50% to 75% |

| | |
|---|---|
| "Very serious" | • Point estimates vary widely<br>• Confidence intervals show minimal or no overlap<br>• Statistical heterogeneity has low p-value (p ≤ 0.1)<br>• I2 of > 75% |

##### 3. Factors that can upgrade confidence

Four properties for a body of evidence (large magnitude of effect, dose response, plausible confounding that would have an impact on the observed association, and consistency across study designs and experimental model systems) are used to determine if the initial confidence rating should be upgraded.

| | |
|---|---|
| **Large magnitude of effect** | when effects might be considered "large" in human studies based primarily on modeling studies that suggest confounding alone is unlikely to explain associations with a relative risk (RR) greater than 2 (or less than 0.5) and very unlikely to explain associations with an RR greater than 5 (or less than 0.2) |
| **Dose Response** | OHAT will upgrade for evidence of a monotonic dose-response gradient (Guyatt et al. 2011g) and for evidence of a non-monotonic dose response when data fit the expected pattern, i.e., prior knowledge leads to expectation for non-monotonic dose response, and/or non-monotonic dose response is consistently observed in the evidence base. |
| **Plausible confounding** | Sources of potential plausible confounding, also known as "residual confounding" or "residual bias" in epidemiology need to be investigated specially among the human body of evidence based with observational studies. |
| **Consistency** | the consistency across animal studies, dissimilar populations and study types. |

##### 4. Final rate of confidence

The overall confidence rating is determined by evaluating all downgrading and upgrading factors in relation to the initial assessment. The possible final ratings for the body of evidence are high, moderate, or low confidence.

| Step | Criteria | Description |
|---|---|---|
| **Step1**<br>**Initial Confidence by Key Features of Study Design** | High (++++) | 4 Features |
| | Moderate (+++) | 3 Features |
| | Low (++) | 2 Features |
| | Very Low (+) | ≤1 Feature |
| **Step2**<br>**Factors Decreasing Confidence** | Risk of Bias | |
| | Unexplained Inconsistency | |
| | Indirectness | |
| | Imprecision | |

| | | |
|---|---|---|
| | Publication Bias | |
| **Step3**<br>**Factors Increasing Confidence** | Large Magnitude of Effect | |
| | Dose Response | |
| | Residual Confounding | • Studies report an effect and residual confounding is toward null<br>• Studies report no effect and residual confounding is away from null |
| | Consistency | • Across animal models or species<br>• Across dissimilar populations<br>• Across study design types |
| | Other | e.g., particularly rare outcomes |
| **Step4**<br>**Confidence in the Body of Evidence** | High (++++) | |
| | Moderate (+++) | |
| | Low (++) | |

#### eAppendix 5. Detailed risk of bias results of the included studies.

***Risk of bias of VoPham et al.2022***, according to instructions reported in Appendix 3:

| Bias Domain | Risk of Bias | Comments |
|---|---|---|
| **CONFOUNDING BIAS. ***<br>Did the study design or analysis account for important confounding and modifying variables? | Probably low | The study adjusted for age, sex, ethnicity, region, year, primary payer, ZIP Code-level median household income, urbanicity, obesity, diabetes, metabolic syndrome, impaired fasting glucose, dyslipidemia, hypertension, obstructive sleep apnea, smoking. Although BMI and alcohol consumption were not directly adjusted for, the study controlled for a wide range of variables associated with metabolic health and socioeconomic status. Therefore, the confounding bias is probably low. |
| **ATTRITION/<br>EXCLUSION BIAS**<br>Were outcome data complete without attrition or exclusion from analysis? | Probably high | The study excluded participants with missing data on key demographic variables (e.g., age, sex, region, income). Although the final models adjusted for these confounders, the authors did not report how many observations were excluded or whether the excluded participants differed systematically from those included. Therefore, the risk of attrition/exclusion bias is considered probably high. |
| **DETECTION BIAS**<br>Can we be confident in the exposure characterization? * | Probably low | The study used PM2.5 data from the U.S. Environmental Protection Agency's Air Quality System, a validated and widely used monitoring network. Annual average PM2.5 concentrations were linked to patient ZIP codes, which is a standard approach in population-based epidemiological research. Although individual-level exposure is not available, the use of region-level ambient data is acceptable given the large-scale design of the study and the recognized lack of a gold standard for air pollution exposure assessment. Therefore, the risk of exposure detection bias is considered probably low. |
| Can we be confident in the outcome assessment? * | Probably low | The diagnosis of NAFLD was identified using the ICD-9 code 571.8 from the Nationwide Inpatient Sample (NIS) database. Although not a gold standard like liver biopsy or imaging, ICD coding is widely used in epidemiologic studies and provides standardized, objective criteria across a large patient population. Therefore, the risk of outcome detection bias is considered probably low. |
| **SELECTIVE REPORTING BIAS**<br>Were all measured outcomes reported? | Probably low | The study clearly defined NAFLD as the primary outcome and reported all relevant associations between PM2.5 exposure and NAFLD, including adjusted and unadjusted models. Subgroup and sensitivity analyses were also presented, which supports the completeness and transparency of outcome reporting. There is no indication that outcomes were selectively withheld or selectively emphasized to support a hypothesis. Therefore, the risk of selective reporting bias is considered probably low. |
| **SELECTION BIAS**<br>Did selection of study participants result in appropriate comparison groups? | Probably low | The study used theNIS database which is a nationally representative inpatient database, and applied consistent inclusion/exclusion criteria. However, information on participation rates and comparability of included versus excluded participants was not explicitly reported. Therefore, the risk of selection bias is considered probably low. |
| **CONFLICT OF INTEREST** | Definitely low | "The authors declare they have no actual or potential competing financial interests." |

***Risk of bias of Guo et al.2022,*** according to instructions reported in Appendix 3:

| Bias Domain | Risk of Bias | Comments |
|---|---|---|
| **CONFOUNDING BIAS. *** Did the study design or analysis account for important confounding and modifying variables? | Probably low | The study adjusted for age, sex, ethnicity, region, education, income, alcohol use, smoking, high-fat diet, low fruit and vegetable intake, low physical activity, second-hand smoke, and indoor air pollution. Although BMI or obesity status was not directly adjusted for, the study controlled for a wide range of variables associated with metabolic health and socioeconomic status. Therefore, the confounding bias is probably low. |
| **ATTRITION/ EXCLUSION BIAS** Were outcome data complete without attrition or exclusion from analysis? | Probably low | The study excluded participants with missing key variables or diagnostic data using a transparent complete-case approach, which likely minimized bias in this large population-based sample. As this was a cross-sectional study without follow-up, attrition over time is not applicable. Therefore, the risk of attrition/exclusion bias is considered probably low. |
| **DETECTION BIAS** Can we be confident in the exposure characterization? * | Probably low | Air pollution exposure was estimated using a satellite-based spatiotemporal model with high spatial resolution, which, although indirect, is widely accepted in environmental epidemiology and provides reliable exposure estimates. Therefore, the risk of exposure detection bias is considered probably low. |
| Can we be confident in the outcome assessment? * | Definitely low | The diagnosis of MAFLD was based on liver ultrasound imaging and metabolic criteria, which, although not the gold standard, are widely used and considered valid in large-scale population studies. Therefore, the risk of outcome detection bias is considered definitely low. |
| **SELECTIVE REPORTING BIAS** Were all measured outcomes reported? | Probably low | The study clearly reported all prespecified outcomes related to MAFLD and air pollution exposure. The analyses were consistent with the stated objectives, and no evidence of selective omission was identified. Therefore, the risk of selective reporting bias is considered probably low. |
| **SELECTION BIAS** Did selection of study participants result in appropriate comparison groups? | Probably low | The study used a large population-based cohort (CMEC) with stratified cluster sampling, and applied clear inclusion and exclusion criteria. Although the study was limited to southwestern China, the design allowed for appropriate internal comparisons between exposed and unexposed groups. Therefore, the risk of selection bias is considered probably low. |
| **CONFLICT OF INTEREST** | Definitely low | "The authors declare the funders had no role in the study design, data collection, data analysis and interpretation, writing of the report or decision to submit the article for publication." |

***Risk of bias of Ji et al.2024,*** according to instructions reported in Appendix 3:

| **Bias Domain** | **Risk of Bias** | **Comments** |
|---|---|---|
| **CONFOUNDING BIAS. *** Did the study design or analysis account for important confounding and modifying variables? | Probably low | The study adjusted for age, sex, ethnicity, education, exercise frequency, smoking status, kitchen exhaust facilities, and solid fuel use. While BMI and alcohol consumption were not explicitly included in the final adjusted model. Therefore, the risk of confounding bias is considered probably low. |
| **ATTRITION/ EXCLUSION BIAS** Were outcome data complete without attrition or exclusion from analysis? | Probably low risk | The study excluded participants with missing or extreme values using defined criteria, and still retained over 2.7 million subjects. As this was a cross-sectional study without follow-up, attrition over time is not applicable. Therefore, the risk of attrition/exclusion bias is considered probably low.so the risk of attrition bias is probably low. |
| **DETECTION BIAS** Can we be confident in the exposure characterization? * | Probably low | Exposure to $PM_1$, $PM_{2.5}$, $PM_{10}$, and $NO_2$ was estimated using satellite-based random forest models, GAM, and traditional regression at ~10 km resolution. The models showed acceptable predictive power (CV $R^2$ of 64%–83%) and exposure was assigned based on geocoded residential addresses. While this approach is scientifically robust and widely used in large cohort studies, the resolution is not fine enough to rule out all potential misclassification at the individual level. Therefore, the risk of detection bias is probably low. |
| Can we be confident in the outcome assessment? * | Definitely low | The diagnosis of MAFLD was based on liver ultrasound and metabolic criteria, although not the gold standard, are widely used and considered valid in large-scale population studies. Therefore, the risk of outcome detection bias is considered definitely low. |
| **SELECTIVE REPORTING BIAS** Were all measured outcomes reported? | Probably low | The study clearly reported all prespecified outcomes related to MAFLD and air pollution exposure. The analyses were consistent with the stated objectives, and no evidence of selective omission was identified. Although there was no mention of a preregistered protocol. Therefore, the risk of selective reporting bias is considered probably low. |
| **SELECTION BIAS** Did selection of study participants result in appropriate comparison groups? | Probably low | The study applied clear and comprehensive exclusion criteria to ensure appropriate comparison groups and reduce the risk of confounding by extreme values or pre-existing liver conditions. Therefore, the risk of selection bias is considered probably low. |
| **CONFLICT OF INTEREST** | Definitely low | "The authors declare that they have no known competing financial interests or personal relationships that could have appeared to influence the work reported in this paper." |

***Risk of bias of Han et al.2023,*** according to instructions reported in Appendix 3:

| **Bias Domain** | **Risk of Bias** | **Comments** |
|---|---|---|
| **CONFOUNDING BIAS. *** Did the study design or analysis account for important confounding and modifying variables? | Definitely low | The study adjusted for age, sex, ethnicity, education, income, residency, BMI, alcohol and tea consumption, smoking status, secondary smoke, Mediterranean dietary, and Mediterranean diet adherence, temperature and relative humidity, indoor pollution. These variables cover key demographic, lifestyle, and dietary factors that are commonly associated with both exposure |

| | | |
|---|---|---|
| | | and outcome. Therefore, the risk of confounding bias is considered definitely low. |
| **ATTRITION/ EXCLUSION BIAS** Were outcome data complete without attrition or exclusion from analysis? | Definitely low | The study clearly documented the exclusion criteria and transparently reported the number of participants excluded due to missing follow-up data or lack of outcome information. These exclusions were transparent, based on objective criteria. Therefore, the risk of attrition/exclusion bias is considered definitely low. |
| **DETECTION BIAS** Can we be confident in the exposure characterization? * | Probably low | Air pollution exposure was estimated using the China High Air Pollutants (CHAP) dataset, with high spatial resolution (1 km × 1 km for PM and 10 km × 10 km for gaseous pollutants), and exposure was assigned based on participants' residential addresses over multiple years to reflect long-term exposure. Although indirect, this method is widely accepted and minimizes exposure misclassification. Therefore, the risk of exposure detection bias is probably low. |
| Can we be confident in the outcome assessment? * | Definitely low | The diagnosis of MAFLD was based on liver ultrasound imaging and metabolic criteria, which, although not the gold standard, are widely used and considered valid in large-scale population studies. Therefore, the risk of outcome detection bias is considered definitely low. |
| **SELECTIVE REPORTING BIAS** Were all measured outcomes reported? | Probably low | The study clearly reported all prespecified outcomes related to MAFLD and air pollution exposure. The analyses were consistent with the stated objectives, and no evidence of selective omission was identified. Although there was no mention of a preregistered protocol. Therefore, the risk of selective reporting bias is considered probably low. |
| **SELECTION BIAS** Did selection of study participants result in appropriate comparison groups? | Probably low | The study applied well-defined inclusion and exclusion criteria to ensure appropriate comparison groups, minimizing confounding due to pre-existing conditions and misclassification of exposure or outcome timing. |
| **CONFLICT OF INTEREST** | Definitely low | "The authors declare there is no conflict of interest to disclose." |

***Risk of bias of Bo et al.2024,*** according to instructions reported in Appendix 3:

| **Bias Domain** | **Risk of Bias** | **Comments** |
|---|---|---|
| **CONFOUNDING BIAS. *** Did the study design or analysis account for important confounding and modifying variables? | Definitely low | The study adjusted for age, sex, educational level, BMI, hypertension, diabetes, dyslipidemia, smoking, alcohol drinking, physical activity, fruit intake, vegetable intake, occupational exposure to dust/organic solvents and season. These variables cover key demographic, lifestyle, and dietary factors that are commonly associated with both exposure and outcome. Therefore, the risk of confounding bias is considered definitely low. |
| **ATTRITION/ EXCLUSION BIAS** Were outcome data complete without attrition or exclusion from analysis? | Probably low | The study clearly documented the exclusion criteria and transparently reported the number of participants excluded due to missing follow-up data or lack of outcome information. These exclusions were transparent, based on objective criteria. As this was a cross-sectional study without follow-up, attrition over time is not applicable. Therefore, the risk of attrition/exclusion bias is |

| | | |
|---|---|---|
| | | considered probably low. |
| **DETECTION BIAS**<br>Can we be confident in the exposure characterization? * | Probably low | Air pollution exposure was estimated using the China High Air Pollutants (CHAP) dataset, with high spatial resolution (1 km × 1 km for PM and 10 km × 10 km for gaseous pollutants), and exposure was assigned based on participants' residential addresses over multiple years to reflect long-term exposure. Although indirect, this method is widely accepted and minimizes exposure misclassification. Therefore, the risk of exposure detection bias is probably low. |
| Can we be confident in the outcome assessment? * | Probably low | The diagnosis of NAFLD was based on HSI/FLI, which, although not the gold standard, are widely used and considered valid in large-scale population studies. Therefore, the risk of outcome detection bias is considered probably low. |
| **SELECTIVE REPORTING BIAS**<br>Were all measured outcomes reported? | Probably low | The study clearly reported all prespecified outcomes related to MAFLD and air pollution exposure. The analyses were consistent with the stated objectives, and no evidence of selective omission was identified. Although there was no mention of a preregistered protocol. Therefore, the risk of selective reporting bias is considered probably low |
| **SELECTION BIAS**<br>Did selection of study participants result in appropriate comparison groups? | Probably low | The study applied well-defined inclusion and exclusion criteria to ensure appropriate comparison groups, minimizing confounding due to pre-existing conditions and misclassification of exposure or outcome timing. Therefore, the risk of selection bias is considered probably low. |
| **CONFLICT OF INTEREST** | Definitely low | "The authors declare there is no conflict of interest to disclose." |

***Risk of bias of Guo et al.2023,*** according to instructions reported in Appendix 3:

| **Bias Domain** | **Risk of Bias** | **Comments** |
|---|---|---|
| **CONFOUNDING BIAS. ***<br>Did the study design or analysis account for important confounding and modifying variables? | Probably low | The study adjusted for age, sex, ethnicity, admission sites, urban area, education level, annual household income, alcohol consumption, smoke status, high-fat diet, low fruit and vegetable intake, physical activity, second-hand smoking exposure, biomass fuel exposure, 3-year average temperature and 3-year average humidity. Although major metabolic indicators such as BMI, diabetes, and hypertension were not included, the model accounted for a broad range of relevant lifestyle and demographic factors, suggesting a probably low risk of confounding bias. |
| **ATTRITION/ EXCLUSION BIAS**<br>Were outcome data complete without attrition or exclusion from analysis? | Probably low risk | The study excluded 4.29% of the eligible population due to liver cirrhosis (n=31) or missing data on key covariates and potential modifiers (n=3443). These exclusions were clearly described and represent a relatively small proportion of the total sample (n=80,201). The final sample size remained large (n=76,727), and exclusions did not appear to be systematically related to exposure or outcome. As this was a cross-sectional study without follow-up, attrition over time is not applicable. Therefore, the risk of attrition/exclusion bias is considered probably low. |
| **DETECTION BIAS** | Probably | $PM_{2.5}$ and its constituents were estimated using the V4.CH.02 product |

| | | |
|---|---|---|
| Can we be confident in the exposure characterization? * | low | developed by the Dalhousie University Atmospheric Composition Analysis Group. This dataset integrates satellite data and ground-based calibration, providing modeled concentrations at a 10 km × 10 km resolution. Exposure was assigned based on participants' geocoded residential addresses using a 3-year average prior to the baseline survey. Although indirect, this method is widely used in air pollution epidemiology and helps reduce exposure misclassification, supporting a probably low risk of detection bias. |
| Can we be confident in the outcome assessment? * | Definitely low | The diagnosis of MAFLD was based on liver ultrasound imaging and metabolic criteria, which, although not the gold standard, are widely used and considered valid in large-scale population studies. Therefore, the risk of outcome detection bias is considered definitely low. |
| **SELECTIVE REPORTING BIAS**<br>Were all measured outcomes reported? | Probably low | All key outcomes relevant to air pollution and MAFLD were reported. The analyses were consistent with the stated objectives, and no evidence of selective reporting was identified, although there was no mention of a preregistered protocol, there is no indication that important outcomes were selectively omitted. Therefore, the risk of selective reporting bias is considered probably low. |
| **SELECTION BIAS**<br>Did selection of study participants result in appropriate comparison groups? | Probably low | The study applied well-defined inclusion and exclusion criteria to ensure appropriate comparison groups, minimizing confounding due to pre-existing conditions and misclassification of exposure or outcome timing. Therefore, the risk of selection bias is considered probably low. |
| **CONFLICT OF INTEREST** | Definitely low | "The authors declare there is no conflict of interest to disclose." |

***Risk of bias of Matthiessen et al.2023,*** according to instructions reported in Appendix 3:

| **Bias Domain** | **Risk of Bias** | **Comments** |
|---|---|---|
| **CONFOUNDING BIAS. ***<br>Did the study design or analysis account for important confounding and modifying variables? | Probably low | The final models adjusted for several key confounders, including age, sex, individual and neighborhood socioeconomic status, diet, physical activity, smoking status, alcohol consumption, and pack-years, environmental tobacco smoke. Although BMI was not included, most major lifestyle and behavioral risk factors were considered, suggesting a probably low risk of confounding bias. |
| **ATTRITION/ EXCLUSION BIAS**<br>Were outcome data complete without attrition or exclusion from analysis? | Probably low | The study began with 4,814 participants, and 749 were excluded due to missing exposure, outcome, or covariate data. Although the excluded individuals differed from the included population (e.g., more men, more smokers, and less physically active), the proportion of excluded participants was moderate, and reasons for exclusion were clearly reported. The characteristics of the included and excluded groups were described and compared, reducing the likelihood of significant bias. As this was a cross-sectional study without follow-up, attrition over time is not applicable. Therefore, the risk of attrition/exclusion bias is considered probably low. |
| **DETECTION BIAS**<br>Can we be confident in the | Probably low | The study employed two well-validated models to assess exposure to air pollution: the EURAD-CTM (chemistry transport model) and the ESCAPE-LUR |

| | | |
|---|---|---|
| exposure characterization? * | | (land use regression) model. Both models are widely accepted, region-specific, and follow standardized methodologies. The EURAD-CTM provided daily concentrations of $PM_{10}$, $PM_{2.5}$, $NO_2$, and particle number at a 1 km² resolution based on residential addresses, using detailed input variables such as transport and industry emissions. The ESCAPE-LUR model used direct pollutant measurements from 20–40 monitoring sites and multiple predictor variables to estimate long-term exposure levels. Both models assessed multi-year averages (2001–2003 for CTM and 2008–2009 for LUR), improving the reliability of long-term exposure estimation. Therefore, the risk of exposure detection bias is probably low. |
| Can we be confident in the outcome assessment? * | Probably low | The outcome was assessed using the Fatty Liver Index (FLI), a validated surrogate marker based on BMI, waist circumference, triglycerides, and GGT. The FLI has been developed through logistic regression in the general population and has been widely used in epidemiological studies. Although it does not include imaging or histological confirmation, the method is transparent, systematic, and independent of exposure status. The same algorithm was applied consistently across participants, minimizing the risk of differential outcome assessment bias. Therefore, the risk of outcome detection bias is probably low. |
| **SELECTIVE REPORTING BIAS**<br>Were all measured outcomes reported? | Probably low | All prespecified outcomes described in the methods were reported in the results. Although no protocol was available, there was no indication of selective reporting. Although there was no mention of a preregistered protocol, there is no indication that important outcomes were selectively omitted. Therefore, the risk of selective reporting bias is considered probably low. |
| **SELECTION BIAS**<br>Did selection of study participants result in appropriate comparison groups? | Probably low | The study applied well-defined inclusion and exclusion criteria to ensure appropriate comparison groups, minimizing confounding due to pre-existing conditions and misclassification of exposure or outcome timing. Therefore, the risk of selective reporting bias is considered probably low. |
| **CONFLICT OF INTEREST** | Definitely low | "The authors declare that they have no conflicts of interest with regard to the content of this report." |

***Risk of bias of Cheng et al.2024,*** according to instructions reported in Appendix 3:

| **Bias Domain** | **Risk of Bias** | **Comments** |
|---|---|---|
| **CONFOUNDING BIAS. ***<br>Did the study design or analysis account for important confounding and modifying variables? | Probably low | The study adjusted for multiple key confounders, including age, sex, marital status, education, income, alcohol consumption, smoking, dietary habits (fried food, vegetables, fruit, sugary drinks), physical activity, and enrollment year. Although BMI was not included, adjustment for alcohol, smoking, and lifestyle factors reduces residual confounding. Therefore the risk of confounding bias is considered probably low. |
| **ATTRITION/**<br>**EXCLUSION BIAS**<br>Were outcome data complete without attrition | Probably low risk | The study clearly documented the reasons for exclusion and the number of participants excluded at each step, including missing exposure or outcome data, underage participants, and specific medical histories. The criteria were pre-defined, consistently applied, and transparently reported. The final sample |

| | | |
|---|---|---|
| or exclusion from analysis? | | size remained large, reducing the likelihood of significant bias due to attrition. As this was a cross-sectional study without follow-up, attrition over time is not applicable. Therefore, the risk of attrition/exclusion bias is considered probably low. |
| **DETECTION BIAS** Can we be confident in the exposure characterization? * | Probably low | Air pollution exposure was assessed using validated land-use regression models combined with machine learning, with a fine spatial resolution (50 m × 50 m grid). Exposure estimation was based on geocoded residential addresses and calculated as 3-year averages prior to health checkups. These methodological strengths reduce the likelihood of serious exposure detection bias. Therefore, the risk of exposure detection bias is probably low. |
| Can we be confident in the outcome assessment? * | Definitely low | The diagnosis of MAFLD was based on liver ultrasound imaging and metabolic criteria, which, although not the gold standard, are widely used and considered valid in large-scale population studies. Therefore, the risk of outcome detection bias is considered definitely low. |
| **SELECTIVE REPORTING BIAS** Were all measured outcomes reported? | Probably low | All prespecified outcomes described in the methods were reported in the results. Although there was no mention of a preregistered protocol, there is no indication that important outcomes were selectively omitted. Therefore, the risk of selective reporting bias is considered probably low. |
| **SELECTION BIAS** Did selection of study participants result in appropriate comparison groups? | Probably low | The study applied well-defined inclusion and exclusion criteria to ensure appropriate comparison groups, minimizing confounding due to pre-existing conditions and misclassification of exposure or outcome timing. Therefore, the risk of selective reporting bias is considered probably low. |
| **CONFLICT OF INTEREST** | Definitely low | "The authors declare no known competing financial interests or personal relationships that could have appeared to influence the work reported in this paper." |

***Risk of bias of Patterson et al.2024***, according to instructions reported in Appendix 3:

| Bias Domain | Risk of Bias | Comments |
|---|---|---|
| **CONFOUNDING BIAS. *** Did the study design or analysis account for important confounding and modifying variables? | Probably low | The study adjusted for age, sex, ethnicity, BMI, cigarette smoking, e-cigarette use, exercise, parental education, and occupation status with town code as a random effect. the risk of confounding bias is considered definitely low. Although alcohol consumption was not included, adjustment for other key factors reduces residual confounding. Therefore the risk of confounding bias is considered probably low. |
| **ATTRITION/ EXCLUSION BIAS** Were outcome data complete without attrition or exclusion from analysis? | Probably low risk | Of the 172 enrolled participants, only 2 (1.2%) were excluded due to missing long-term $NO_2$ exposure data. The final analysis included 124 participants with complete exposure data. Although 46 participants had incomplete data, the comparison of baseline characteristics indicated that participants with complete data were only slightly younger, with no significant differences in other key variables. These details suggest limited attrition-related bias, supporting a probably low risk rating. |
| **DETECTION BIAS** Can we be confident in the | Probably low | Air pollution exposure was assessed using data from U.S. EPA monitoring stations employing Federal Reference Method (FRM) monitors for $NO_2$ and $O_3$ |

| | | |
|---|---|---|
| exposure characterization? * | | and Federal Equivalent Method (FEM) monitors for $PM_{2.5}$ and $PM_{10}$. Monthly average concentrations were interpolated from up to four stations within 50 km using inverse-distance squared weighting. Although individual-level exposure misclassification is possible, the use of standardized and validated federal monitoring methods with spatial weighting reduces the likelihood of substantial exposure bias. Therefore, the risk of exposure detection bias is probably low. |
| Can we be confident in the outcome assessment? * | Definitely low | NAFLD was assessed using 3-Tesla whole-abdominal magnetic resonance imaging (MRI) with ≥5.5% hepatic fat fraction (HFF) as the diagnostic criterion. Although liver biopsy is considered the gold standard, it is not feasible in population studies. MRI is considered the gold standard for non-invasive hepatic fat quantification, reducing the likelihood of outcome misclassification. Therefore, the risk of outcome detection bias is definitely low. |
| **SELECTIVE REPORTING BIAS** Were all measured outcomes reported? | Probably low | All prespecified outcomes described in the methods were reported in the results. Although there was no mention of a preregistered protocol, there is no indication that important outcomes were selectively omitted. Therefore, the risk of selective reporting bias is considered probably low. |
| **SELECTION BIAS** Did selection of study participants result in appropriate comparison groups? | Probably low | The study applied well-defined inclusion and exclusion criteria to ensure appropriate comparison groups, minimizing confounding due to pre-existing conditions and misclassification of exposure or outcome timing. Therefore, the risk of selective reporting bias is considered probably low. |
| **CONFLICT OF INTEREST** | Definitely low | "The authors declare that they have no known competing financial interests or personal relationships that could have appeared to influence the work reported in this paper." |

##### *Risk of bias of Kim et al.2019, according to instructions reported in Appendix 3:*

| **Bias Domain** | **Risk of Bias** | **Comments** |
|---|---|---|
| **CONFOUNDING BIAS. *** Did the study design or analysis account for important confounding and modifying variables? | Probably low | The study adjusted for several important covariates including age, sex, education level, marital status, hypertension, smoking status, urine creatinine, total cholesterol, and NHANES cycle. However, it did not adjust for BMI and alcohol consumption, major determinants of NAFLD, which may result in residual confounding. Therefore, the risk of bias due to confounding is considered probably low. |
| **ATTRITION/ EXCLUSION BIAS** Were outcome data complete without attrition or exclusion from analysis? | Probably low risk | Although 1,133 participants were excluded due to missing key data (e.g., BMI, liver enzyme levels, alcohol intake, viral hepatitis status, pregnancy), the exclusions were clearly defined and systematically applied. The proportion of excluded subjects (13%) is moderate, and the reasons were not likely related to outcomes. As this was a cross-sectional study without follow-up, attrition over time is not applicable. Therefore, the risk of attrition/exclusion bias is considered probably low. |
| **DETECTION BIAS** Can we be confident in the | Definitely low | The study measured urinary BPA using solid-phase extraction combined with high-performance liquid chromatography and tandem mass spectrometry |

| | | |
|---|---|---|
| exposure characterization? * | | (HPLC-MS/MS), which is a highly sensitive and specific analytical method. The use of isotopically labelled internal standards and a comprehensive quality control system further ensured the accuracy and reliability of BPA detection. The detection limits (0.1–2.3 ng/mL) were appropriate for detecting BPA in the general population. These details confirm that BPA exposure was assessed using validated, accurate, and reproducible methods, supporting a rating of definitely low risk of detection bias. |
| Can we be confident in the outcome assessment? * | Prabably low | The study used two non-invasive surrogate indices to define NAFLD: the Hepatic Steatosis Index (HSI) and the US Fatty Liver Index (USFLI), both of which were calculated based on biochemical and anthropometric parameters. These indices have been externally validated in epidemiological studies. However, neither method directly measures liver fat through imaging or histology. Therefore, while the outcome definition is standardized and based on validated formulas, the indirect nature and limited sensitivity of these indices introduce some uncertainty, supporting a rating of probably low risk of bias. |
| **SELECTIVE REPORTING BIAS** Were all measured outcomes reported? | Probably low | All prespecified outcomes described in the methods were reported in the results. Although no protocol was available, there was no indication of selective reporting. Although there was no mention of a preregistered protocol, there is no indication that important outcomes were selectively omitted. Therefore, the risk of selective reporting bias is considered probably low. |
| **SELECTION BIAS** Did selection of study participants result in appropriate comparison groups? | Probably low | The study applied well-defined inclusion and exclusion criteria to ensure appropriate comparison groups, minimizing confounding due to pre-existing conditions and misclassification of exposure or outcome timing. Therefore, the risk of selective reporting bias is considered probaly low. |
| **CONFLICT OF INTEREST** | Definitely low | The authors explicitly declared no conflicts of interest (“Nothing to disclose”). |

***Risk of bias of Peng et al.2023,*** according to instructions reported in Appendix 3:

| **Bias Domain** | **Risk of Bias** | **Comments** |
|---|---|---|
| **CONFOUNDING BIAS. *** Did the study design or analysis account for important confounding and modifying variables? | Probably low | The study adjusted for multiple potential confounders, including ethnicity, education, hypertension, diabetes, drinking, log-transformed levels of triglycerides, high-density lipoprotein cholesterol, glucose, glycosylated hemoglobin A1c, urine creatinine (tertiles), and the log-transformed concentration of BPA. However, key variables such as age, sex, and BMI were not included in the adjustment. Therefore, the risk of confounding bias is considered probably low. |
| **ATTRITION/ EXCLUSION BIAS** Were outcome data complete without attrition or exclusion from analysis? | Probably low risk | The study clearly described its exclusion criteria, including the removal of participants with missing key demographic and biochemical variables. This transparent approach helps reduce bias introduced by incomplete data. Additionally, the analysis was conducted using a nationally representative dataset (NHANES), which is known for its high data quality. However, since the study employed a cross-sectional design without any follow-up, there was no opportunity to assess longitudinal attrition. While the risk of attrition bias |

| | | |
|---|---|---|
| | | appears limited, the absence of follow-up data supports a rating of probably low risk of bias. |
| **DETECTION BIAS**<br>Can we be confident in the exposure characterization? * | Definitely low | Urinary concentrations of BPA, BPS, and BPF were measured using a validated method: solid-phase extraction with high-performance liquid chromatography and tandem mass spectrometry (HPLC-MS/MS), performed by the CDC. This standardized approach ensures reliable and consistent exposure assessment. Urinary creatinine was also used to correct for dilution. Therefore, the risk of outcome detection bias is definitly low. |
| Can we be confident in the outcome assessment? * | Prabably low | The study defined NAFLD using the hepatic steatosis index (HSI), a widely used non-invasive algorithm based on ALT, AST, BMI, sex, and diabetes status. Although HSI is not the gold standard, it has been validated in population-based studies and offers a practical alternative to biopsy. Given the standardized formula and prior validation, the risk of outcome misclassification is limited, supporting a probably low rating. |
| **SELECTIVE REPORTING BIAS**<br>Were all measured outcomes reported? | Probably low | All prespecified outcomes described in the methods were reported in the results. Although no protocol was available, there was no indication of selective reporting. Although there was no mention of a preregistered protocol, there is no indication that important outcomes were selectively omitted. Therefore, the risk of selective reporting bias is considered probably low. |
| **SELECTION BIAS**<br>Did selection of study participants result in appropriate comparison groups? | Probably low | The study applied well-defined inclusion and exclusion criteria to ensure appropriate comparison groups, minimizing confounding due to pre-existing conditions and misclassification of exposure or outcome timing. Therefore, the risk of selective reporting bias is considered probably low. |
| **CONFLICT OF INTEREST** | Definitely low | "No conflict of interest with respect to this manuscript to disclose." |

***Risk of bias of An et al.2021,*** according to instructions reported in Appendix 3:

| **Bias Domain** | **Risk of Bias** | **Comments** |
|---|---|---|
| **CONFOUNDING BIAS. ***<br>Did the study design or analysis account for important confounding and modifying variables? | Probably low | The study adjusted for several potential confounders, including age, sex, marital status, education, income, hypertension, diabetes, hyperlipidemia, smoking, drinking, exercise, and creatinine, helping to control for key lifestyle and metabolic factors. However, important variables such as BMI were not included in the adjustment. Therefore, while confounding was partially addressed, the residual risk supports a rating of probably low. |
| **ATTRITION/**<br>**EXCLUSION BIAS**<br>Were outcome data complete without attrition or exclusion from analysis? | Probably low risk | The study clearly described exclusion criteria, such as missing exposure or outcome data, significant alcohol use, pregnancy, hepatitis, and abnormal AST/ALT ratios. Although follow-up data was unavailable due to the cross-sectional design, the transparency and justification of exclusions support a rating of probably low risk of bias. |
| **DETECTION BIAS**<br>Can we be confident in the exposure characterization? | Definitely low | Urinary BPA concentrations were measured using a validated and highly sensitive method (UPLC-MS/MS) by the National Institute of Environmental Research. Sample collection, storage, and analysis followed standardized |

| | | |
|---|---|---|
| * | | procedures, ensuring accurate and consistent exposure assessment. Therefore, the risk of exposure detection bias is definitely low. |
| Can we be confident in the outcome assessment? * | Prabably low | The study used the hepatic steatosis index (HSI), a widely accepted non-invasive biomarker for assessing NAFLD in large population-based studies. The HSI incorporates ALT, AST, BMI, sex, and diabetes status, and a validated cut-off of 36 was applied. While liver biopsy is the gold standard, HSI is widely accepted and practical for large-scale studies. Thus, outcome assessment is considered reasonably accurate, supporting a probably low risk of bias. |
| **SELECTIVE REPORTING BIAS**<br>Were all measured outcomes reported? | Probably low | The study clearly stated its objectives and reported results for the primary associations between urinary BPA and NAFLD, as outlined in the methods. Statistical methods were pre-specified, and outcomes were consistently reported in both the methods and results sections. No evidence suggests that outcomes were selectively omitted or that only significant results were highlighted. |
| **SELECTION BIAS**<br>Did selection of study participants result in appropriate comparison groups? | Probably low | The study utilized data from the Korean National Environmental Health Survey (KoNEHS) III, which employed a multi-stage stratified random sampling method to recruit participants based on age, sex, region, and socioeconomic status. This population-based approach enhances representativeness and reduces selection bias. Although there was no mention of a preregistered protocol, there is no indication that important outcomes were selectively omitted. Therefore, the risk of selective reporting bias is considered probably low. |
| **CONFLICT OF INTEREST** | Definitely low | "The authors declare that they have no competing interests." |

***Risk of bias of He et al.2023***, according to instructions reported in Appendix 3:

| Bias Domain | Risk of Bias | Comments |
|---|---|---|
| **CONFOUNDING BIAS. ***<br>Did the study design or analysis account for important confounding and modifying variables? | Probably low | The study adjusted for multiple potential confounders, including age, sex, ethnicity, marital status, education level, poverty income ratio, hypertension, diabetes, smoking, alcohol use, energy intake, and physical activity. These factors cover many relevant lifestyle and metabolic variables. However, BMI was not included, which is an important confounder for liver outcomes. Therefore, while confounding was partially addressed, the remaining risk supports a rating of probably low. |
| **ATTRITION/<br>EXCLUSION BIAS**<br>Were outcome data complete without attrition or exclusion from analysis? | Probably low | The study clearly described the exclusion criteria, including viral hepatitis, use of steatogenic medications, excessive alcohol consumption, pregnancy, and incomplete elastography data. Although some participants were excluded due to missing or partial data, these exclusions were transparently reported and justified. Because the study was cross-sectional and not longitudinal, follow-up was not applicable. Overall, the clear reporting and rationale support a rating of probably low risk. |
| **DETECTION BIAS**<br>Can we be confident in the exposure characterization? | Definitely low | Urinary phthalate metabolites were measured using a validated and sensitive method—HPLC-ESI-MS/MS (high-performance liquid chromatography-electrospray ionization-tandem mass spectrometry). This method ensures high |

| | | |
|---|---|---|
| * | | specificity and accuracy. Samples were properly collected, stored at −20 °C, and processed by the National Center for Environmental Health, following CDC guidelines. The procedures used are consistent with high-quality standards for biomonitoring, supporting a low risk of bias in exposure assessment. |
| Can we be confident in the outcome assessment? * | Definitely low | NAFLD was assessed using the controlled attenuation parameter (CAP) via FibroScan, a validated and widely accepted imaging-based method for detecting hepatic steatosis. Although liver biopsy is considered the gold standard, it is not feasible in population studies. CAP offers a reliable, non-invasive alternative with high sensitivity and specificity. Given its robust diagnostic performance and standardization in large-scale studies, the risk of outcome misclassification is considered definitely low. |
| **SELECTIVE REPORTING BIAS**<br>Were all measured outcomes reported? | Probably low | The study clearly stated its objectives and reported results for the primary associations between urinary PAEs metabolites and NAFLD, as outlined in the methods. Statistical methods were pre-specified, and outcomes were consistently reported in both the methods and results sections. No evidence suggests that outcomes were selectively omitted or that only significant results were highlighted. Therefore, the risk of selective reporting bias is considered probably low. |
| **SELECTION BIAS**<br>Did selection of study participants result in appropriate comparison groups? | Probably low | Participants were selected from the same large-scale population (NHANES) using a consistent and standardized method. Although baseline characteristics between NAFLD and non-NAFLD groups differed in several aspects (e.g., age, sex, BMI, diabetes, and hypertension), these differences are commonly observed in observational studies and were likely adjusted for in the statistical analysis. Given the indirect evidence of comparable selection processes and the use of standard inclusion/exclusion criteria, the risk of selection bias was rated as probably low. |
| **CONFLICT OF INTEREST** | Definitely low | "The authors declare that they have no known competing financial interests or personal relationships that could have appeared to influence the work reported in this paper." |

***Risk of bias of Yang et al.2021,*** according to instructions reported in Appendix 3:

| **Bias Domain** | **Risk of Bias** | **Comments** |
|---|---|---|
| **CONFOUNDING BIAS. ***<br>Did the study design or analysis account for important confounding and modifying variables? | Probably low | The study adjusted for multiple potential confounders, including age, sex, marital status, education, socioeconomic status, hypertension, diabetes, hyperlipidemia smoking, drinking, exercise and creatinine. These factors cover many relevant lifestyle and metabolic variables. However, BMI was not included, which is an important confounder for liver outcomes. While BMI was not included in the final model, many relevant confounders were addressed, making the risk probably low. |
| **ATTRITION/ EXCLUSION BIAS**<br>Were outcome data complete without attrition | Probably low | The study clearly described the exclusion criteria, including participants with missing phthalate metabolite or liver enzyme data, significant alcohol consumption, hepatitis or hepatic disease, pregnancy, or extremely high AST/ALT ratios. Although some participants were excluded due to missing or |

| | | |
|---|---|---|
| or exclusion from analysis? | | partial data, these exclusions were transparently reported and justified. Since the study was cross-sectional and not longitudinal, follow-up was not applicable. Overall, the clear reporting and rationale support a rating of probably low risk. |
| **DETECTION BIAS**<br>Can we be confident in the exposure characterization? * | Definitely low | The study did not detail the exposure method, but KoNEHS II used a validated and standardized technique (HPLC–ESI–MS/MS) to quantify urinary phthalate metabolites. Therefore, the risk of exposure detection bias is definitely low. |
| Can we be confident in the outcome assessment? * | Prabably low | The study used the hepatic steatosis index (HSI), a widely accepted non-invasive biomarker for assessing NAFLD in large population-based studies. The HSI incorporates ALT, AST, BMI, sex, and diabetes status, and a validated cut-off of 36 was applied. While liver biopsy is the gold standard, HSI is widely accepted and practical for large-scale studies. Thus, outcome assessment is considered reasonably accurate, supporting a probably low risk of bias. |
| **SELECTIVE REPORTING BIAS**<br>Were all measured outcomes reported? | Probably low | The study clearly described its objectives and reported results for the primary associations between urinary phthalate metabolites and NAFLD, as outlined in the methods section. Statistical analysis plans were detailed, including covariate adjustments and subgroup analyses. Outcomes related to NAFLD prevalence and urinary phthalates were consistently reported in both the methods and results sections. There is no indication that outcomes were selectively omitted or that only significant results were selectively highlighted. Therefore, the risk of selective reporting bias is rated as probably low. |
| **SELECTION BIAS**<br>Did selection of study participants result in appropriate comparison groups? | Probably low | Participants were selected from the Korean National Environmental Health Survey II (2012–2014), which used a national, multistage, stratified, and clustered probability sampling method to ensure representativeness of the South Korean adult population. Although the study excluded some individuals due to missing data, excessive alcohol intake, hepatitis, or pregnancy, these criteria were clearly reported and justified. However, the included NAFLD and non-NAFLD groups showed differences in several baseline characteristics such as age, sex, and BMI. These variables were adjusted for in the multivariable models, mitigating concerns of biased selection. Given the large-scale population-based design and statistical adjustments, the risk of selection bias is considered probably low. |
| **CONFLICT OF INTEREST** | Definitely low | "The authors declare no conflict of interest." |

***Risk of bias of Cai et al.2021,*** according to instructions reported in Appendix 3:

| **Bias Domain** | **Risk of Bias** | **Comments** |
|---|---|---|
| **CONFOUNDING BIAS. ***<br>Did the study design or analysis account for important confounding and modifying variables? | Definitely low | The study adjusted for a comprehensive range of potential confounding variables, including age, sex, race, marital status, education level, poverty income ratio (PIR), smoking, physical activity, BMI, alcohol consumption, hypertension, total cholesterol, and survey cycle. These factors cover major demographic, socioeconomic, behavioral, and metabolic confounders relevant to both exposure and outcomes. Therefore, the risk of confounding bias is |

| | | |
|---|---|---|
| | | considered definitely low. |
| **ATTRITION/ EXCLUSION BIAS** Were outcome data complete without attrition or exclusion from analysis? | Probably low | The study clearly described the exclusion criteria, including participants with viral hepatitis (HBsAg or anti-HCV positivity), pregnancy, steatogenic medication use, and missing covariate or fasting data. Although a large number of participants were excluded, these exclusions were transparently reported and appropriately justified. Since the study was cross-sectional and not longitudinal, loss to follow-up was not applicable. Overall, the clear reporting and rationale support a rating of probably low risk. |
| **DETECTION BIAS** Can we be confident in the exposure characterization? * | Definitely low | The study employed validated and standardized methods for measuring urinary phthalate metabolites, including high-performance liquid chromatography-electrospray ionization-tandem mass spectrometry (HPLC-ESI-MS/MS). All biospecimens were collected, processed, and tested by the National Center for Environmental Health (NCEH) under NHANES protocols with comprehensive quality control. Urinary metabolite levels were creatinine-standardized, and values below the detection limit were handled using a defined imputation method. These methodological strengths justify a rating of definitely low risk for exposure detection bias. |
| Can we be confident in the outcome assessment? * | Probably low | The study used the hepatic steatosis index (HSI), a widely accepted non-invasive biomarker for assessing NAFLD in large population-based studies. The HSI incorporates ALT, AST, BMI, sex, and diabetes status, and a validated cut-off of 36 was applied. While liver biopsy is the gold standard, HSI is widely accepted and practical for large-scale studies. Thus, outcome assessment is considered reasonably accurate, supporting a probably low risk of bias. |
| **SELECTIVE REPORTING BIAS** Were all measured outcomes reported? | Probably low | The study clearly stated its objectives and consistently reported results for the associations between individual urinary phthalate metabolites and two validated indices of NAFLD (HSI and USFLI). Both significant and non-significant findings were reported across different models with varying covariate adjustments. There is no indication of selective omission or overemphasis of favorable results. Therefore, the risk of selective reporting bias is rated as probably low. |
| **SELECTION BIAS** Did selection of study participants result in appropriate comparison groups? | Probably low | Participants were selected from the NHANES 2003–2016 cycles, which employed a nationally representative, multistage, stratified sampling design to ensure generalizability to the US adult population. Although the study excluded individuals with missing data, viral hepatitis, pregnancy, or steatogenic medication use, these criteria were clearly defined and appropriately justified. The included NAFLD and non-NAFLD groups showed differences in baseline characteristics such as age, sex, and BMI, but these variables were adjusted for in multivariable models. Given the population-based design and covariate adjustment, the risk of selection bias is considered probably low. |
| **CONFLICT OF INTEREST** | Definitely low | “The authors declare that they have no known competing financial interests or personal relationships that could have appeared to influence the work reported in this paper.” |

***Risk of bias of Chen et al.2022,*** according to instructions reported in Appendix 3:

| **Bias Domain** | **Risk of Bias** | **Comments** |
|---|---|---|
| **CONFOUNDING BIAS. *** Did the study design or analysis account for important confounding and modifying variables? | Probably low | The study adjusted for age, sex, ethnicity, educational level, BMI, diabetes, smoking status, physical activity, total cholesterol, and systolic blood pressure. Although alcohol consumption was not included, participants with excessive alcohol intake were excluded from the analysis, partly removing the potential confounding effect of alcohol consumption. Therefore, the risk of confounding bias is considered probably low. |
| **ATTRITION/ EXCLUSION BIAS** Were outcome data complete without attrition or exclusion from analysis? | Probably low | The study clearly described the exclusion criteria, including participants with missing data, viral hepatitis B or C, significant alcohol consumption, and those ineligible for VCTE exam. Although a considerable number of participants were excluded (e.g., 2,970 lacked urinary phthalate data), these exclusions were transparently reported and appropriately justified. Moreover, since the study was cross-sectional and not longitudinal, there was no issue of loss to follow-up. Given the detailed reporting and clear rationale for exclusions, the risk of attrition or exclusion bias is considered probably low. |
| **DETECTION BIAS** Can we be confident in the exposure characterization? * | Definitely low | The study used validated and standardized methods, including HPLC-ESI-MS/MS, to measure urinary phthalate metabolites. All analyses followed NHANES protocols with quality control. Values below the detection limit were imputed using standard methods. Therefore, the risk of exposure detection bias is considered definitely low. |
| Can we be confident in the outcome assessment? * | Definitely low | NAFLD was assessed using the controlled attenuation parameter (CAP) via FibroScan, a validated and widely used imaging technique for detecting hepatic steatosis. The examinations were performed by trained technicians following standardized procedures, including fasting, minimum measurement criteria, and quality control parameters. Although liver biopsy is the gold standard, CAP provides a reliable and non-invasive alternative suitable for population-based studies. Given its diagnostic accuracy and standardized implementation in NHANES, the risk of outcome misclassification is considered definitely low. |
| **SELECTIVE REPORTING BIAS** Were all measured outcomes reported? | Probably low | The study explicitly stated its objectives and reported results for the associations between individual urinary phthalate metabolites and NAFLD diagnosed by CAP. Both significant and non-significant findings were presented with multivariable adjustments. There was no indication of selective omission or emphasis of favourable outcomes. Therefore, the risk of selective reporting bias is considered probably low. |
| **SELECTION BIAS** Did selection of study participants result in appropriate comparison groups? | Probably low | Participants were selected from the NHANES 2017–2018 cycle using a multistage, stratified, and nationally representative sampling design. Individuals with viral hepatitis, significant alcohol use, or missing data were excluded based on pre-defined and justified criteria. Although the NAFLD and non-NAFLD groups differed in variables such as age, sex, BMI, diabetes, and phthalate levels, these covariates were appropriately adjusted for in the multivariable models. Given the representative sampling strategy and appropriate statistical control, the risk of selection bias is considered probably low. |

| | | |
|---|---|---|
| **CONFLICT OF INTEREST** | Definitely low | "The authors declare that the research was conducted in the absence of any commercial or financial relationships that could be construed as a potential conflict of interest." |

***Risk of bias of Yun et al.2025***, according to instructions reported in Appendix 3:

| **Bias Domain** | **Risk of Bias** | **Comments** |
|---|---|---|
| **CONFOUNDING BIAS. *** Did the study design or analysis account for important confounding and modifying variables? | Probably low | The study adjusted for age, sex, education, diabetes, dyslipidemia, hypertension, regular exercise, and smoking. However, BMI and alcohol consumption were not adjusted for, the study excluded participants with excessive alcohol intake during recruitment, which reduces its confounding effect. Therefore, the risk of confounding bias is considered probably low. |
| **ATTRITION/ EXCLUSION BIAS** Were outcome data complete without attrition or exclusion from analysis? | Probably low | The study clearly defined and justified exclusion criteria, including known liver diseases, missing survey or blood data, abnormal AST/ALT ratios, suggestive of alcoholic liver disease, and heavy alcohol consumption. All exclusions were transparent and occurred before analysis. As this was a cross-sectional study without follow-up, attrition bias is considered probably low. |
| **DETECTION BIAS** Can we be confident in the exposure characterization? * | Definitely low | The study used validated and standardized methods to measure serum PFAS levels. Blood samples were processed following strict protocols, including freezing at −70°C and extraction using acetonitrile. Quantification was performed using HPLC-MS/MS with clearly defined limits of detection for each PFAS compound. These procedures ensure high analytical accuracy and reliability. Therefore, the risk of exposure detection bias is considered definitely low. |
| Can we be confident in the outcome assessment? * | Probably low | The study used the hepatic steatosis index (HSI), a widely accepted non-invasive biomarker for assessing NAFLD in large population-based studies. The HSI incorporates ALT, AST, BMI, sex, and diabetes status, and a validated cut-off of 36 was applied. While liver biopsy is the gold standard, HSI is widely accepted and practical for large-scale studies. Thus, outcome assessment is considered reasonably accurate, supporting a probably low risk of bias. |
| **SELECTIVE REPORTING BIAS** Were all measured outcomes reported? | Probably low | The study clearly described its objectives and reported associations between serum PFAS levels and NAFLD. Both significant and non-significant results were presented across different models. There was no indication of selective omission or emphasis on favourable findings. Therefore, the risk of selective reporting bias is considered probably low. |
| **SELECTION BIAS** Did selection of study participants result in appropriate comparison groups? | Probably low | Participants were selected from the 4th cycle of the KoNEHS using a nationwide representative sampling strategy. Individuals with viral hepatitis, liver cancer, cirrhosis, missing data, and excessive alcohol consumption were excluded based on predefined and justified criteria. Although NAFLD and non-NAFLD groups differed in variables such as age, sex, education, and comorbidities, these covariates were adjusted for in multivariable analysis. Therefore, the risk of selection bias is considered probably low. |
| **CONFLICT OF INTEREST** | Definitely low | The authors declare no conflicts of interest. |

***Risk of bias of Wu et al.2024,*** according to instructions reported in Appendix 3:

| **Bias Domain** | **Risk of Bias** | **Comments** |
|---|---|---|
| **CONFOUNDING BIAS. *** Did the study design or analysis account for important confounding and modifying variables? | Probably low | The study adjusted for a comprehensive range of potential confounders, including age, sex, monthly household income, occupation, BMI, hypertension, fasting blood glucose, smoking, and drinking tea. However, alcohol consumption was not adjusted for, the study excluded participants with excessive alcohol intake, which reduces its confounding effect. Therefore, the risk of confounding bias is considered probably low. |
| **ATTRITION/ EXCLUSION BIAS** Were outcome data complete without attrition or exclusion from analysis? | Definitely low | This nested case-control study applied clear and justified exclusion criteria, including missing B-ultrasound data, invalid PFAS data, missing demographic variables, and loss to follow-up. Participants with viral hepatitis, cirrhosis, hepatocellular cancer, or a history of fatty liver disease were also excluded. The loss to follow-up rates across three waves (2.68%, 12.74%, and 15.56%) were explicitly reported, demonstrating transparency. Given the reasonable follow-up rates and detailed exclusion process, the risk of attrition/exclusion bias is considered definitely low. |
| **DETECTION BIAS** Can we be confident in the exposure characterization? * | Definitely low | PFAS concentrations were measured using validated protocols, including fasting blood collection, −80 °C storage, and analysis with HPLC-Triple Quadrupole Mass Spectrometry. Six PFAS with detection rates ≥90% were analyzed. Detection limits were clearly reported, and values below the limit were imputed using LOD/ √ 2. These standardized methods ensure high reliability, indicating a definitely low risk of exposure detection bias. |
| Can we be confident in the outcome assessment? * | Definitely low | NAFLD was diagnosed using standardized B-ultrasound based on national guidelines and confirmed by at least two ultrasound experts. Although liver biopsy is considered the gold standard, it is rarely used in large-scale epidemiological studies. Therefore, ultrasound is widely accepted as the practical gold standard in this context. The exclusion of other liver diseases further supports diagnostic validity. Hence, the risk of outcome detection bias is definitely low. |
| **SELECTIVE REPORTING BIAS** Were all measured outcomes reported? | Probably low | The study clearly reported its objectives, methodology, exposure and outcome assessment, and provided detailed results for all main variables. Although a pre-registered protocol was not mentioned, the comprehensive and transparent reporting suggests a low likelihood of selective reporting. Thus, the risk of selective reporting bias is considered probably low. |
| **SELECTION BIAS** Did selection of study participants result in appropriate comparison groups? | Probably low | This nested case-control study was based on the Jinchang cohort, which enrolled participants through a defined survey process and collected blood samples for PFAS assessment. Participants with missing data, invalid serum PFAS measurements, or certain liver conditions were excluded based on predefined and justified criteria. The selection of 476 NAFLD cases and 952 matched controls was performed using clear matching variables (age and sex). Although residual confounding cannot be completely ruled out, the use of a well-defined population and transparent selection process suggests the risk of |

| | | |
|---|---|---|
| | | selection bias is probably low. |
| **CONFLICT OF INTEREST** | Definitely low | The authors declare that they have no known competing financial interests or personal relationships that could have appeared to influence the work reported in this paper. |

***Risk of bias of Wu et al.2023***, according to instructions reported in Appendix 3:

| **Bias Domain** | **Risk of Bias** | **Comments** |
|---|---|---|
| **CONFOUNDING BIAS. ***<br>Did the study design or analysis account for important confounding and modifying variables? | Probably low | The study adjusted for age, sex, ethnicity, family income-to-poverty ratio, BMI, diabetes, hypertension, physical activity, cigarettes per day, healthy eating index (HEI) score, and total blood cholesterol level. However, alcohol consumption was not adjusted for, the study excluded participants with excessive alcohol intake, which reduces its confounding effect. Therefore, the risk of confounding bias is considered probably low. |
| **ATTRITION/**<br>**EXCLUSION BIAS**<br>Were outcome data complete without attrition or exclusion from analysis? | Probably low | The study clearly defined and justified exclusion criteria, including missing data on covariates, PFAS exposures, or key clinical factors, and excluded individuals with viral hepatitis, significant alcohol use, steatogenic medications, and pregnancy. All exclusions were transparent and occurred before analysis. As this was a cross-sectional study without follow-up, attrition bias is considered probably low. |
| **DETECTION BIAS**<br>Can we be confident in the exposure characterization? * | Definitely low | The study measured serum PFASs (PFOA, PFOS, PFNA, PFHxS) using validated protocols, including automated solid-phase extraction and HPLC-MS/MS. For values below the lower detection limit (LOD), the study imputed concentrations using the LOD divided by the square root of two, a widely accepted approach. The use of standardized and robust analytical procedures ensures high reliability of exposure assessment. Therefore, the risk of exposure detection bias is considered definitely low. |
| Can we be confident in the outcome assessment? * | Probably low | The study defined NAFLD using the hepatic steatosis index (HSI), a widely used non-invasive algorithm based on ALT, AST, BMI, sex, and diabetes status. Although HSI is not the gold standard, it has been validated in population-based studies and offers a practical alternative to biopsy. Given the standardized formula and prior validation, the risk of outcome misclassification is limited, supporting a probably low rating. |
| **SELECTIVE REPORTING BIAS**<br>Were all measured outcomes reported? | Probably low | The study clearly stated its objectives, exposure and outcome assessments, and provided detailed results for all PFASs and statistical models described in the methods. Although a preregistered analysis protocol was not mentioned, the comprehensive reporting of variables, subgroup analyses, and statistical adjustments suggests a low likelihood of selective reporting. Therefore, the risk of selective reporting bias is considered probably low. |
| **SELECTION BIAS**<br>Did selection of study participants result in appropriate comparison groups? | Probably low | The study was based on a nationally representative survey (NHANES) using a multistage, probability sampling strategy. Participants aged ≥60 years with complete exposure and covariate data were included. Exclusions were based on clear clinical criteria, such as viral hepatitis, steatogenic medication, and significant alcohol consumption, aiming to minimize confounding. Although the analysis focused on a specific subpopulation, the use of defined criteria |

| | | |
|---|---|---|
| | | and a representative source population suggests a low risk of bias. Thus, the risk of selection bias is considered probably low. |
| **CONFLICT OF INTEREST** | Definitely low | "The authors declare no competing interests." |

***Risk of bias of Zhang et al.2023,*** according to instructions reported in Appendix 3:

| **Bias Domain** | **Risk of Bias** | **Comments** |
|---|---|---|
| **CONFOUNDING BIAS. *** Did the study design or analysis account for important confounding and modifying variables? | Definitely low | The study adjusted for a comprehensive set of potential confounders, including age, sex, ethnicity, smoking history (ever smoker), alcohol consumption (ever drink alcohol), total physical activity, BMI, history of diabetes, cancer, hypertension, aspirin use, and high-fat diet. These variables cover key demographic, lifestyle, and dietary factors that are commonly associated with both exposure and outcome. Therefore, the risk of confounding bias is considered definitely low. |
| **ATTRITION/ EXCLUSION BIAS** Were outcome data complete without attrition or exclusion from analysis? | Probably low | The study clearly defined and justified its exclusion criteria, including participants younger than 20 years, those without valid serum PFAS measurements, VCTE assessments, or demographic data, as well as individuals with viral hepatitis or on steatogenic medications. These exclusions were applied transparently before analysis. Since the study used cross-sectional NHANES data without follow-up, the risk of attrition bias is considered probably low. |
| **DETECTION BIAS** Can we be confident in the exposure characterization? * | Definitely low | The study assessed serum PFAS concentrations online solid-phase extraction coupled with high-performance liquid chromatography – turboionspray ionization – tandem mass spectrometry (HPLC-TIS-MS/MS). Only PFAS compounds detected in ≥80% of participants were included in the analysis, and the lower limit of detection (LOD) was 0.10 ng/mL. This rigorous and transparent approach, including exclusion of low-detection PFAS and summing of relevant compounds for total PFAS estimation, minimizes measurement error. Therefore, the risk of exposure detection bias is considered definitely low. |
| Can we be confident in the outcome assessment? * | Definitely low | The study diagnosed NAFLD using vibration-controlled transient elastography (VCTE), with CAP values >285 dB/m, a widely used non-invasive technique for assessing hepatic steatosis. While liver biopsy is the gold standard, VCTE has been validated in large population-based studies and provides a reliable alternative with good sensitivity and specificity for detecting steatosis. Therefore, the risk of outcome detection bias is considered definitely low. |
| **SELECTIVE REPORTING BIAS** Were all measured outcomes reported? | Probably low | The study clearly stated its objectives, exposure, and outcome assessments. It provided detailed results for all included PFAS compounds and reported effect estimates across different statistical, including adjustments for multiple covariates. Although a preregistered analysis protocol was not mentioned, the consistent presentation of subgroup analyses, exposure quantiles, sensitivity analyses, and full data tables in both the main and supplementary materials indicate comprehensive reporting. Therefore, the risk of selective reporting bias is considered probably low. |

| | | |
|---|---|---|
| **SELECTION BIAS**<br>Did selection of study participants result in appropriate comparison groups? | Probably low | The study clearly described its objectives, exposure and outcome assessments, and presented detailed results for all relevant variables. Although a preregistered analysis protocol was not mentioned, the use of publicly available NHANES data, comprehensive reporting of variables and covariates, and clear statistical methods suggest a low likelihood of selective reporting. Therefore, the risk of selective reporting bias is considered probably low. |
| **CONFLICT OF INTEREST** | Probably high | "AD has provided volunteer and paid consultation for communities exposed to PFAS-contaminated drinking water." "Dr. Xuehong Zhang is supported by National Cancer Institute U01 CA259208-01A1, American Cancer Society Research Scholar Grant (RSG NEC-130476), and American Cancer Society Interdisciplinary Team Award (PASD-22-1003396-01)." Although the conflict of interest was disclosed, the authors did not report any measures to mitigate its potential influence on the study outcomes. Therefore, the risk of conflict of interest bias is considered probably high. |

***Risk of bias of Cheng et al.2023,*** according to instructions reported in Appendix 3:

| **Bias Domain** | **Risk of Bias** | **Comments** |
|---|---|---|
| **CONFOUNDING BIAS. ***<br>Did the study design or analysis account for important confounding and modifying variables? | Probably low | The study adjusted for age, sex, ethnicity, education, and PIR, and excluded participants with excess alcohol intake and low BMI (<18.5). However, key metabolic factors (e.g., diabetes, hypertension, physical activity.) were not fully adjusted for, which may have led to residual confounding. Therefore, the risk of exposure detection bias is probably. |
| **ATTRITION/**<br>**EXCLUSION BIAS**<br>Were outcome data complete without attrition or exclusion from analysis? | Probably low | The exclusion criteria were clearly defined and transparently reported, including reasons such as age under 20, viral hepatitis, excessive alcohol consumption, and low BMI (<18.5). As the study was cross-sectional in design and did not involve longitudinal follow-up, the risk of attrition bias is considered probably low. |
| **DETECTION BIAS**<br>Can we be confident in the exposure characterization? * | Definitely low | PFAS levels were measured using SPE-HPLC-TIS-MS/MS, a standard and reliable lab technique. Blood samples were stored at –30°C before testing. Values below the detection limit were handled by replacing them with LLOD divided by the square root of 2, which is a common and accepted method. The coefficient of variation was kept within 15%, suggesting consistent and accurate measurements. Based on this, the risk of exposure detection bias is considered low. |
| Can we be confident in the outcome assessment? * | Definitely low | Hepatic steatosis was assessed using the CAP via FibroScan, which is a well-established and non-invasive method commonly used in population studies. While liver biopsy remains the gold standard, it's not practical in large-scale research. CAP has shown good sensitivity and specificity and is widely accepted for detecting liver fat. Therefore, the risk of outcome assessment bias is considered definitely low. |
| **SELECTIVE REPORTING BIAS**<br>Were all measured outcomes reported? | Probably low | The study clearly stated its objectives, exposures, and outcomes, and reported all results for all PFAS compounds. Effect estimates were shown with adjustments for key covariates, and subgroup analyses, exposure quantiles, and sensitivity analyses were all presented in detail. Although the study wasn't |

| | | |
|---|---|---|
| | | preregistered, the way results were presented in both the main text and the supplementary materials suggests a comprehensive approach to reporting. Therefore, the risk of selective reporting bias seems probably low. |
| **SELECTION BIAS**<br>Did selection of study participants result in appropriate comparison groups? | Probably low | Participants were selected from the NHANES 2017–2018 cycle, using standardized inclusion and exclusion criteria. Both the NAFLD and healthy groups came from the same population base and were assessed within the same time frame. The study reported detailed baseline characteristics, showing good comparability between groups. However, since the data were limited to a single survey cycle, the findings may not fully reflect broader or long-term patterns. So, the risk of selection bias is considered probably low. |
| **CONFLICT OF INTEREST** | Definitely low | "The authors declare that they do not have any conflicts of interest related to this study." |

***Risk of bias of Zhang et al.2024,*** according to instructions reported in Appendix 3:

| **Bias Domain** | **Risk of Bias** | **Comments** |
|---|---|---|
| **CONFOUNDING BIAS. ***<br>Did the study design or analysis account for important confounding and modifying variables? | Probably low | The study adjusted for age, sex, marital status, education, smoking, drinking, and physical activity. However, key metabolic indicators (e.g., BMI, diabetes, and hypertension) were not included in the models. Given that MASLD is closely linked to metabolic dysfunction, the lack of these adjustments may introduce residual confounding. Therefore, the risk of confounding bias is considered probably low. |
| **ATTRITION/ EXCLUSION BIAS**<br>Were outcome data complete without attrition or exclusion from analysis? | Probably low | The exclusion criteria were clearly defined and transparently reported. Participants were excluded for reasons such as absence of abdominal B-scan ultrasonography data, presence of cirrhosis or chronic liver disease, self-reported hepatitis, incomplete information on alcohol consumption, or missing key variables. As the study was a cross-sectional analysis based on a nested cohort, there was no long-term follow-up. Therefore, the risk of attrition/exclusion bias is considered probably low. |
| **DETECTION BIAS**<br>Can we be confident in the exposure characterization? * | Definitely low | The study measured PFAS concentrations in serum using UPLC-MS/MS, which is a well-established and widely used method. The procedures for storing, preparing, and analyzing samples were clearly described. Internal standards were used to improve accuracy, and the treatment of values below the detection limit followed standard practice by replacing them with the LOD divided by the square root of two. Considering these factors, the risk of exposure detection bias is considered definitely low. |
| Can we be confident in the outcome assessment? * | Definitely low | The diagnosis of MAFLD was based on liver ultrasound imaging and metabolic criteria, which, although not the gold standard, are widely used and considered valid in large-scale population studies. Therefore, the risk of outcome assessment bias is considered definitely low. |
| **SELECTIVE REPORTING BIAS**<br>Were all measured outcomes reported? | Probably low | The study clearly stated its objectives, exposure measures, and outcomes. Results were comprehensively reported for all PFASs with detection rates above 80%, including subgroup analyses, exposure quantiles, and sensitivity analyses. Both the main text and supplementary materials provided detailed findings. Although the study was not preregistered, the consistency between |

| | | |
|---|---|---|
| | | the methods and results suggests that outcome reporting was transparent. Therefore, the risk of selective reporting bias is considered probably low. |
| **SELECTION BIAS**<br>Did selection of study participants result in appropriate comparison groups? | Probably low | The study used a nested case-control design within the Dongfeng-Tongji cohort, with clear inclusion and exclusion criteria. Participants with diabetes, cancer, and cardiovascular disease were excluded from the original cohort, and further exclusions were applied based on missing data, liver disease, or excessive alcohol intake. These criteria were consistently applied to all participants prior to outcome assessment. However, as the cohort consisted of retired workers from a single enterprise, the sample may not fully represent the general population. Despite this limitation in representativeness, the internal validity of the study is unlikely to be affected. Therefore, the risk of selection bias is considered probably low. |
| **CONFLICT OF INTEREST** | Definitely low | "The authors declare that they have no known competing financial interests or personal relationships that could have appeared to influence the work reported in this paper." |

***Risk of bias of Li et al.2020,*** according to instructions reported in Appendix 3:

| Bias Domain | Risk of Bias | Comments |
|---|---|---|
| **CONFOUNDING BIAS. *** Did the study design or analysis account for important confounding and modifying variables? | Probably low | The study adjusted for age, marital status, education level, metabolic syndrome, insulin resistance, smoking, and drinking. However, it did not adjust for BMI. The absence of this adjustment may introduce residual confounding. Therefore, the risk of confounding bias is considered probably low. |
| **ATTRITION/ EXCLUSION BIAS** Were outcome data complete without attrition or exclusion from analysis? | Probably low | The study clearly defined and transparently reported exclusion criteria for both the cross-sectional and longitudinal datasets. Reasons for exclusion included missing information, excessive alcohol consumption, hepatic disease, and unwillingness to participate. Among the 1594 participants at baseline, 566 were followed up. A total of 88 participants were lost to follow-up, representing an attrition rate of 6.96 percent. Since the exclusion criteria were pre-defined and consistently applied, and the attrition rate was relatively low with clear reasons provided, the risk of attrition or exclusion bias is considered probably low. |
| **DETECTION BIAS** Can we be confident in the exposure characterization? * | Definitely low | The study measured serum metal concentrations using inductively coupled plasma mass spectrometry (ICP-MS), a widely accepted and sensitive method for trace metal detection. Values below the detection limit were imputed as the detection limit divided by the square root of two. Only metals with detection rates above 80 percent were included in the analysis. Considering these factors, the risk of exposure detection bias is considered definitely low. |
| Can we be confident in the outcome assessment? * | Definitely low | The diagnosis of NAFLD was based on liver ultrasound imaging and metabolic criteria, which, although not the gold standard, are widely used and considered valid in large-scale population studies. Therefore, the risk of outcome assessment bias is considered definitely low. |
| **SELECTIVE REPORTING BIAS** Were all measured outcomes reported? | Probably low | The study clearly described its objectives, exposure assessments, and outcomes. Statistical analyses were reported for all serum metals with detection rates above 80 percent, including subgroup and sensitivity analyses. Both the main text and supplementary materials provided detailed findings. Although the study was not preregistered, the consistency between the methods and reported results suggests that selective reporting is unlikely. Therefore, the risk of selective reporting bias is considered probably low. |
| **SELECTION BIAS** Did selection of study participants result in appropriate comparison groups? | Probably low | The study recruited participants from a hospital-based cohort of Chinese men in Guangxi, China, through the Fangchenggang Area Male Health and Examination Survey (FAMHES). Although the population was limited to men and derived from a single hospital, the study included a relatively large sample size and applied clear inclusion and exclusion criteria. There is no indication of differential selection related to exposure or outcome. Therefore, the risk of selection bias is considered probably low. |
| **CONFLICT OF INTEREST** | Definitely low | "The authors declare that they have no known competing financial interests or personal relationships that could have appeared to influence the work reported in this paper." |

***Risk of bias of Zhang et al.2023***, according to instructions reported in Appendix 3:

| **Bias Domain** | **Risk of Bias** | **Comments** |
|---|---|---|
| **CONFOUNDING BIAS. ***<br>Did the study design or analysis account for important confounding and modifying variables? | Definitely low | The study adjusted for a comprehensive set of potential confounders, including age, occupation, BMI, diabetes, hypertension, smoking status, drinking status, total energy intake, triglycerides, high-density lipoprotein, alanine transaminase, and the other six blood metal concentrations. These factors are major confounders known to influence both exposure and outcome in studies of metabolic liver disease. Therefore, the risk of confounding bias is considered definitely low. |
| **ATTRITION/ EXCLUSION BIAS**<br>Were outcome data complete without attrition or exclusion from analysis? | Probably low | The study clearly defined and transparently reported exclusion criteria. Out of 1896 male participants from the Kailuan cohort, 348 were excluded due to predefined reasons, including lack of abdominal ultrasound data, excessive alcohol consumption, hepatitis B, liver cirrhosis, cancer, or missing covariate data. These exclusion criteria were consistently applied. Among the remaining 1548 participants, 648 NAFLD and 648 non-NAFLD subjects were randomly selected with age-group frequency matching. Since the exclusion process was systematic and clearly justified, the risk of attrition or exclusion bias is considered probably low. |
| **DETECTION BIAS**<br>Can we be confident in the exposure characterization? * | Definitely low | The study measured seven blood metal concentrations (Ca, Cu, Fe, Mg, Mn, Se, and Zn) using inductively coupled plasma mass spectrometry (ICP-MS), a highly sensitive and widely accepted method for trace element detection. Intra-assay and inter-assay coefficients of variation were reported and within acceptable ranges, demonstrating good reliability. For metals with values below the detection limit the data were regarded as missing, rather than imputed, which helps avoid bias from inappropriate estimations. Therefore, the risk of exposure detection bias is considered definitely low. |
| Can we be confident in the outcome assessment? * | Definitely low | The diagnosis of NAFLD was based on liver ultrasound imaging and metabolic criteria, which, although not the gold standard, are widely used and considered valid in large-scale population studies. Therefore, the risk of outcome assessment bias is considered definitely low. |
| **SELECTIVE REPORTING BIAS**<br>Were all measured outcomes reported? | Probably low | The study clearly stated its objectives and reported the results for all seven measured blood metal concentrations (Ca, Cu, Fe, Mg, Mn, Se, and Zn). The data were systematically presented in the main text and tables, including statistical comparisons between NAFLD and non-NAFLD groups. There is no indication that any key outcomes were selectively omitted. Although no pre-registration or protocol was mentioned, the consistency between the stated aims, measured exposures, and reported results suggests that selective reporting is unlikely. Therefore, the risk of selective reporting bias is considered probably low. |
| **SELECTION BIAS**<br>Did selection of study participants result in appropriate comparison groups? | Probably low | The study used a case-control design. NAFLD cases were identified through abdominal ultrasonography, and non-NAFLD controls were randomly selected using age-group frequency matching. The study applied clear inclusion and exclusion criteria to minimize confounding and ensure comparability. Although the sample was limited to male participants, and derived from a specific cohort |

| | | |
|---|---|---|
| | | in Tangshan, China, the internal selection process was transparent and systematic. Therefore, the risk of selection bias is considered probably low. |
| **CONFLICT OF INTEREST** | Definitely low | "The authors declare that they have no known competing financial interests or personal relationships that could have appeared to influence the work reported in this paper." |

***Risk of bias of Park et al.2021****,* according to instructions reported in Appendix 3:

| **Bias Domain** | **Risk of Bias** | **Comments** |
|---|---|---|
| **CONFOUNDING BIAS. ***<br>Did the study design or analysis account for important confounding and modifying variables? | Definitely low | The study adjusted for age, sex, log-transformed blood lead levels, family income, education level, BMI, alcohol consumption, smoking status, physical activity, and survey year. These covariates were comprehensively considered and are known to influence both the exposure and the outcome. Therefore, the risk of confounding bias is considered definitely low. |
| **ATTRITION/<br>EXCLUSION BIAS**<br>Were outcome data complete without attrition or exclusion from analysis? | Probably low | The study clearly defined and reported the exclusion criteria. Participants were excluded if they had missing data on key covariates such as BMI, alcohol consumption, or physical activity, or if they had viral hepatitis, abnormal AST/ALT levels, or other liver diseases. The reasons for exclusion were reasonable and consistently applied. As the study was cross-sectional in design, there was no long-term follow-up. Therefore, the risk of attrition/exclusion bias is considered probably low. |
| **DETECTION BIAS**<br>Can we be confident in the exposure characterization? * | Definitely low | The study measured blood cadmium and lead concentrations using graphite furnace atomic absorption spectrometry (GFAAS), a well-established and widely accepted method for trace element detection. The limits of detection (LODs) were clearly reported, and values below the LOD were imputed using a standard approach (LOD divided by 2). Additionally, the laboratory adhered to rigorous internal and external quality control standards, including participation in U.S. CDC and German external quality assurance programs. Therefore, the risk of exposure detection bias is considered definitely low. |
| Can we be confident in the outcome assessment? * | Probably low | In this study, NAFLD was defined using HSI and FLI, which are based mainly on blood test results. While these are not the gold standard like liver biopsy or imaging techniques, they are non-invasive and widely accepted tools for identifying fatty liver in large-scale studies. Since the diagnosis relied on validated indices, even though not the most definitive methods, the risk of outcome detection bias is considered probably low. |
| **SELECTIVE REPORTING BIAS**<br>Were all measured outcomes reported? | Probably low | The study clearly stated its objectives and reported results for all seven measured blood metal concentrations. The data were systematically presented in both the main text and tables, with no signs that key outcomes were selectively omitted. Although the study didn't mention a pre-registration or protocol, the consistency between its aims, measured exposures, and reported findings suggests selective reporting is unlikely. Therefore, the risk of selective reporting bias is considered probably low. |
| **SELECTION BIAS**<br>Did selection of study participants result in | Probably low | The study used data from the Korea National Health and Nutrition Examination Survey (KNHANES), which employs a nationally representative, multistage, stratified, probability-based sampling design. Participants were randomly |

| | | |
|---|---|---|
| appropriate comparison groups? | | selected to reflect the general Korean population across sex, age, and region. Moreover, all participants included in the final analysis completed detailed questionnaires and physical examinations following standardized protocols. While some exclusions were made due to missing data or specific health conditions, these criteria were clearly defined and consistently applied. Therefore, the risk of selection bias is considered probably low. |
| **CONFLICT OF INTEREST** | Definitely low | "The authors disclose no conflicts" |

***Risk of bias of Maodong et al.2023***, according to instructions reported in Appendix 3:

| **Bias Domain** | **Risk of Bias** | **Comments** |
|---|---|---|
| **CONFOUNDING BIAS. ***<br>Did the study design or analysis account for important confounding and modifying variables? | Probably low | The study adjusted for age, sex, ethnicity, education level, family PIR, BMI, hypertension, diabetes, smoke status, triglyceride, total cholesterol, LDL-cholesterol, Direct HDL-Cholesterol and urinary creatinine. However, alcohol consumption was not adjusted for, the study excluded participants with excessive alcohol intake, which reduces its confounding effect. Therefore, the risk of confounding bias is considered probably low. |
| **ATTRITION/**<br>**EXCLUSION BIAS**<br>Were outcome data complete without attrition or exclusion from analysis? | Probably low | The study used data from the NHANES survey and clearly outlined the reasons for participant exclusion at each step, including age under 20, missing data for calculating USFLI, missing urinary barium data, hepatitis B/C infection, pregnancy, and excessive alcohol intake. As the study was cross-sectional in design, there was no long-term follow-up. Therefore, the risk of attrition/exclusion bias is considered probably low. |
| **DETECTION BIAS**<br>Can we be confident in the exposure characterization? * | Definitely low | Urinary barium levels were measured using inductively coupled plasma mass spectrometry (ICP-MS), a highly sensitive and reliable method for detecting trace elements in biological samples. The analysis followed standardized procedures outlined in the NHANES Laboratory/Medical Technologists Procedures Manual. Additionally, the detection limit (LOD) was clearly defined and consistently applied across survey cycles, with 94.6% of participants having detectable levels. Values below the LOD were imputed according to NHANES guidelines. Given the use of standardized and validated laboratory procedures, the risk of exposure detection bias is considered definitely low. |
| Can we be confident in the outcome assessment? * | Probably low | NAFLD was determined using the U.S. Fatty Liver Index (USFLI), a validated, non-invasive diagnostic tool that incorporates demographic and biochemical variables such as age, race, waist circumference, γ-glutamyltransferase, fasting insulin, and glucose levels. Although USFLI is not the gold standard (i.e., liver biopsy or imaging), it has been extensively used in epidemiological studies for liver disease assessment and provides a reliable, standardized alternative for large-scale population-based studies. Therefore, the risk of outcome detection bias is considered probably low. |
| **SELECTIVE REPORTING BIAS**<br>Were all measured outcomes reported? | Probably low | The study clearly stated its objectives and reported results for all relevant exposure and outcome variables. The primary exposure (urinary barium levels) and outcome (USFLI-defined NAFLD) were described in the methods section and systematically presented in the results. Key findings were reported in both |

| | | |
|---|---|---|
| | | tables and the main text, and subgroup or sensitivity analyses were also included. Although the study did not mention pre-registration or a published protocol, the consistency between the stated aims, methods, and reported findings suggests that selective reporting is unlikely. Therefore, the risk of selective reporting bias is considered probably low. |
| **SELECTION BIAS** Did selection of study participants result in appropriate comparison groups? | Probably low | The study used data from the NHANES 2005–2016, which employs a multistage, stratified, probability sampling design to represent the non-institutionalized U.S. population. The inclusion and exclusion criteria were clearly defined and systematically applied: individuals under 20 years old, those with missing data for urinary barium or USFLI, those with hepatitis B or C infection, pregnant women, and individuals with excessive alcohol intake were excluded. These exclusions were based on standard clinical and methodological considerations to avoid confounding. Since NHANES is designed to be representative and the exclusions are justified, the risk of selection bias is considered probably low. |
| **CONFLICT OF INTEREST** | Definitely low | "There are no conflicts of interest" |

***Risk of bias of Fan et al.2024,*** according to instructions reported in Appendix 3:

| **Bias Domain** | **Risk of Bias** | **Comments** |
|---|---|---|
| **CONFOUNDING BIAS. *** Did the study design or analysis account for important confounding and modifying variables? | Definitely low | The study adjusted for age, sex, ethnicity, marital status, PIR, BMI, smoking status and drink status. These covariates were comprehensively considered and are known to influence both the exposure and the outcome. Therefore, the risk of confounding bias is considered definitely low. |
| **ATTRITION/ EXCLUSION BIAS** Were outcome data complete without attrition or exclusion from analysis? | Probably low | The study used NHANES data from 2003–2018 and clearly documented the stepwise exclusion of participants. Reasons for exclusion included age under 20, missing values for key variables such as urinary arsenic, urinary creatinine, BMI, alcohol use, and hepatitis status. A detailed flowchart was provided to illustrate the exclusion process. As this was a cross-sectional analysis with no follow-up period, attrition due to loss during follow-up was not applicable. Therefore, the risk of attrition/exclusion bias is considered probably low. |
| **DETECTION BIAS** Can we be confident in the exposure characterization? * | Definitely low | Although the article did not explicitly describe the measurement method, the data were extracted from NHANES, where urinary arsenic is consistently measured using ICP-MS. Therefore, the risk of exposure detection bias is considered definitely low. |
| Can we be confident in the outcome assessment? * | Probably low | Although the article did not explicitly describe the diagnosis method, the data were extracted from NHANES, where NAFLD is consistently measured using FLI. Therefore, the risk of outcome detection bias is considered probably low. |
| **SELECTIVE REPORTING BIAS** Were all measured outcomes reported? | Probably low | The study clearly defined its objectives and reported the results for both the exposure and outcome. All key variables mentioned in the methods were analyzed and reported in the results. The study presented findings in both tables and figures and also conducted subgroup and sensitivity analyses. Although the article did not mention pre-registration or a published study |

| | | |
|---|---|---|
| | | protocol, the consistency between the stated objectives and reported findings suggests a low risk of selective outcome reporting. Therefore, the risk of selective reporting bias is considered probably low. |
| **SELECTION BIAS**<br>Did selection of study participants result in appropriate comparison groups? | Probably low | The study utilized NHANES data from 2003 to 2018, which applies a multistage, stratified probability sampling design to represent the non-institutionalized U.S. population. The inclusion and exclusion criteria were clearly stated and consistently applied. Participants under 20 years of age, those with missing data on urinary arsenic, urinary creatinine, BMI, alcohol use, and hepatitis infection were excluded. These exclusions were based on standard clinical or methodological considerations to reduce potential confounding. Since the sampling frame of NHANES is designed to ensure representativeness, and the applied criteria were justified, the risk of selection bias is considered probably low. |
| **CONFLICT OF INTEREST** | Definitely low | "The authors declare that they have no known competing financial interests or personal relationships that could have appeared to influence the work reported in this paper." |

***Risk of bias of Nguyen et al.2022,*** according to instructions reported in Appendix 3:

| **Bias Domain** | **Risk of Bias** | **Comments** |
|---|---|---|
| **CONFOUNDING BIAS. ***<br>Did the study design or analysis account for important confounding and modifying variables? | Definitely low | The study adjusted for age, sex, educational level, monthly household incomes, residential area BMI, family history of dyslipidemia, drinking status, physical activity, cotinine verified smokers and ln2-transformed creatinine levels. These covariates were comprehensively considered and are known to influence both the exposure and the outcome. Therefore, the risk of confounding bias is considered definitely low. |
| **ATTRITION/**<br>**EXCLUSION BIAS**<br>Were outcome data complete without attrition or exclusion from analysis? | Probably low | The study used data from the Korea National Health and Nutrition Examination Survey (KNHANES) from 2009–2013 and 2016–2017. The exclusion criteria were clearly documented, including missing serum heavy metal measurements, excessive alcohol consumption (defined as >21 drinks/week for men and >14 for women), positive hepatitis B/C virus markers, self-reported liver disease, and missing outcome/covariate data. A flowchart was also provided to illustrate the exclusion process. As this was a cross-sectional study without follow-up, attrition due to loss during follow-up is not applicable. Therefore, the risk of attrition/exclusion bias is considered probably low. |
| **DETECTION BIAS**<br>Can we be confident in the exposure characterization? * | Definitely low | The study provided detailed information on the methods used to assess serum heavy metals (mercury, lead, cadmium), including the use of validated instruments: graphite furnace atomic absorption spectrometry for lead and cadmium, and a direct mercury analyzer for mercury. The detection limits were explicitly stated, and internal quality assurance procedures using commercial standards were also described. These indicate the use of accurate and standardized exposure assessment techniques. Therefore, the risk of exposure detection bias is considered definitely low. |
| Can we be confident in the outcome assessment? * | Probably low | Although the study did not use imaging or biopsy, NAFLD was assessed using the Fatty Liver Index (FLI), a widely accepted surrogate marker based on |

| | | |
|---|---|---|
| | | metabolic parameters. The use of FLI is common in large-scale epidemiological studies. Therefore, the risk of outcome detection bias is considered probably low. |
| **SELECTIVE REPORTING BIAS**<br>Were all measured outcomes reported? | Probably low | The study clearly stated its objectives and reported results for all relevant exposure and outcome variables. Key variables were described in the methods and reported in both tables and the main text. Although the study did not mention pre-registration or a published study protocol, the consistency between the stated aims and reported findings suggests that selective reporting is unlikely. Therefore, the risk of selective reporting bias is considered probably low. |
| **SELECTION BIAS**<br>Did selection of study participants result in appropriate comparison groups? | Probably low | The study used data from the Korean National Health and Nutrition Examination Survey (KNHANES), which employs a multistage, stratified, probability-based sampling method to represent the non-institutionalized Korean population. The inclusion and exclusion criteria were clearly stated and systematically applied, including the exclusion of individuals without serum heavy metal data, excessive alcohol consumption, viral hepatitis, and missing covariate information. These exclusions were based on clinical and methodological justifications to reduce confounding. Therefore, the risk of selection bias is considered probably low. |
| **CONFLICT OF INTEREST** | Definitely low | "The authors declare that they have no known competing financial interests or personal relationships that could have appeared to influence the work reported in this paper." |

***Risk of bias of Xie et al.2023****,* according to instructions reported in Appendix 3:

| Bias Domain | Risk of Bias | Comments |
|---|---|---|
| **CONFOUNDING BIAS. ***<br>Did the study design or analysis account for important confounding and modifying variables? | Probably low | The study adjusted for age, sex, ethnicity, education, PIR, BMI, diabetes, hypertension, smoking status, physical activity and HDL cholesterol. These covariates were comprehensively considered and are known to influence both the exposure and the outcome. Although alcohol consumption was not included as a covariate, participants with excessive alcohol intake were excluded from the NAFLD analysis, minimizing residual confounding from alcohol. Therefore, the risk of confounding bias is considered probably low. |
| **ATTRITION/**<br>**EXCLUSION BIAS**<br>Were outcome data complete without attrition or exclusion from analysis? | Probably low | The study used data from eight continuous NHANES cycles (2003–2018). A large number of participants were excluded due to missing information on the U.S. Fatty Liver Index (USFLI) and key covariates such as education, income, smoking, and physical activity. In addition, those with excessive alcohol intake, positive hepatitis B/C markers were also excluded for the NAFLD analysis. These criteria were clearly explained, and a flowchart was provided. Since the study was cross-sectional without follow-up, attrition due to loss over time was not an issue. Therefore, the risk of attrition/exclusion bias is considered probably low. |
| **DETECTION BIAS**<br>Can we be confident in the exposure characterization? | Definitely low | Urinary heavy metals, including barium, lead, cadmium, mercury, and arsenic species, were measured using validated and standardized methods (mainly ICP-MS and ICP-DRC-MS for metals, and HPLC for arsenic species). These |

| | | |
|---|---|---|
| * | | techniques are well-recognized for their accuracy and sensitivity in biological samples. NHANES applied consistent quality control procedures, and the detection limits and imputation methods for values below the LOD were clearly stated. Overall, given the standardized laboratory protocols and high detection rates, the risk of exposure detection bias is considered definitely low. |
| Can we be confident in the outcome assessment? * | Probably low | Hepatic steatosis was assessed using the USFLI score, a validated non-invasive index based on lab and clinical data. A cutoff of ≥30 was used to define steatosis. While this is not as accurate as liver biopsy, it's a widely accepted method for large-scale population studies like NHANES. The criteria for MAFLD and NAFLD were clearly defined and based on objective measures (e.g., BMI, glucose, HbA1c). Given the use of validated tools and clear diagnostic thresholds, the risk of outcome detection bias is probably low. |
| **SELECTIVE REPORTING BIAS**<br>Were all measured outcomes reported? | Probably low | The study clearly stated its objectives and reported results for all relevant exposure (heavy metals) and outcome (NAFLD and MAFLD) variables. The key variables were described in the methods section and consistently reported in the results, including both significant and non-significant associations. Although the study did not mention pre-registration or a published protocol, the alignment between the study aims and reported findings suggests that selective reporting is unlikely. Therefore, the risk of selective reporting bias is considered probably low. |
| **SELECTION BIAS**<br>Did selection of study participants result in appropriate comparison groups? | Probably low | The study used data from the Korea National Health and Nutrition Examination Survey (KNHANES), which employs a stratified, multistage, probability-cluster sampling design to ensure national representativeness. The study population was selected based on clearly defined inclusion and exclusion criteria, such as age, availability of heavy metal measurements, and absence of viral hepatitis or excessive alcohol intake. These criteria were applied consistently, and the large sample size reduces the likelihood of systematic differences between included and excluded individuals. Therefore, the risk of selection bias is considered probably low. |
| **CONFLICT OF INTEREST** | Definitely low | "The authors declare that the research was conducted in the absence of any commercial or financial relationships that could be construed as a potential conflict of interest." |

***Risk of bias of Yang et al.2021,*** according to instructions reported in Appendix 3:

| **Bias Domain** | **Risk of Bias** | **Comments** |
|---|---|---|
| **CONFOUNDING BIAS. ***<br>Did the study design or analysis account for important confounding and modifying variables? | Definitely low | The study adjusted for age, sex, marital status, education, income, hypertension, diabetes, hyperlipidemia, smoking, drinking, exercise, seafood consumption within one week, HSI and blood mercury level. These covariates were comprehensively considered and are known to influence both the exposure and the outcome. Therefore, the risk of confounding bias is considered definitely low. |
| **ATTRITION/<br>EXCLUSION BIAS**<br>Were outcome data | Probably low | The study analyzed data from the Korean National Environmental Health Survey (KoNEHS) conducted in 2012–2014. A total of 6,478 adults (≥19 years old) were initially recruited. Participants were excluded if they had missing |

| | | |
|---|---|---|
| complete without attrition or exclusion from analysis? | | biochemical values (e.g., ALT or AST), significant alcohol consumption (defined as >3 drinks/day or >7 drinks/week for men and >2 drinks/day or >4 drinks/week for women), history of hepatic diseases, current pregnancy, or were taking antidiabetic or antihypertensive medications. These criteria were clearly described, and a flow diagram was provided to illustrate the exclusion process. As this was a cross-sectional study without follow-up, attrition due to loss over time was not applicable. Therefore, the risk of attrition/exclusion bias is considered probably low. |
| **DETECTION BIAS**<br>Can we be confident in the exposure characterization? * | Probably low | The study used data from the Korea National Environmental Health Survey (KoNEHS), which follows standardized national protocols for sample collection and heavy metal analysis. Although this paper didn't specify the mercury detection method, previous studies using the same KoNEHS dataset consistently reported that blood mercury levels were measured using the gold amalgam collection method with a DMA-80 analyzer. This method is well-recognized and reliable for mercury detection. Given the use of validated national procedures and the consistency across KoNEHS-based studies, the risk of exposure detection bias is considered probably low. |
| Can we be confident in the outcome assessment? * | Probably low | The study defined NAFLD using the hepatic steatosis index (HSI), a widely used non-invasive algorithm based on ALT, AST, BMI, sex, and diabetes status. Although HSI is not the gold standard, it has been validated in population-based studies and offers a practical alternative to biopsy. Given the standardized formula and prior validation, the risk of outcome misclassification is limited, supporting a probably low rating. |
| **SELECTIVE REPORTING BIAS**<br>Were all measured outcomes reported? | Probably low | The study clearly described its objectives and reported results for both exposure (urinary mercury) and outcomes (NAFLD and liver enzyme levels). Key variables were explained in the methods section and consistently presented in the results, including both significant and non-significant associations. Although the authors didn't mention a pre-registered protocol, the alignment between their stated aims and reported findings suggests that selective reporting is unlikely. Therefore, the risk of selective reporting bias is considered probably low. |
| **SELECTION BIAS**<br>Did selection of study participants result in appropriate comparison groups? | Probably low | The study used data from the Korean National Environmental Health Survey (KoNEHS), which is designed to represent the general Korean population using multistage stratified sampling. Participants were selected from adults aged ≥19 years with available data on urinary mercury and liver-related variables. Although the final sample size was reduced due to exclusion criteria (e.g., missing data, hepatitis, excessive alcohol intake), the use of a nationally representative sampling framework reduces concerns about selection bias. Therefore, the risk of selection bias is considered probably low. |
| **CONFLICT OF INTEREST** | Definitely low | "The authors declare no conflict of interest" |

***Risk of bias of Kim et al.2023,*** according to instructions reported in Appendix 3:

| **Bias Domain** | **Risk of Bias** | **Comments** |
|---|---|---|

| | | |
|---|---|---|
| **CONFOUNDING BIAS. ***<br>Did the study design or analysis account for important confounding and modifying variables? | Probably low | The study adjusted for age, sex, education, personal income, occupation, and smoking, which are relevant confounders. However, it did not adjust for BMI and alcohol consumption or other metabolic-related factors, which could also influence the risk of alcoholic liver disease. Therefore, the risk of confounding bias is considered probably low. |
| **ATTRITION/ EXCLUSION BIAS**<br>Were outcome data complete without attrition or exclusion from analysis? | Probably low | The study analyzed data from KNHANES 2010–2013 and 2016–2017 and clearly documented the sample selection and exclusion criteria. From a total of 14,862 participants, subjects were excluded for being under 19 years old, missing alcohol intake data, incomplete ALD/NAFLD index data, or having hepatitis B or C. These exclusion criteria were clearly reported, and a final sample of 11,993 participants was included in the analysis. As this is a cross-sectional study without longitudinal follow-up, attrition over time is not a concern. The transparent reporting of exclusion criteria and relatively small proportion of exclusions support a probably low risk of attrition/exclusion bias. |
| **DETECTION BIAS**<br>Can we be confident in the exposure characterization? * | Definitely low | The study clearly reported the detection methods for all included metals. Blood total mercury levels were measured using the gold amalgam collection method with a DMA analyzer, while cadmium and lead levels were measured using graphite furnace atomic absorption spectrometry (GFAAS). All of these are well-established and reliable techniques for heavy metal quantification. Therefore, the risk of exposure detection bias is considered definitely low. |
| Can we be confident in the outcome assessment? * | Probably low | The study defined alcoholic liver disease (ALD) using a composite diagnostic approach that included both alcohol consumption thresholds (≥210 g/week for men and ≥140 g/week for women) and an ALD/NAFLD index > 0. The index incorporates mean corpuscular volume (MCV), the AST/ALT ratio, BMI, and sex. Although not the gold standard (e.g., liver biopsy or imaging), it remains a standardized, non-invasive, and previously validated surrogate approach. Therefore, the risk of outcome detection bias is considered probably low. |
| **SELECTIVE REPORTING BIAS**<br>Were all measured outcomes reported? | Probably low | The study clearly stated its objectives and reported predefined outcomes. Results for mercury, cadmium, and lead were fully presented, with adjusted models and subgroup analyses included. Although there was no mention of a pre-registered protocol, the consistency between study aims and reported findings suggests selective reporting is unlikely. Therefore, the risk of selective reporting bias is considered probably low. |
| **SELECTION BIAS**<br>Did selection of study participants result in appropriate comparison groups? | Probably low | The study used data from the Korea National Health and Nutrition Examination Survey (KNHANES), which employed a complex, stratified, multi-stage probability sampling method designed to represent the civilian, non-institutionalized Korean population. This nationally representative sampling framework minimizes the risk of selection bias. Furthermore, clear inclusion and exclusion criteria were applied (e.g., excluding those under 19 years old, individuals with viral hepatitis, or missing data), and the final analysis included nearly 12,000 participants. Therefore, the risk of selection bias is considered probably low. |
| **CONFLICT OF INTEREST** | Definitely low | "The authors have no potential conflicts of interest to disclose." |

***Risk of bias of Wan et al.2022,*** according to instructions reported in Appendix 3:

| **Bias Domain** | **Risk of Bias** | **Comments** |
|---|---|---|
| **CONFOUNDING BIAS. *** Did the study design or analysis account for important confounding and modifying variables? | Probably high | The study adjusted for a limited set of covariates, specifically age, sex, and current smoking status. However, it did not account for key metabolic risk factors such as BMI, alcohol consumption, physical activity, diet, or other lifestyle-related variables. The omission of these important confounders may bias the observed associations between environmental pollutants and liver steatosis. Therefore, the risk of confounding bias is considered probably high. |
| **ATTRITION/ EXCLUSION BIAS** Were outcome data complete without attrition or exclusion from analysis? | Probably low | The study clearly reported the exclusion criteria, stating that participants who could not be diagnosed with NAFLD/MAFLD or had missing blood lead level (BLL) data were excluded. A total of 3,066 participants were included in the final analysis. As a cross-sectional study, attrition over time is not a concern. The exclusion criteria were transparently defined and applied, and the proportion of excluded participants appears reasonable. Therefore, the risk of attrition/exclusion bias is considered probably low. |
| **DETECTION BIAS** Can we be confident in the exposure characterization? * | Definitely low | The study measured blood lead levels using atomic absorption spectrometry a widely accepted and reliable method for quantifying lead in biological samples. The analysis was performed at a central laboratory under controlled conditions. Therefore, the risk of exposure detection bias is considered definitely low. |
| Can we be confident in the outcome assessment? * | Definitely low | Hepatic steatosis was diagnosed using abdominal ultrasonography based on standardized procedures. While liver biopsy remains the gold standard, ultrasound is a validated, non-invasive, and widely used method in population studies for detecting fatty liver. Therefore, the risk of outcome detection bias is considered definitely low. |
| **SELECTIVE REPORTING BIAS** Were all measured outcomes reported? | Probably low | The study clearly stated its objectives, which were to assess the association between blood lead levels and hepatic steatosis and reported relevant outcomes accordingly. Key variables such as blood lead levels, NAFLD prevalence, insulin resistance, and associated metabolic factors were presented in both unadjusted and adjusted models. Although there was no mention of a preregistered protocol, there is no indication that important outcomes were withheld or selectively omitted. Therefore, the risk of selective reporting bias is considered probably low. |
| **SELECTION BIAS** Did selection of study participants result in appropriate comparison groups? | Probably low | Participants were recruited from 23 sites across four provinces in East China using community-based sampling. Citizens aged 18 years or older who had lived in their current area for at least six months were included. Clear inclusion and exclusion criteria were applied, and individuals with missing blood lead level or liver disease data were excluded. A final sample of 3,066 participants was analyzed. Although this was not a randomized sample, the multi-site design and transparent reporting reduce the risk of systematic differences between included and excluded individuals. Therefore, the risk of selection bias is considered probably low. |
| **CONFLICT OF INTEREST** | Definitely | "The authors declare that they have no known competing financial interests or |

| | | |
|---|---|---|
| | low | personal relationships that could have appeared to influence the work reported in this paper." |

***Risk of bias of Ye et al.2024,*** according to instructions reported in Appendix 3:

| **Bias Domain** | **Risk of Bias** | **Comments** |
|---|---|---|
| **CONFOUNDING BIAS. *** Did the study design or analysis account for important confounding and modifying variables? | Definitely low | The study adjusted for a comprehensive set of confounders, including age, sex, ethnicity, marital status, education level, PIR, BMI, diabetes, hypertension, dyslipidemia, cardiovascular disease, chronic obstructive pulmonary disease, cancer, alcohol consumption, smoking status, physical activity, daily calorie intake, and zinc intake. These variables cover sociodemographic, metabolic, lifestyle, and dietary factors. Therefore, the risk of confounding bias is considered definitely low. |
| **ATTRITION/ EXCLUSION BIAS** Were outcome data complete without attrition or exclusion from analysis? | Probably low | The study clearly documented the exclusion criteria and reported the flow of participants from the initial pool of 29,902 to the final analytic sample of 3,398. Participants were excluded due to missing data on serum zinc levels, HSI/NFS components, or covariates, as well as due to pregnancy, age below 20, or a history of liver disease or excessive alcohol intake. These exclusions were transparent, based on objective criteria, and unrelated to the outcomes. As this was a cross-sectional study without follow-up, attrition over time is not applicable. Therefore, the risk of attrition/exclusion bias is considered probably low. |
| **DETECTION BIAS** Can we be confident in the exposure characterization? * | Definitely low | Serum zinc concentrations were measured using inductively coupled plasma dynamic reaction cell mass spectrometry (ICP-DRC-MS), a highly sensitive and validated method for trace metal detection. Stringent collection, storage, and processing procedures were followed, and gallium was used as the internal standard. The method is reliable and widely accepted for quantifying serum zinc levels in population-based studies. Therefore, the risk of exposure detection bias is considered definitely low. |
| Can we be confident in the outcome assessment? * | Probably low | NAFLD was diagnosed using the Hepatic Steatosis Index (HSI), a widely used non-invasive surrogate marker based on standardized clinical parameters. Although it is not the gold standard (e.g., imaging or liver biopsy), HSI is validated and commonly applied in large-scale population studies. The use of a consistent, non-invasive method supports a low risk of outcome detection bias. Therefore, the risk of outcome detection bias is considered probably low. |
| **SELECTIVE REPORTING BIAS** Were all measured outcomes reported? | Probably low | The study clearly stated its objectives and reported relevant outcomes, including associations between serum zinc levels, NAFLD, and advanced fibrosis. Both unadjusted and adjusted models were presented, and subgroup analyses were included. Although there was no mention of a preregistered protocol, there is no indication that important outcomes were selectively omitted. Therefore, the risk of selective reporting bias is considered probably low. |
| **SELECTION BIAS** Did selection of study participants result in | Probably low | The study utilized data from NHANES, a nationally representative cross-sectional survey using a multistage probability sampling design. Clear inclusion and exclusion criteria were applied, including removal of individuals with viral |

| | | |
|---|---|---|
| appropriate comparison groups? | | hepatitis, liver cancer, pregnancy, or excessive alcohol use. The final analysis included 3,398 participants. Although not a randomized study, the use of a representative sample and clear criteria reduces the risk of selection bias. Therefore, the risk of selection bias is considered probably low. |
| **CONFLICT OF INTEREST** | Definitely low | "The authors declare no competing interests." |

***Risk of bias of Liu et al.2024***, according to instructions reported in Appendix 3:

| **Bias Domain** | **Risk of Bias** | **Comments** |
|---|---|---|
| **CONFOUNDING BIAS. *** Did the study design or analysis account for important confounding and modifying variables? | Definitely low | The study adjusted for a comprehensive range of potential confounders, including age, sex, BMI, education, smoking status, alcohol consumption, physical activity, meat intake, and vegetable intake. These variables cover key demographic, lifestyle, and dietary factors that are commonly associated with both exposure and outcome. Therefore, the risk of confounding bias is considered definitely low. |
| **ATTRITION/ EXCLUSION BIAS** Were outcome data complete without attrition or exclusion from analysis? | Probably low | The study clearly documented the exclusion criteria and transparently reported the number of participants excluded due to missing urine samples, out-of-range age, or missing clinical measurements such as hepatic ultrasound, BMI, waist circumference, blood pressure, glucose, lipid levels, or renal function. These exclusions were based on objective criteria, and the final sample size remained large (3,651 participants). As this was a cross-sectional study without follow-up, attrition over time is not applicable. Therefore, the risk of attrition/exclusion bias is considered probably low. |
| **DETECTION BIAS** Can we be confident in the exposure characterization? * | Definitely low | Urinary concentrations of 20 metals were measured using inductively coupled plasma mass spectrometry (ICP-MS), a highly sensitive and widely accepted method for trace metal detection. Measurements were creatinine-adjusted to account for urine dilution, and the limits of detection ranged from 0.00004 to 0.25 μg/L. These procedures ensure accuracy and reliability in exposure quantification. Therefore, the risk of exposure detection bias is considered definitely low. |
| Can we be confident in the outcome assessment? * | Definitely low | MAFLD was diagnosed based on hepatic steatosis identified by ultrasound, along with the presence of overweight/obesity, diabetes, or metabolic dysregulation, following international expert consensus. Although ultrasound is not the gold standard, it is a widely used, validated, and accepted method for large-scale epidemiological studies. Therefore, the risk is considered definitely low. |
| **SELECTIVE REPORTING BIAS** Were all measured outcomes reported? | Probably low | The study clearly stated its primary objective and reported the main findings regarding associations between urinary metal levels and MAFLD. Both unadjusted and adjusted models were presented, and subgroup analyses were conducted. Although the study did not mention a pre-registered protocol, there is no indication that important outcomes were selectively omitted. Therefore, the risk of selective reporting bias is considered probably low. |
| **SELECTION BIAS** Did selection of study | Probably low | Participants were recruited from the Medical Physical Examination Center of Tongji Hospital, and inclusion/exclusion criteria were clearly defined. A total of |

| | | |
|---|---|---|
| participants result in appropriate comparison groups? | | 4185 individuals were initially enrolled, and 3651 participants were retained for final analysis after applying objective exclusion criteria. Although the study population was not randomly sampled from the general population, the use of a structured recruitment process and transparent criteria reduces the risk of systematic differences between included and excluded individuals. Therefore, the risk of selection bias is considered probably low. |
| **CONFLICT OF INTEREST** | Definitely low | “The authors declare that they have no known competing financial interests or personal relationships that could have appeared to influence the work reported in this paper.” |

#### eTable 1. Joanna Briggs Institute (JBI) checklist (applied to cross-sectional studies)

| First Author | Publication Year | Were the criteria for inclusion in the sample clearly defined? | Were the study subjects and the setting described in detail? | Was the exposure measured in a valid and reliable way? | Were objective, standard criteria used for measurement of the condition? | Were confounding factors identified? | Were strategies to deal with confounding factors stated? | Were the outcomes measured in a valid and reliable way? | Was appropriate statistical analysis used? |
|---|---|---|---|---|---|---|---|---|---|
| VoPham et al. | 2022 | Unclear | Yes | Yes | Yes | Yes | Yes | Yes | Yes |
| Guo et al. | 2022 | Yes | Yes | Yes | Yes | Yes | Yes | Yes | Yes |
| Ji et al. | 2024 | Yes | Yes | Yes | Yes | Yes | Yes | Yes | Yes |
| Bo et al. | 2024 | Yes | Yes | Yes | yes | Yes | Yes | Yes | Yes |
| Guo et al. | 2023 | Yes | Yes | Yes | Yes | Yes | Yes | Yes | Yes |
| Matthiessen et al. | 2023 | Yes | Yes | Yes | Yes | Yes | Yes | Yes | Yes |
| Cheng et al. | 2024 | Yes | Yes | Yes | Yes | Yes | Yes | Yes | Yes |
| Patterson et al. | 2023 | Yes | Yes | Yes | Yes | Yes | Yes | Yes | Yes |
| Yun et al. | 2025 | Yes | Yes | Yes | Yes | Yes | Yes | Yes | Yes |
| Wu et al. | 2023 | Yes | Yes | Yes | Yes | Yes | Yes | Yes | Yes |
| Zhang et al. | 2023 | Yes | Yes | Yes | Yes | Yes | Yes | Yes | Yes |
| Cheng et al. | 2023 | Yes | Yes | Yes | yes | Yes | Unclear | Yes | Yes |
| Zhang et al. | 2024 | Yes | Yes | Yes | yes | Yes | Unclear | Yes | Yes |
| He et al. | 2023 | Yes | Yes | Yes | Yes | Yes | Unclear | Yes | Yes |
| Yang et al. | 2021 | Yes | Yes | Yes | Yes | Yes | Unclear | Yes | Yes |
| Cai et al. | 2021 | Yes | Yes | Yes | Yes | Yes | Yes | Yes | Yes |
| Chen et al. | 2022 | Yes | Yes | Yes | Yes | Yes | Unclear | Yes | Yes |
| Kim et al. | 2019 | Yes | Yes | Yes | Yes | Yes | Unclear | Yes | Yes |
| Peng et al. | 2022 | Yes | Yes | Yes | Yes | Yes | Unclear | Yes | Yes |
| An et al. | 2021 | Yes | Yes | Yes | Yes | Yes | Unclear | Yes | Yes |
| Li et al. | 2020 | Yes | Yes | Yes | Yes | Yes | Unclear | Yes | Yes |
| Maodong et al. | 2023 | Yes | Yes | Yes | Yes | Yes | Unclear | Yes | Yes |
| Park et al. | 2021 | Yes | Yes | Yes | Yes | Yes | Yes | Yes | Yes |
| Fan et al. | 2024 | Yes | Yes | Yes | Yes | Yes | Yes | Yes | Yes |
| Nguyen et al. | 2022 | Yes | Yes | Yes | Yes | Yes | Yes | Yes | Yes |
| Xie et al. | 2023 | Yes | Yes | Yes | Yes | Yes | Unclear | Yes | Yes |
| Yang et al. | 2021 | Yes | Yes | Yes | Yes | Yes | Yes | Yes | Yes |
| Kim et al. | 2023 | Yes | Yes | Yes | Yes | Yes | Unclear | Yes | Yes |
| Wan et al. | 2022 | Yes | Yes | Yes | Yes | Unclear | Unclear | Yes | Yes |
| Ye et al. | 2024 | Yes | Yes | Yes | Yes | Yes | Yes | Yes | Yes |
| Liu et al. | 2024 | Yes | Yes | Yes | Yes | Yes | Yes | Yes | Yes |

**Note:** The JBI checklist includes eight criteria: “inclusion criteria,” “study subjects,” “exposure measurement,” “condition measurement,” “identification of confounding factors,” “adjustment for confounding factors,” “outcome measurement,” and “use of valid statistical analysis methods." Each criterion is evaluated as "Yes," "No," "Unclear," or "Not applicable." In this study, articles with at least six "Yes" ratings were considered high quality.

#### eTable 2. The Newcastle–Ottawa Scale (NOS) checklist (applied to case-control studies)

| First Author | Year | Selection | | | Comparability | | Exposure | | | Study Score |
|---|---|---|---|---|---|---|---|---|---|---|
| | | Representativeness of the cases | Selection of the controls | Is the case definition adequate | Definition of Controls | Comparability of cohorts on the basis of the design or analysis | Assessment of outcome | Same assess methods for cases and control | No-response rate | |
| Wu et al. | 2024 | 1 | 1 | 1 | 1 | 1 | 1 | 1 | 1 | 8 |
| Zhang et al. | 2023 | 1 | 1 | 1 | 1 | 1 | 1 | 1 | 0 | 7 |

#### eTable 3. The Newcastle–Ottawa Scale (NOS) checklist (applied to cohort studies)

| First Author | Year | Selection | | | | Comparability | Exposure | | | Study Score |
|---|---|---|---|---|---|---|---|---|---|---|
| | | Representativeness of the exposed cohort | Selection of the non-exposed cohort | Ascertainment of exposure | Demonstration that outcome of interest was not present at start of study | Comparability of cohorts on the basis of the design or analysis | Assessment of outcome | Was follow-up long enough for outcomes to occur | Adequacy of follow-up of cohorts | |
| Han et al. | 2023 | 1 | 1 | 1 | 1 | 2 | 1 | 1 | 0 | 8 |

**Note:** The NOS assesses three domains: "selection of study groups" (maximum 5 points), "comparability of study groups" (maximum 2 points), and "ascertainment of exposure and outcomes" (maximum 3 points). Studies scoring more than 6 points were often considered of moderate-to-high quality and are included in our meta-analyses.

#### eTable 4. Risk of bias for included studies

| | Selection Bias | Confounding Bias | Attrition/Exclusion Bias | Detection bias: Exposure Characterization | Detection Bias: Outcome Characterization | Selective Reporting Bias | Conflict of interest | Summary Tiered classification |
|---|---|---|---|---|---|---|---|---|
| VoPham et al.2022 | + | + | - | + | + | + | ++ | **T2** |
| Guo et al.2022 | + | + | + | + | ++ | + | ++ | **T1** |
| Ji et al.2024 | + | + | + | + | ++ | + | ++ | **T1** |
| Han et al.2023 | + | ++ | ++ | + | ++ | + | ++ | **T1** |
| Bo et al.2024 | + | ++ | + | + | + | + | ++ | **T1** |
| Guo et al.2023 | + | + | + | + | ++ | + | ++ | **T1** |
| Matthiessen et al.2023 | + | + | + | + | + | + | ++ | **T1** |
| Cheng et al.2024 | + | + | + | + | ++ | + | ++ | **T1** |
| Patterson et al.2023 | + | + | + | + | ++ | + | ++ | **T1** |
| Kim et al.2019 | + | + | + | ++ | + | + | ++ | **T1** |
| Peng et al.2023 | + | + | + | ++ | + | + | ++ | **T1** |
| An et al.2021 | + | + | + | ++ | + | + | ++ | **T1** |
| He et al.2023 | + | + | + | ++ | ++ | + | ++ | **T1** |
| Yang et al.2021 | + | + | + | ++ | + | + | ++ | **T1** |
| Cai et al.2021 | + | ++ | + | ++ | + | + | ++ | **T1** |
| Chen et al.2022 | + | + | + | ++ | ++ | + | ++ | **T1** |
| Yun et al.2025 | + | + | + | ++ | + | + | ++ | **T1** |
| Wu et al.2024 | + | + | ++ | ++ | ++ | + | ++ | **T1** |
| Wu et al.2023 | + | + | + | ++ | + | + | ++ | **T1** |
| Zhang et al.2023 | + | ++ | + | ++ | ++ | + | - | **T2** |
| Cheng et al.2023 | + | + | + | ++ | ++ | + | ++ | **T1** |
| Zhang et al.2024 | + | + | + | ++ | ++ | + | ++ | **T1** |
| Li et al.2020 | + | + | + | ++ | ++ | + | ++ | **T1** |
| Zhang et al.2023 | + | ++ | + | ++ | ++ | + | ++ | **T1** |
| Park et al.2021 | + | ++ | + | ++ | + | + | ++ | **T1** |
| Maodong et al.2023 | + | + | + | ++ | + | + | ++ | **T1** |
| Fan et al.2024 | + | ++ | + | ++ | + | + | ++ | **T1** |
| Nguyen et al.2022 | + | ++ | + | ++ | + | + | ++ | **T1** |
| Xie et al.2023 | + | + | + | ++ | + | + | ++ | **T1** |
| Yang et al.2021 | + | ++ | + | + | + | + | ++ | **T1** |
| Kim et al.2023 | + | + | + | ++ | + | + | ++ | **T1** |
| Wan et al.2022 | + | - | + | ++ | ++ | + | ++ | **T2** |
| Ye et al.2024 | + | ++ | + | ++ | + | + | ++ | **T1** |
| Liu et al.2024 | + | ++ | + | ++ | ++ | + | ++ | **T1** |

**Personal Levels**

| | |
|---|---|
| ++ | Definitely low risk of bias |
| + | Probably low risk of bias |

| | |
|---|---|
| - | Probably high risk of bias |
| -- | Definitely high risk of bias |

#### eTable 5. Confidence in the level of evidence

| | | Rate | Comments |
|---|---|---|---|
| Initials rate of confidence | | Low to Moderate confidence | Total 34 studies, most of them (31)were cross-sectional studies, 2 case-control and 1 cohort. |
| Downgrading factors | Risk of bias | No downgrade | Most individual studies were classified in the Tier 1 (n = 31) and Tier 2 (n = 3). None of studies was classified in Tier 3. |
| | Unexplanied inconsistency | No downgrade | Although heterogeneity in the some studies, further explanations(such as subgroups analysis, sensitivity analysis and publication bias))were perfomed to explore the origin of heterogenity. |
| | Indirectness | No downgrade | No evidence of lack of applicability of populations or study design was found across studies. |
| | Imprecision | No downgrade | The confidence intervals were not considered especially concerning to penalize the confidence. |
| | Publication bias | No downgrade | Despite limited studies included, no additional evidence of publication bias was found (e.g. private sponsorship, unpublished studies) |
| Upgrading factors | Large magnitude | No upgrade | Insufficiently to upgrade. |
| | Dose-response | No upgrade | The result of air pollutants revealed potential Dose-resonse gradient, but insufiicient to upgrade the rating. |
| | Residual confounding | No upgrade | No evidence of confounding that would bias toward null. |
| | Consistency | No upgrade | The consistency was considered to be moderate and insufficient to upgrade the rating |
| Final rate of confidence | | Low to Moderate confidence | |
| Level of evidence | | Low to Moderate | |

#### eTable 6. Estimation of publication bias

| Pollutant | Study Number | Egger's test | | | |
|---|---|---|---|---|---|
| | | P value | t | SE | df |
| PM2.5 | 4 | 0.4538 | 0.92 | 3.454 | 2 |
| PM10 | 3 | 0.5447 | 0.87 | 3.8785 | 1 |
| PM1 | 3 | 0.4817 | 1.06 | 8.0844 | 1 |
| BPA | 3 | 0.7268 | 0.46 | 1.8324 | 1 |
| MECPP | 4 | 0.2204 | 1.76 | 1.6232 | 2 |
| MEHHP | 4 | 0.2117 | 1.81 | 0.9912 | 2 |
| MEOHP | 4 | 0.7809 | 0.32 | 2.1004 | 2 |
| PFDeA | 4 | 0.7679 | 0.34 | 6.6335 | 2 |
| PFHxS | 4 | 0.6185 | -0.58 | 2.0218 | 2 |
| PFNA | 6 | 0.7140 | 0.39 | 3.4069 | 4 |
| PFOA | 6 | 0.7735 | 0.31 | 1.1557 | 4 |
| PFOS | 6 | 0.8815 | -0.16 | 3.5171 | 4 |
| Pb | 3 | 0.5304 | 0.91 | 0.9415 | 1 |
| Cd | 3 | 0.3109 | 1.88 | 2.0182 | 1 |
| Hg | 3 | 0.1496 | -4.18 | 1.0138 | 1 |
| Ba | 3 | 0.8862 | 0.18 | 12.9502 | 1 |
| As | 3 | 0.1734 | -3.58 | 1.2885 | 1 |

**Note**: Egger's test was conducted to assess the presence of publication bias across the included pollutants. The p-values for most pollutants exceed the conventional threshold of 0.05, indicating no strong evidence of publication bias. The t-values represent the test statistic from Egger’s regression, where a larger absolute t-value suggests a stronger bias if significant. The standard errors (SE) indicate the variability of the regression coefficient, with larger SE values reflecting greater uncertainty in bias estimation. The t-values and SE vary considerably across pollutants, reflecting differences in study heterogeneity and sample size. The degrees of freedom (df) determined the statistical power of the test. Since most pollutants have df = 1 or 2, the test’s power to detect bias is limited due to the small number of included studies. Given the small number of studies included for each pollutant, statistical power to detect publication bias may be limited.

#### eFigure 1. Forest plot ORs (95% CI) for the association between PM2.5 per 1ug/m3 increase and per SD increase and SLD.

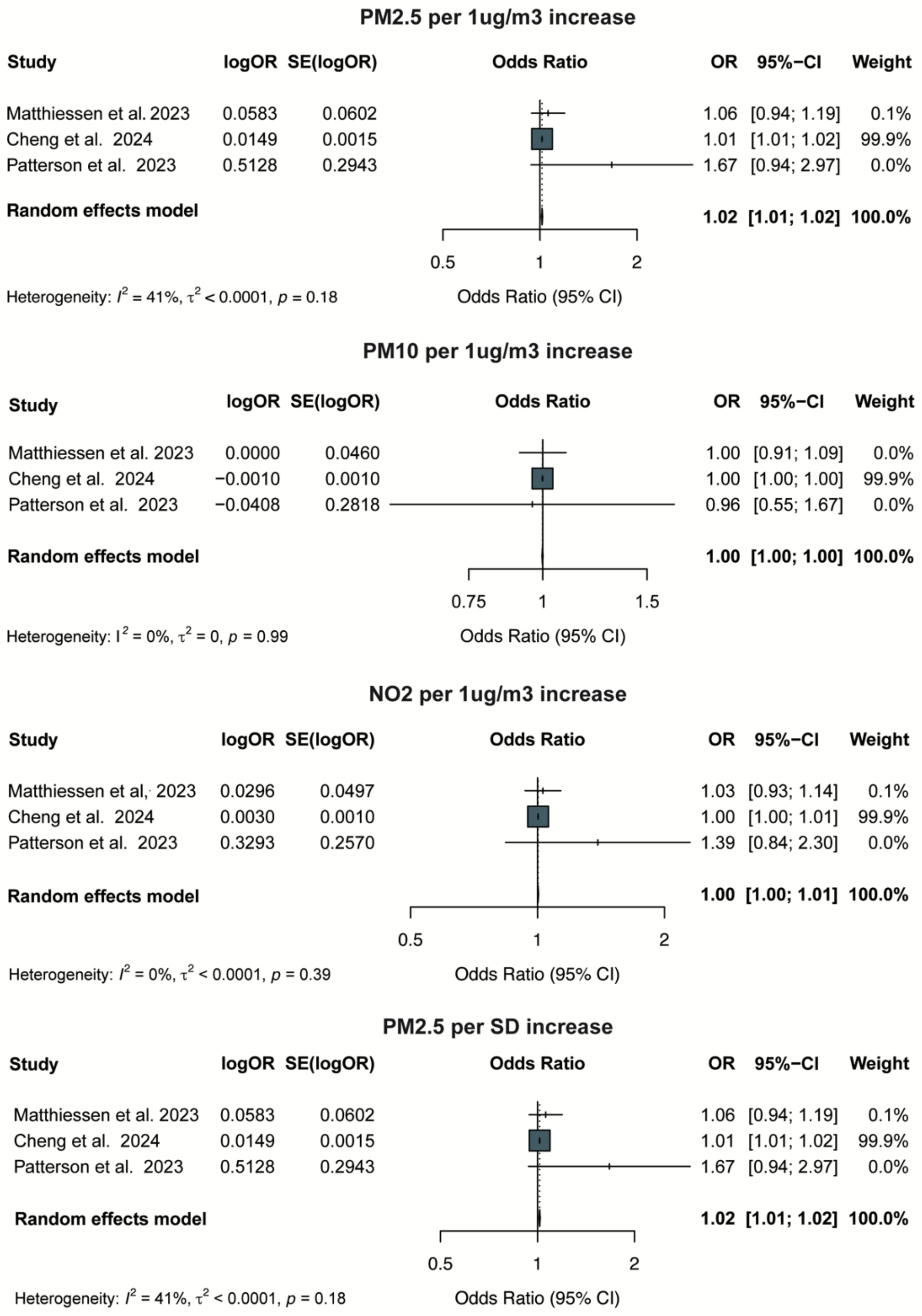


**Note:** The pooled odds ratio (OR) for PM2.5 per 1 μg/m³ and per SD increase indicated a significant association with increased SLD risk, while PM10 and NO2 per 1 μg/m³ increase showed no significant associations.

**eFigure 2. Forest plot ORs (95% CI) for the association between Bisphenol A (BPA) and SLD.**

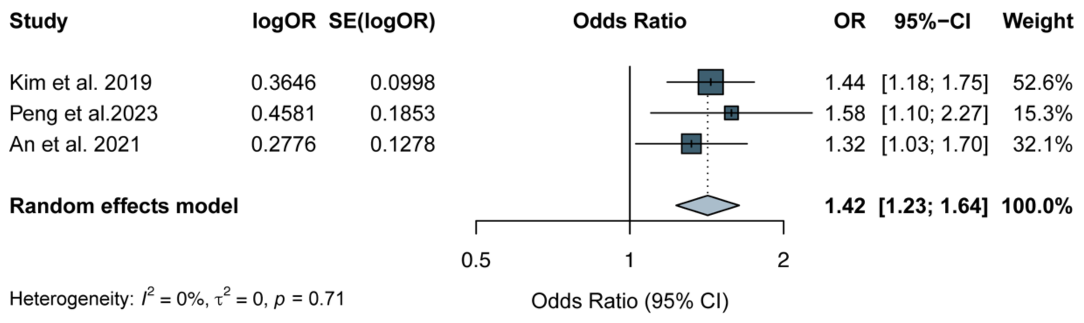

**eFigure 3. Forest plot ORs (95% CI) for the association between perfluoroalkyl substance (PFAS) exposure and SLD.**

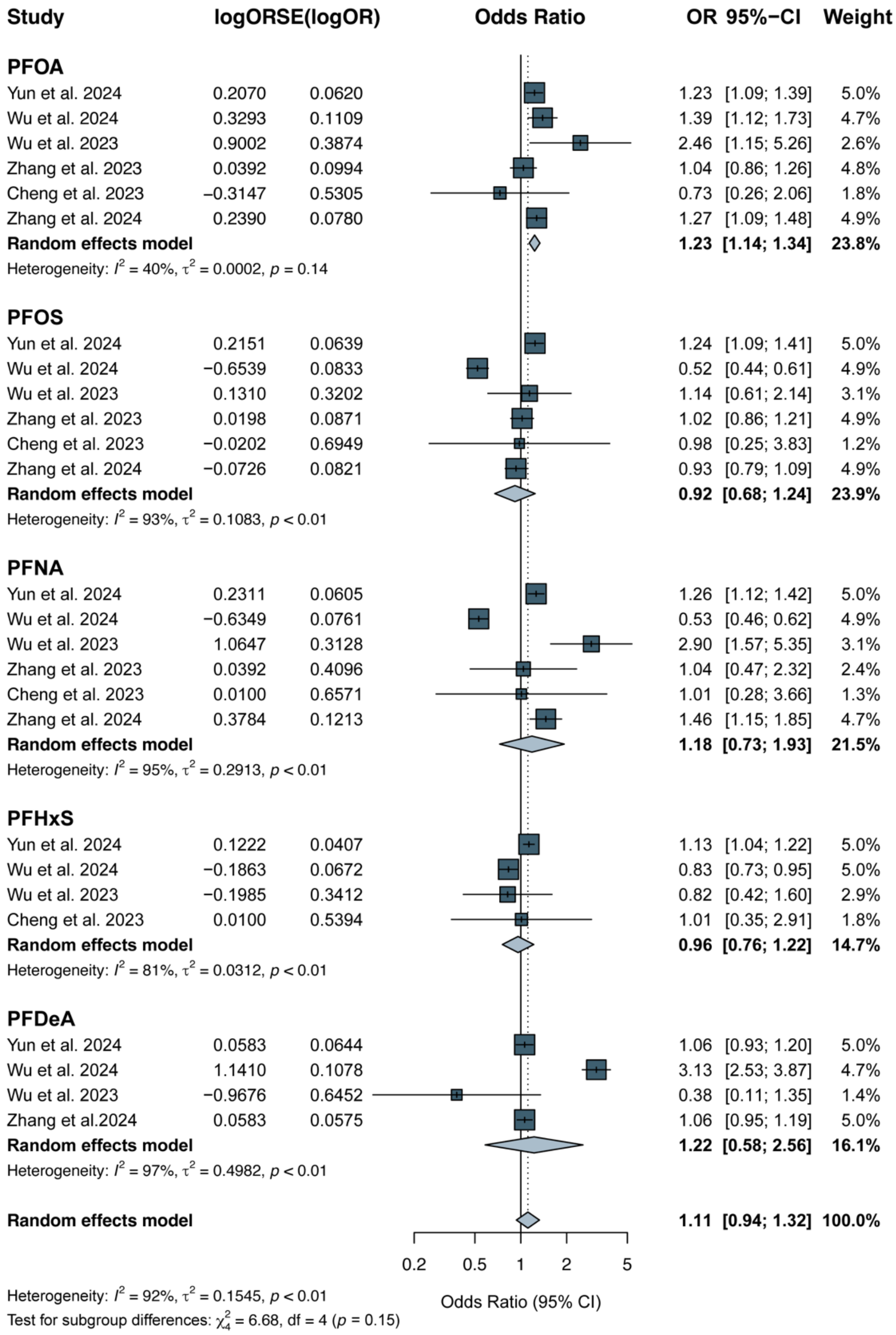

**eFigure 4. Forest plot ORs (95% CI) for the association between phthalate esters (PAEs)exposure and SLD**

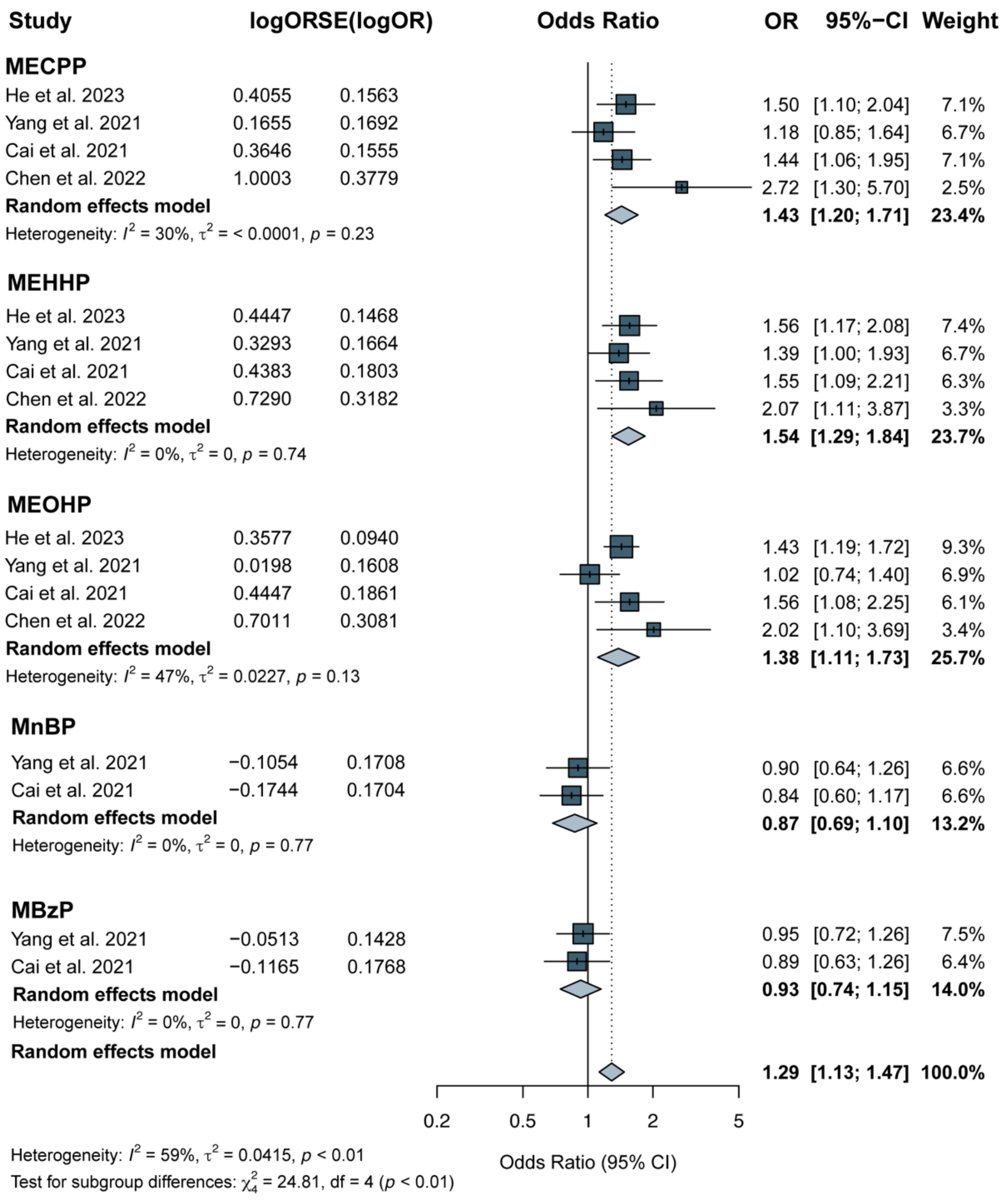

#### eFigure 5. Subgroup Analysis of MEOHP and SLD Risk Based on Adjustment for BMI

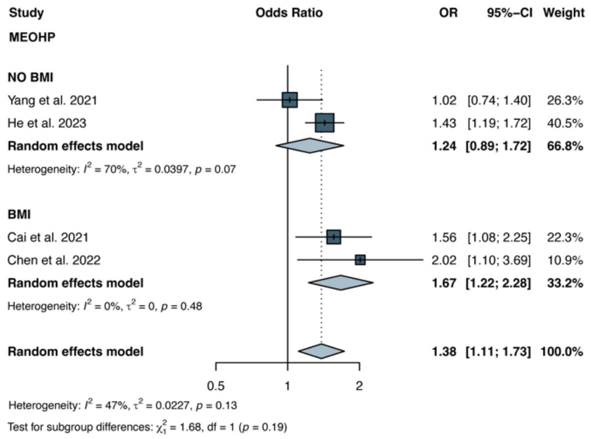


**Note:** Subgroup analysis was conducted for MEOHP to examine whether adjustment for body mass index (BMI) influenced heterogeneity in the effect estimates. The overall heterogeneity was moderate ($I^2$ = 47%, $\tau^2$ = 0.0227). In the subgroup without BMI adjustment, heterogeneity was substantial ($I^2$ = 70%, $\tau^2$ = 0.0397), while in the BMI-adjusted subgroup, heterogeneity was absent ($I^2$ = 0%, $\tau^2$ = 0).

**eFigure 6. Subgroup Analysis of PFAS Exposure and SLD Risk Based on Diagnostic Methods**

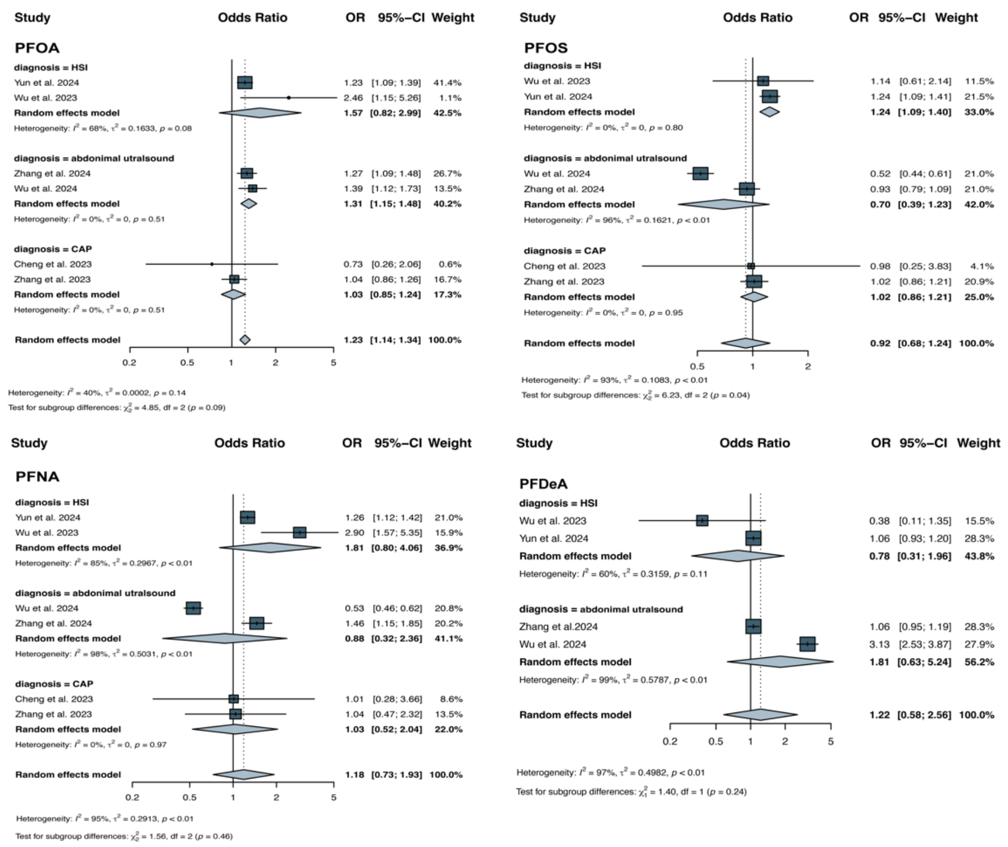


**Note:** Subgroup analyses were conducted for PFOA, PFOS, PFNA, and PFDeA to assess whether differences in steatotic liver disease (SLD) diagnostic methods contributed to heterogeneity in effect estimates. Diagnostic approaches included hepatic steatosis index (HSI), ultrasound, and controlled attenuation parameter (CAP). Compared to the overall pooled analysis, heterogeneity varied substantially across diagnostic subgroups. For PFOA, overall heterogeneity was moderate ($I^2 = 40\%$, $\tau^2 = 0.0002$), and decreased in the ultrasound ($I^2 = 0\%$, $\tau^2 = 0$) and CAP ($I^2 = 0\%$, $\tau^2 = 0$) subgroups, whereas the HSI subgroup showed higher heterogeneity ($I^2 = 68\%$, $\tau^2 = 0.1633$). For PFOS, overall heterogeneity was high ($I^2 = 93\%$, $\tau^2 = 0.1083$) and appeared to be primarily driven by the ultrasound subgroup ($I^2 = 96\%$, $\tau^2 = 0.1621$), while heterogeneity was absent in the HSI and CAP groups ($I^2 = 0\%$, $\tau^2 = 0$). For PFNA, heterogeneity in the overall analysis was substantial ($I^2 = 95\%$, $\tau^2 = 0.2913$), with the ultrasound subgroup showing the highest heterogeneity ($I^2 = 98\%$, $\tau^2 = 0.5031$), while the HSI subgroup had lower but still considerable heterogeneity ($I^2 = 85\%$, $\tau^2 = 0.2967$), and CAP remained low ($I^2 = 0\%$, $\tau^2 = 0$). Similarly, for PFDeA, overall heterogeneity was extremely high ($I^2 = 97\%$, $\tau^2 = 0.4982$), driven largely by the ultrasound subgroup ($I^2 = 99\%$, $\tau^2 = 0.5787$), while heterogeneity in the HSI group was relatively lower ($I^2 = 60\%$, $\tau^2 = 0.3150$).

#### eFigure 7. Subgroup Analysis of PFAS Exposure and SLD Risk Based on Adjustment for diabetes

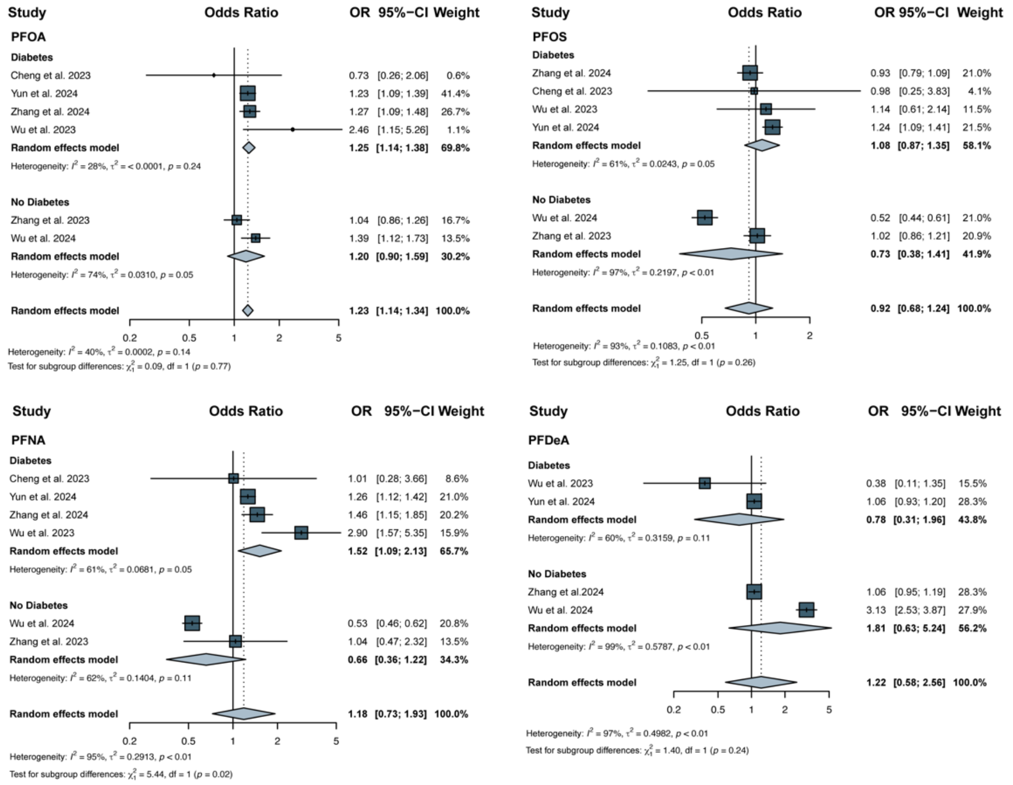


**Note:** Subgroup analyses were conducted for PFOA, PFOS, PFNA, and PFDeA to evaluate whether adjustment for diabetes influenced heterogeneity in the association between PFAS exposure and steatotic liver disease (SLD). Compared to the overall pooled analysis, heterogeneity was reduced in all compounds when restricted to diabetes-adjusted models. For PFOA, heterogeneity decreased from $I^2 = 40\%$, $\tau^2 = 0.0002$ to $I^2 = 28\%$, $\tau^2 < 0.0001$; for PFOS, from $I^2 = 93\%$, $= 0.1083$ to $I^2 = 61\%$, $\tau^2 = 0.0243$; for PFNA, from $I^2 = 95\%$, $\tau^2 = 0.2913$ to $I^2 = 61\%$, $\tau^2 = 0.0681$; and for PFDeA, from $I^2 = 97\%$, $\tau^2 = 0.4982$ to $I^2 = 60\%$, $\tau^2 = 0.3159$. These findings suggest that diabetes may act as a potential source of heterogeneity in PFAS-related SLD risk estimates.

#### eFigure 8. Subgroup Analysis of PFAS Exposure and SLD Risk Based on Adjustment for BMI

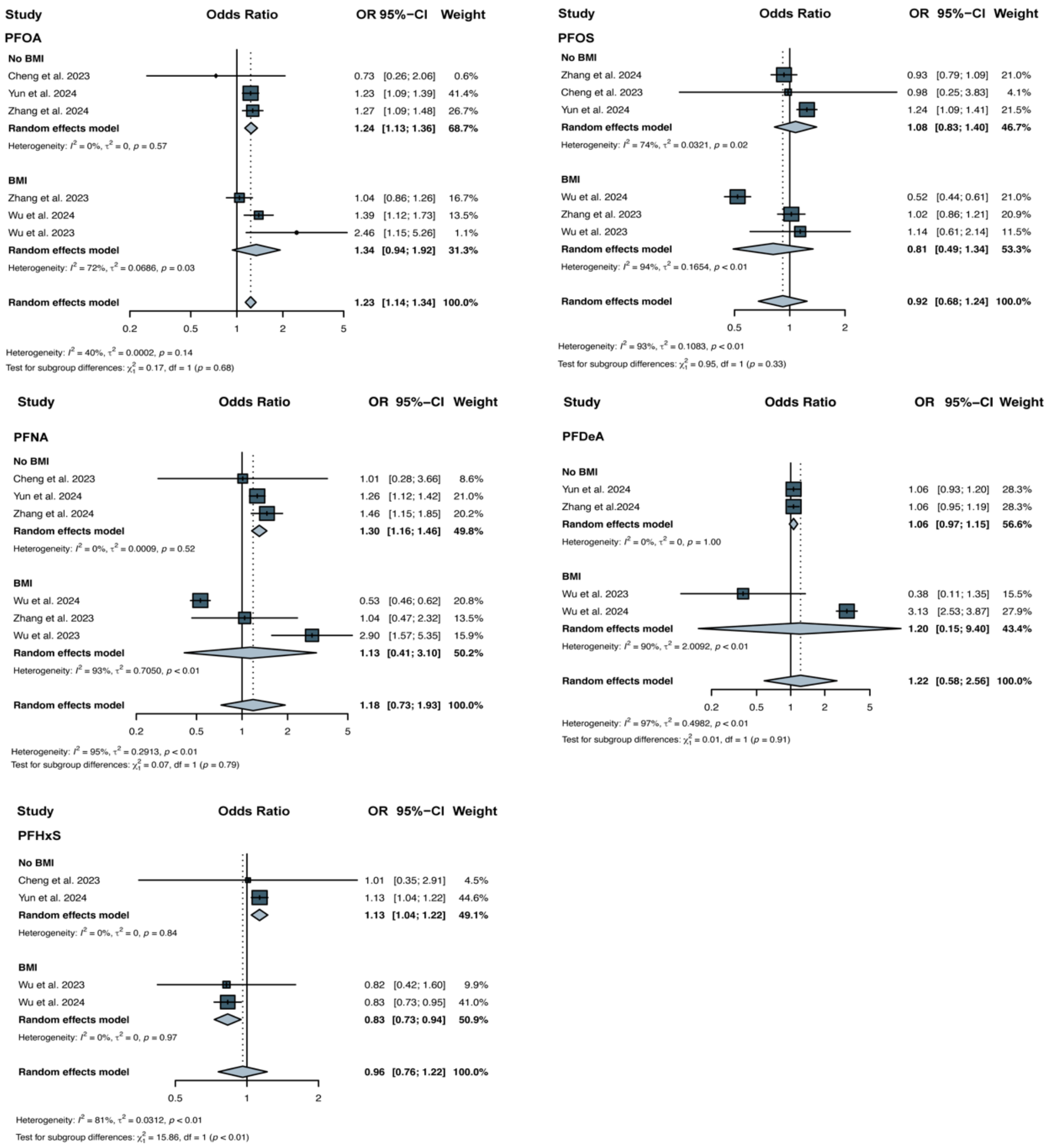


**Note:** Subgroup analyses were conducted for PFOA, PFOS, PFNA, PFDeA, and PFHxS to evaluate whether adjustment for body mass index (BMI) contributed to heterogeneity in effect estimates. For PFOA, the overall heterogeneity was moderate ($I^2$ = 40%, $\tau^2$ = 0.0002), and the BMI-adjusted subgroup showed slightly higher heterogeneity ($I^2$ = 72%, $\tau^2$ = 0.0686) compared to the unadjusted subgroup ($I^2$ = 0%, $\tau^2$ = 0). For PFOS, overall heterogeneity was high ($I^2$ = 93%, $\tau^2$ = 0.1083), primarily driven by the BMI-adjusted group ($I^2$ = 94%, $\tau^2$ = 0.1654), while the unadjusted group showed relatively low heterogeneity ($I^2$ = 74%, $\tau^2$ = 0.0321). For PFNA, substantial heterogeneity was observed overall ($I^2$ = 95%, $\tau^2$ = 0.2913), with greater heterogeneity in the BMI-adjusted subgroup ($I^2$ = 93%, $\tau^2$ = 0.7050) than in the unadjusted group ($I^2$ = 0%, $\tau^2$ =0.0009). For PFDeA, heterogeneity was again high overall ($I^2$ = 97%, $\tau^2$ = 0.4982), driven largely by the BMI-adjusted group ($I^2$ = 90%, $\tau^2$ = 2.0092), while the unadjusted subgroup showed no heterogeneity ($I^2$ = 0%, $\tau^2$ = 0). In contrast, for PFHxS, overall heterogeneity was modest ($I^2$ = 81%, $\tau^2$ = 0.0312), but both BMI-adjusted and unadjusted subgroups showed minimal to no heterogeneity ($I^2$ = 0%, $\tau^2$ = 0).

#### eFigure 9. Subgroup Analysis of PM2.5 and SLD Risk Based on Adjustment for Alcohol Consumption

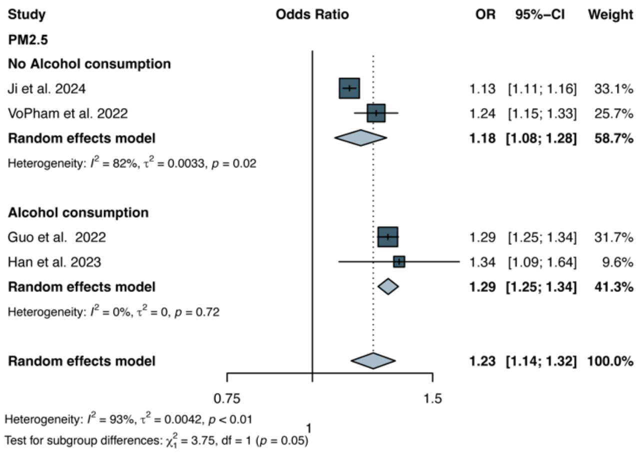


**Note:** Subgroup analysis was conducted for PM2.5 to assess whether adjustment for alcohol consumption influenced heterogeneity in the effect estimates. Overall heterogeneity was high ($I^2$ = 93%, $\tau^2$ = 0.0042, $p < 0.01$). In the subgroup without alcohol adjustment, heterogeneity remained substantial ($I^2$ = 82%, $\tau^2$ = 0.0033, p = 0.02), whereas in the alcohol-adjusted subgroup, heterogeneity was completely eliminated ($I^2$ = 0%, $\tau^2$ = 0, p = 0.72).

#### eFigure 10. Subgroup Analysis of PFAS and SLD Risk Based on Adjustment for Alcohol Consumption

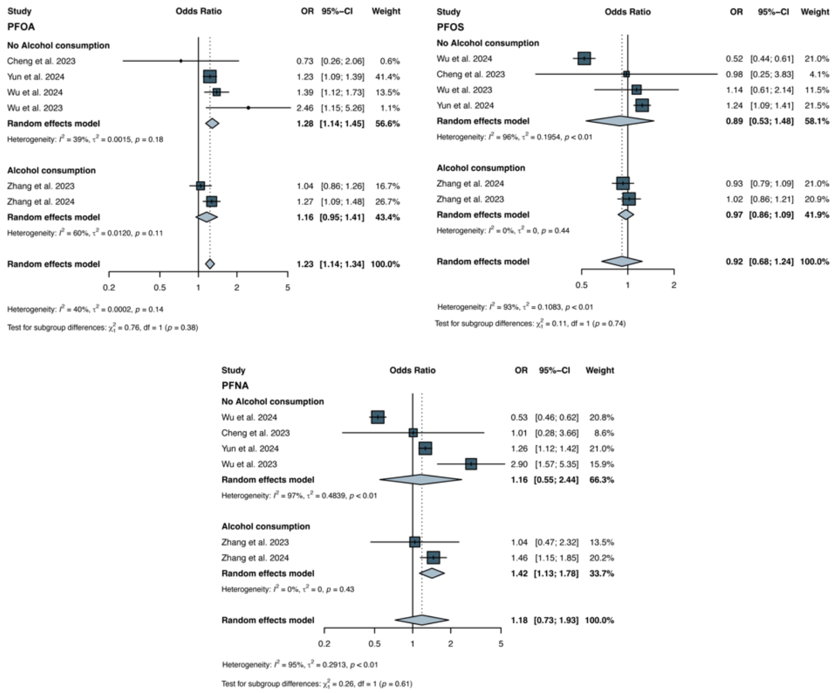


**Note:** Subgroup analyses were conducted for PFOA, PFOS, and PFNA to assess whether adjustment for alcohol consumption contributed to heterogeneity in effect estimates. For PFOA, overall heterogeneity was moderate ($I^2 = 40\%$, $\tau^2 = 0.0002$), with higher heterogeneity observed in the alcohol-adjusted subgroup ($I^2 = 60\%$, $\tau^2 = 0.0120$) than in the unadjusted subgroup ($I^2 = 39\%$, $\tau^2 = 0.0015$). In contrast, for both PFOS and PFNA, alcohol adjustment substantially reduced heterogeneity to zero. For PFOS, the unadjusted subgroup showed high heterogeneity ($I^2 = 96\%$, $\tau^2 = 0.1954$), while the adjusted subgroup had no heterogeneity ($I^2 = 0\%$, $\tau^2 = 0$).

#### eFigure 11. Sensitivity analysis of air pollutants (per 10ug/m3 increase)

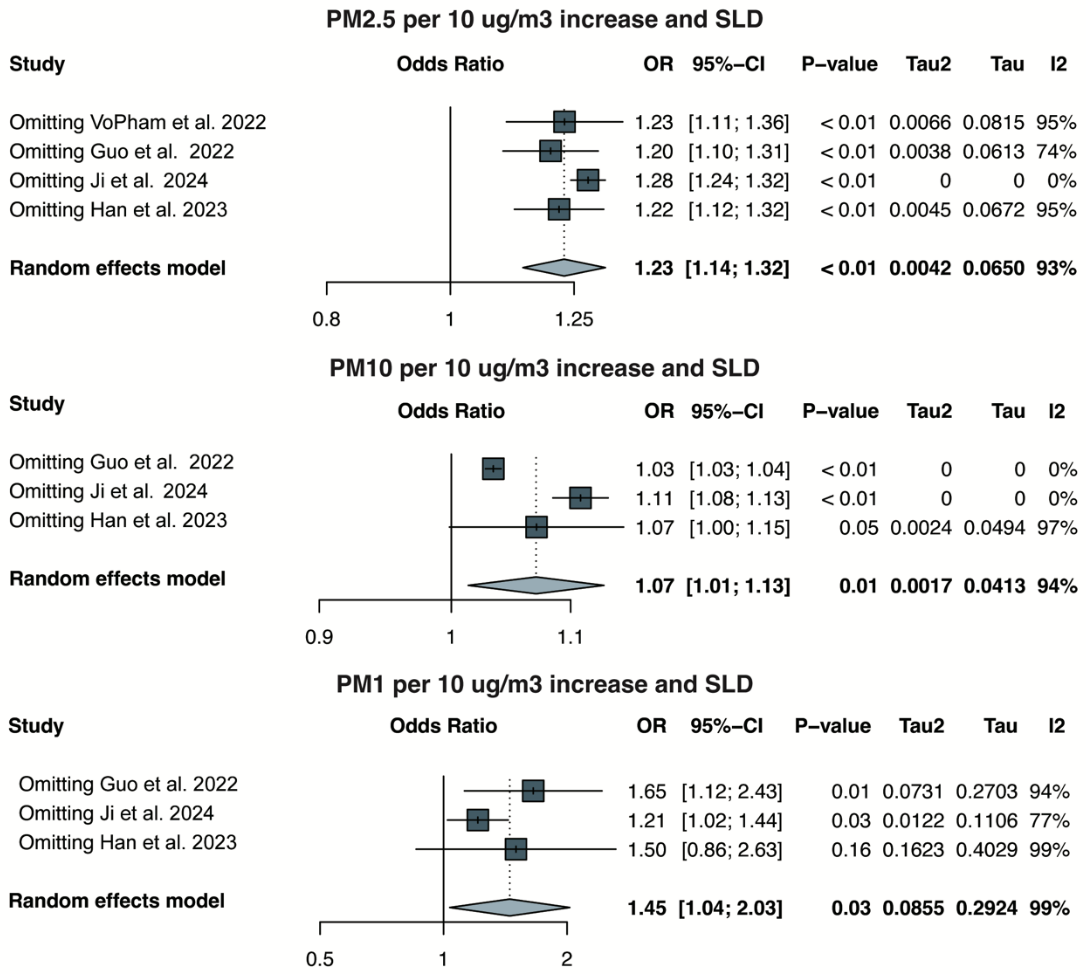


**Note:** For PM2.5, PM10, and PM1, substantial heterogeneity was observed in the overall analysis ($I^2$ = 93%, $\tau^2$ = 0.0042; $I^2$ = 94%, $\tau^2$ = 0.0017; and $I^2$ = 99%, $\tau^2$ = 0.0855, respectively).For PM2.5, excluding Ji et al. (2024) reduced heterogeneity to 0%, and the reanalysis confirmed a robust association between PM2.5 exposure and SLD risk (OR = 1.28, 95% CI: 1.24–1.32). For PM10, excluding either Guo et al. (2022) or Ji et al. (2024) also reduced heterogeneity to 0%. However, exclusion of Guo et al. (2022) produced more consistent results with a narrower confidence interval, further supporting the link between PM10 exposure and SLD. For PM1, removing Ji et al. (2024) reduced heterogeneity from $I^2$ = 99% to 77% and $\tau^2$ from 0.0855 to 0.0122. Although heterogeneity remained moderate, the association remained consistent with the original findings (OR = 1.21, 95% CI: 1.02–1.44).

### eFigure 12. Sensitivity analysis of PFAS

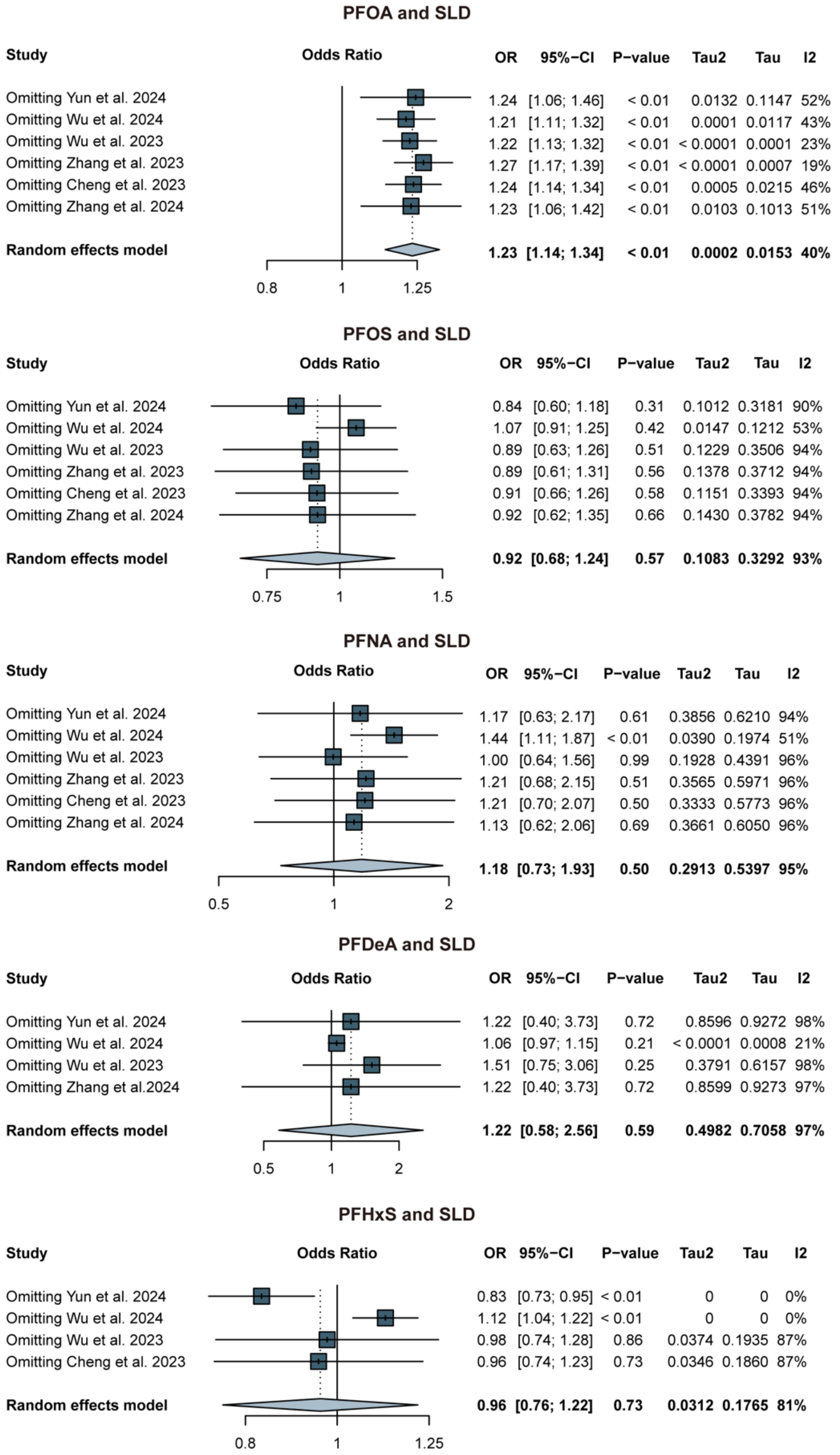


**Note:** For PFOA, which showed moderate heterogeneity ($I^2$ = 40%, $\tau^2$ = 0.0002), sensitivity analysis indicated that excluding Zhang et al. (2023) reduced heterogeneity to $I^2$ = 19%, $\tau^2$ < 0.0001. For PFOS, which exhibited high heterogeneity ($I^2$ = 93%, $\tau^2$ = 0.1083), removing Wu et al. (2024) decreased heterogeneity to $I^2$ = 53%, $\tau^2$ = 0.0147. For PFDeA ($I^2$ = 97%, $\tau^2$ = 0.4982), exclusion of Wu et al. (2024) reduced heterogeneity to $I^2$ = 21%, $\tau^2$ < 0.0001. However, the overall results for PFOA, PFOS, and PFDeA remained nonsignificant after sensitivity analysis. In contrast, for PFNA ($I^2$ = 95%, $\tau^2$ = 0.2913), excluding Wu et al. (2024) reduced heterogeneity to $I^2$ = 51%, $\tau^2$ = 0.0390, and yielded a statistically significant association with SLD (OR = 1.44, 95% CI: 1.11–1.87). For PFHxS ($I^2$ = 81%, $\tau^2$ = 0.0312), removing Yun et al. (2024) reduced heterogeneity to 0% and resulted in a negative association with SLD. Conversely, excluding Wu et al. (2024) also reduced heterogeneity to 0%, but the association turned positive. These inconsistent findings suggest that the association between PFHxS and SLD is not robust and warrants further investigation.

#### eFigure 13. Sensitivity analysis of heavy metals

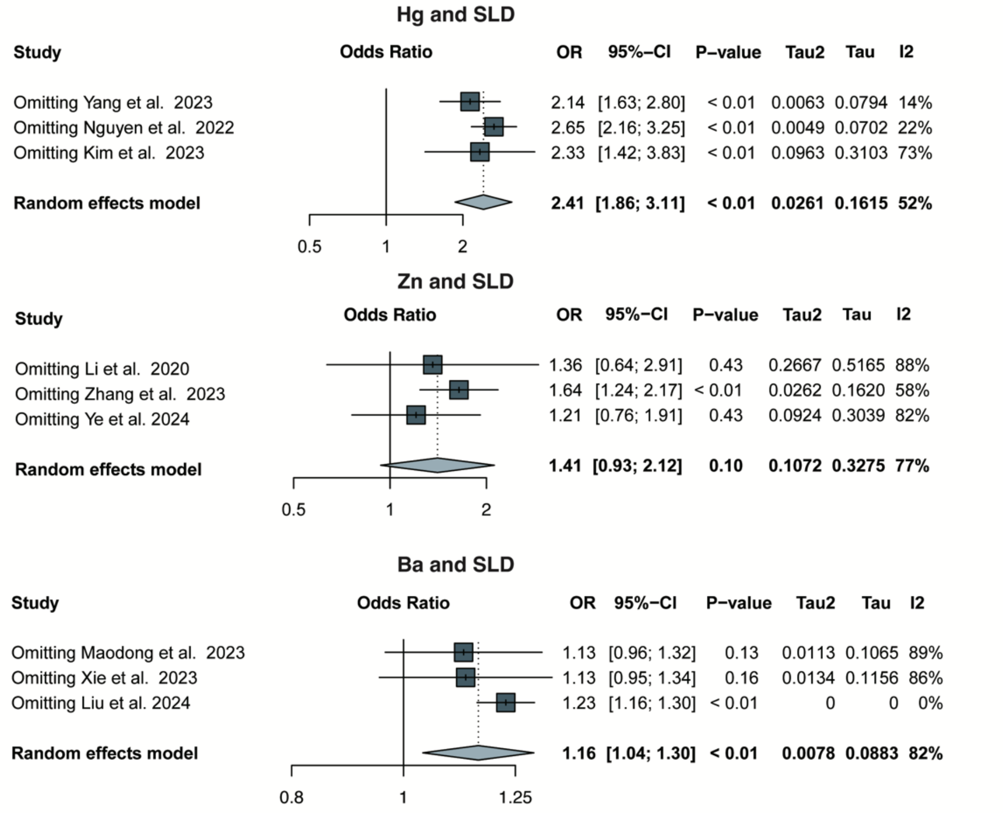


**Note:** For mercury (Hg), which showed moderate heterogeneity ($I^2$ = 52%, $\tau^2$ = 0.0261), sensitivity analysis revealed that excluding Yang et al. (2023) reduced heterogeneity to $I^2$ = 14%, $\tau^2$ = 0.0063, while the association with SLD remained statistically significant (OR = 2.14, 95% CI: 1.63–2.80). For barium (Ba), excluding Liu et al. (2024) reduced heterogeneity to 0%, and the association with SLD remained statistically significant (OR = 1.23, 95% CI: 1.16–1.30). For zinc (Zn), excluding Zhang et al. (2023) decreased heterogeneity from $I^2$ = 77%, $\tau^2$ = 0.1072 to $I^2$ = 58%, $\tau^2$ = 0.0262. Although heterogeneity remained moderate, the direction and significance of the association changed, suggesting that Zn exposure may be positively associated with SLD risk (OR = 1.64, 95% CI: 1.24–2.17).

#### eFigure14. Funnel Plots of Air Pollutants Before and After Trim-and-Fill Adjustment

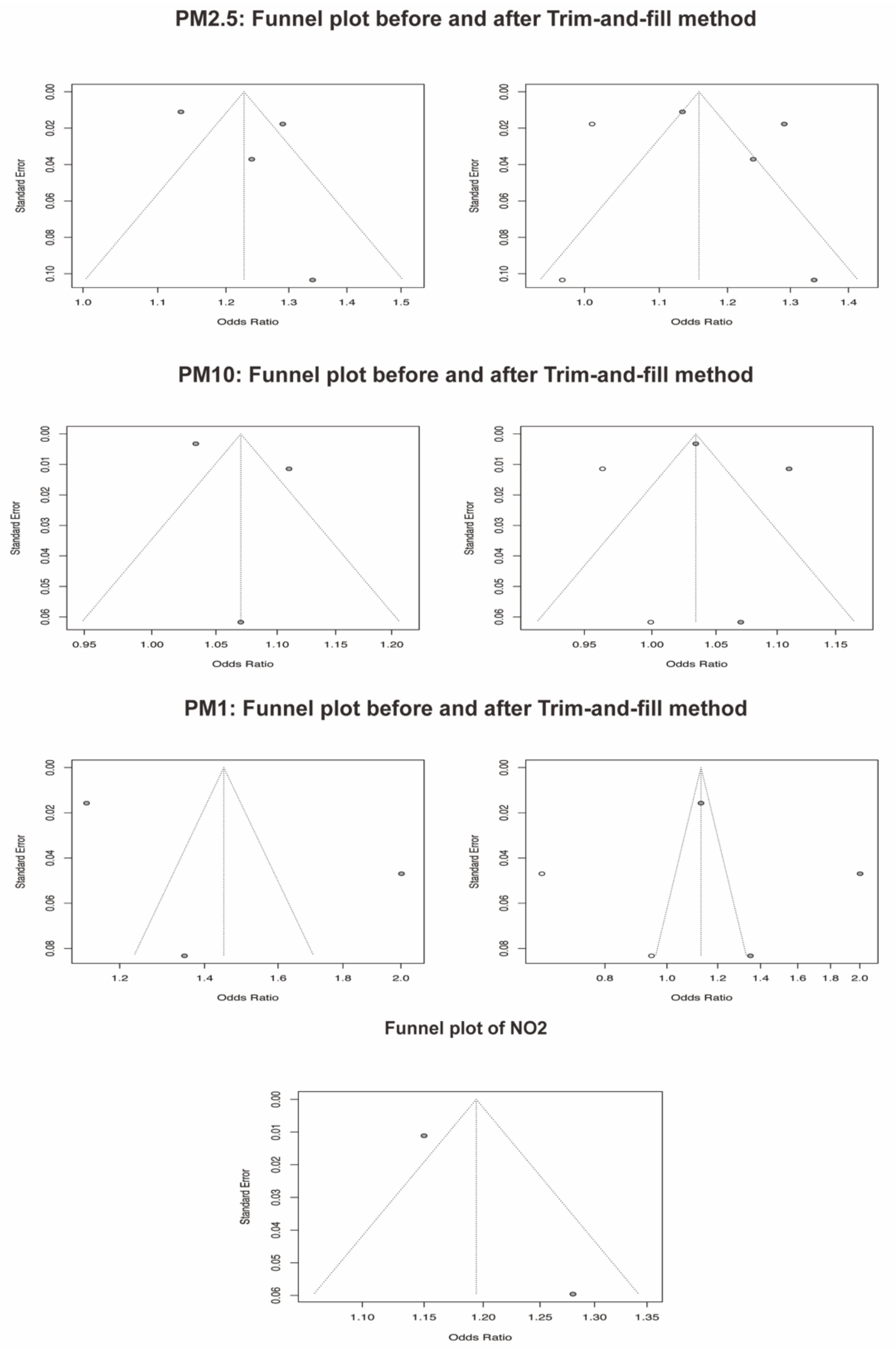


**Note**: Funnel plots were used to assess potential publication bias for air pollutants. For PM2.5, PM10, and PM1, plots before and after trim-and-fill adjustment are shown. Asymmetry was observed in the original plots, and the trim-and-fill method imputed additional studies to correct for potential bias. For $NO_2$, the funnel plot was presented without trim-and-fill adjustment due to the limited number of studies (n < 3).

#### eFigure15. Funnel Plots of PFAS Before and After Trim-and-Fill Adjustment

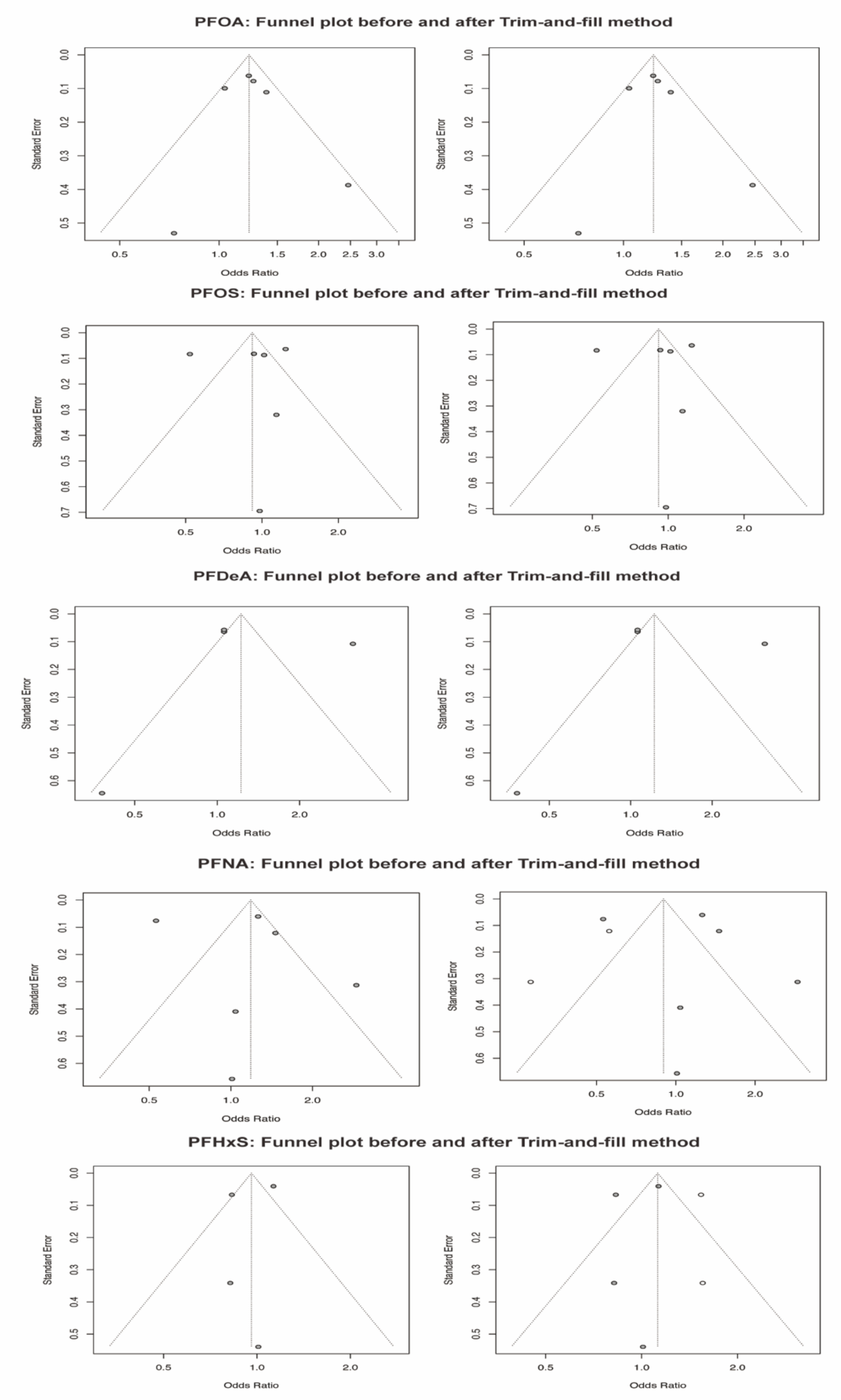


**Note:** Funnel plots were used to evaluate potential publication bias for five PFAS compounds: PFOA, PFOS, PFDeA, PFNA, and PFHxS. For each compound, plots before and after trim-and-fill adjustment are shown. Asymmetry was observed in the plots of PFNA and PFHxS, and the trim-and-fill method imputed additional studies to adjust for potential bias.

#### eFigure16. Funnel Plots of PAEs Before and After Trim-and-Fill Adjustment

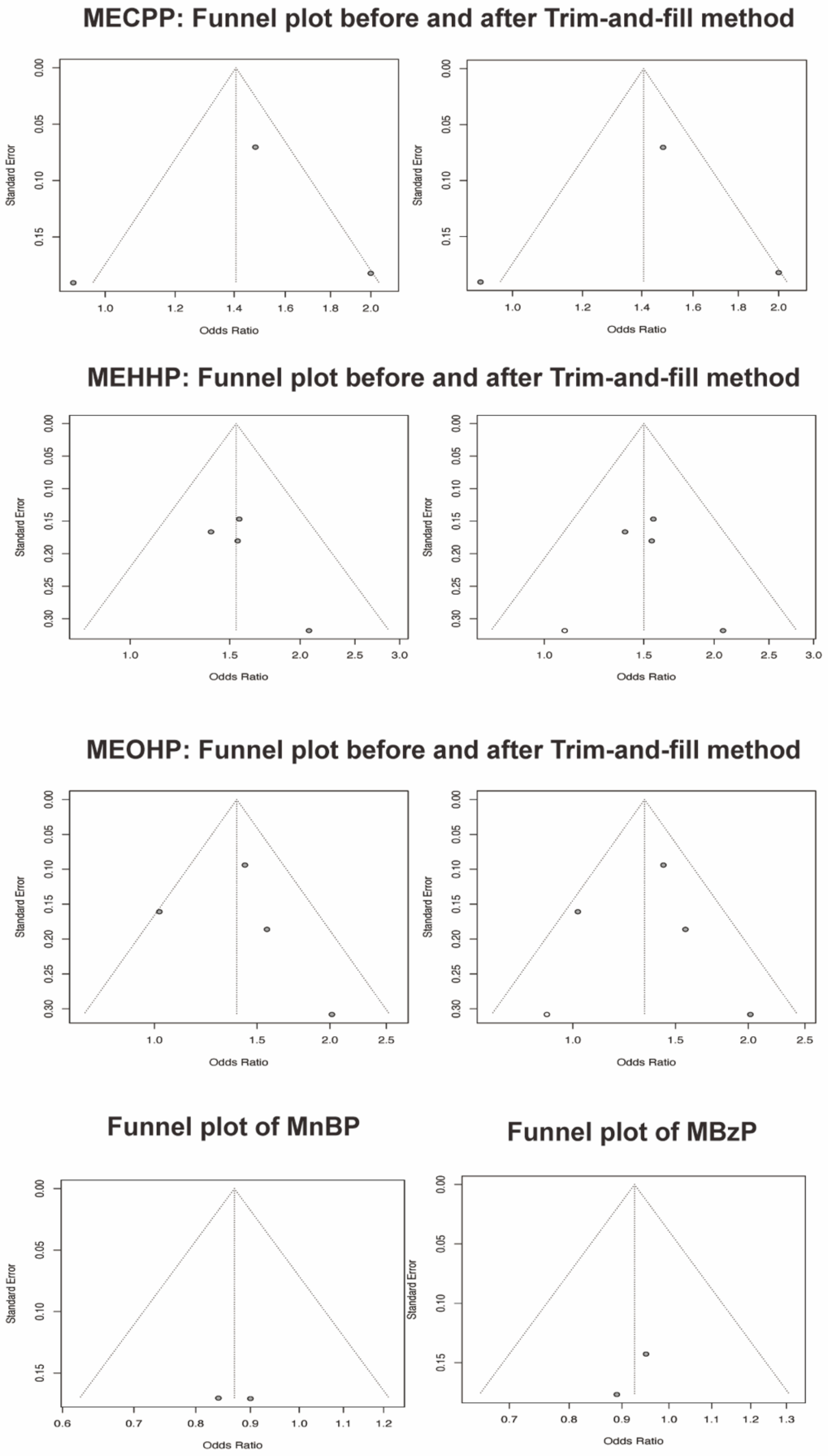


**Note:** Funnel plots were used to assess potential publication bias for five phthalate metabolites. For MECPP, MEHHP, and MEOHP, plots before and after trim-and-fill adjustment are shown. For MEHHP and MEOHP, the trim-and-fill method imputed additional studies where asymmetry was observed, indicating possible publication bias. For MnBP and MBzP, only the original funnel plots are displayed, as the number of included studies was insufficient to reliably perform trim-and-fill correction.

#### eFigure17. Funnel Plot of BPA Before and After Trim-and-Fill Adjustment

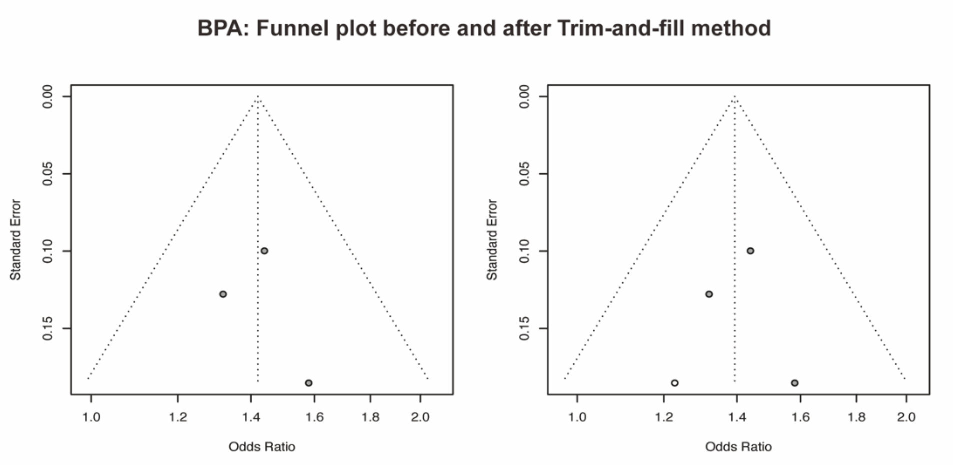


**Note:** Funnel plots were used to evaluate potential publication bias for bisphenol A (BPA). Asymmetry observed in the original plot prompted the imputation of additional studies, suggesting possible publication bias.

#### eFigure18. Funnel Plots of Metal Pollutants (n < 3)

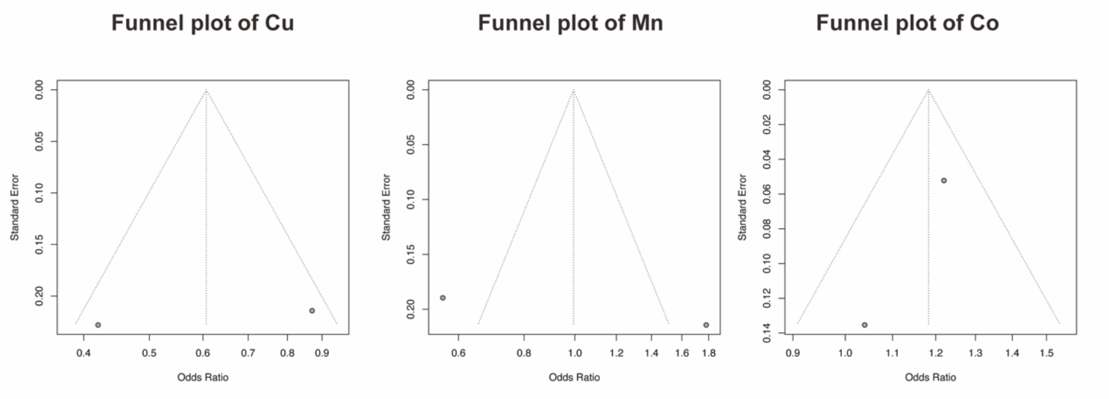


**Note**: Funnel plots were generated for four metal pollutants: copper (Cu), manganese (Mn), and cobalt (Co), each with fewer than three included studies. Due to the limited number of studies, the trim-and-fill method was not applied.

#### eFigure19. Funnel Plots of Metal Pollutants (n ≥ 3)

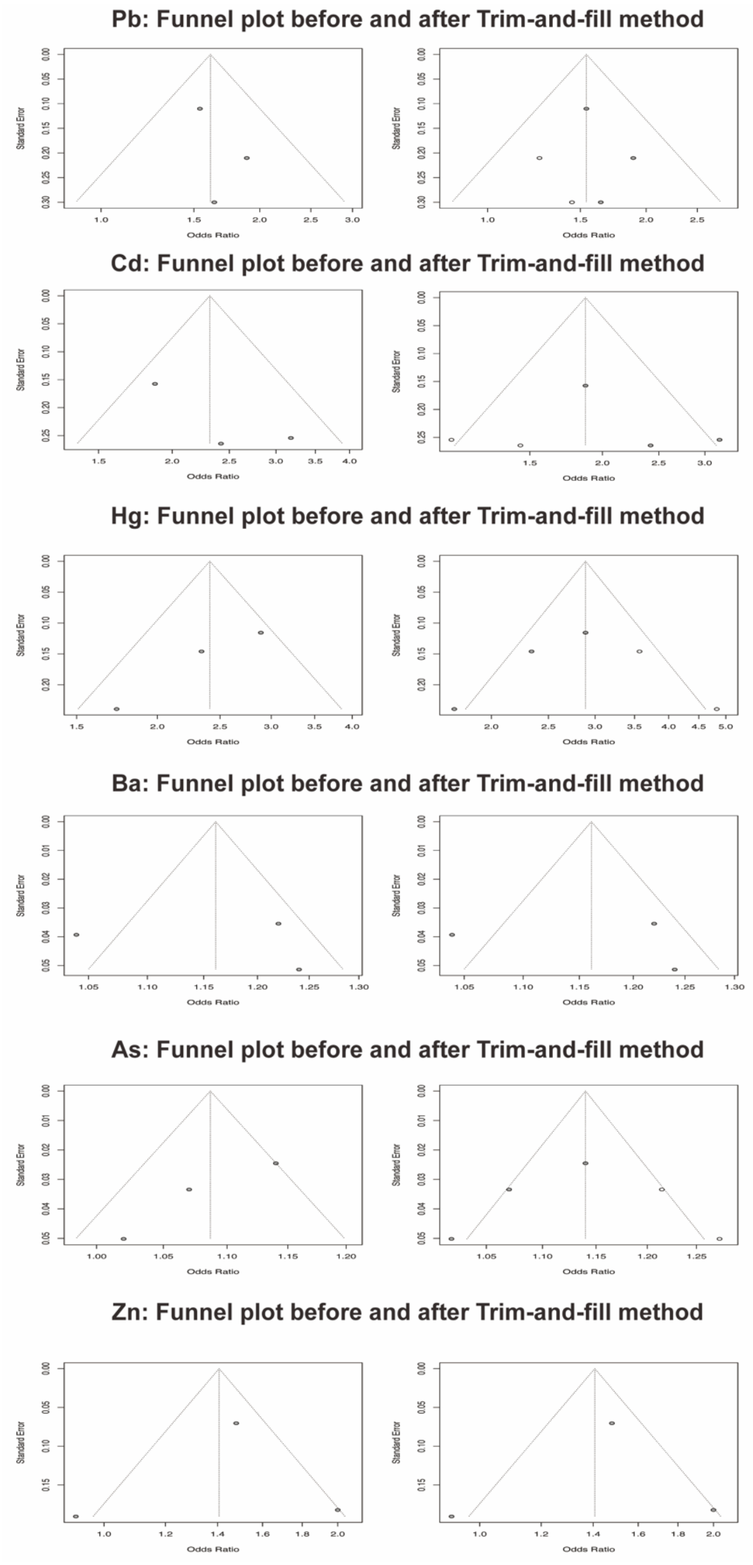


**Note:** Funnel plots were used to assess potential publication bias for six metal pollutants: lead (Pb), cadmium (Cd), mercury (Hg), barium (Ba), arsenic (As), and zinc (Zn). For each pollutant, the left panel shows the original funnel plot, while the right panel displays the adjusted plot after applying the trim-and-fill method. Asymmetry was observed in Pb, Cd, Hg, and As, and additional studies were imputed accordingly, indicating possible publication bias.
