## Supplemental file for "Mapping the association between environmental pollutants and steatotic liver disease: a systematic review and meta-analysis"

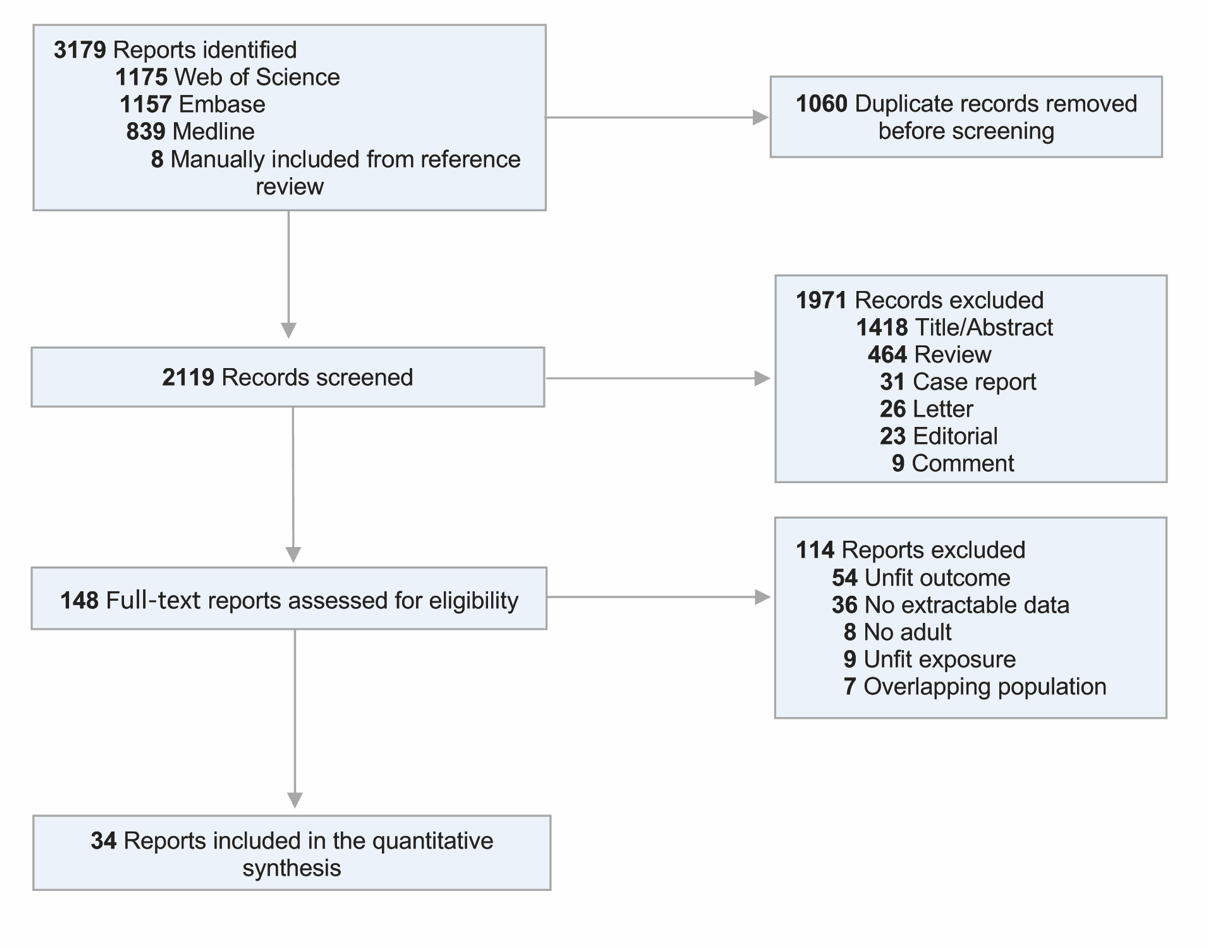
**Figure 1. Flow Diagram of included studies**

Figure 1. The flowchart illustrates the study selection process following the PRISMA guideline.

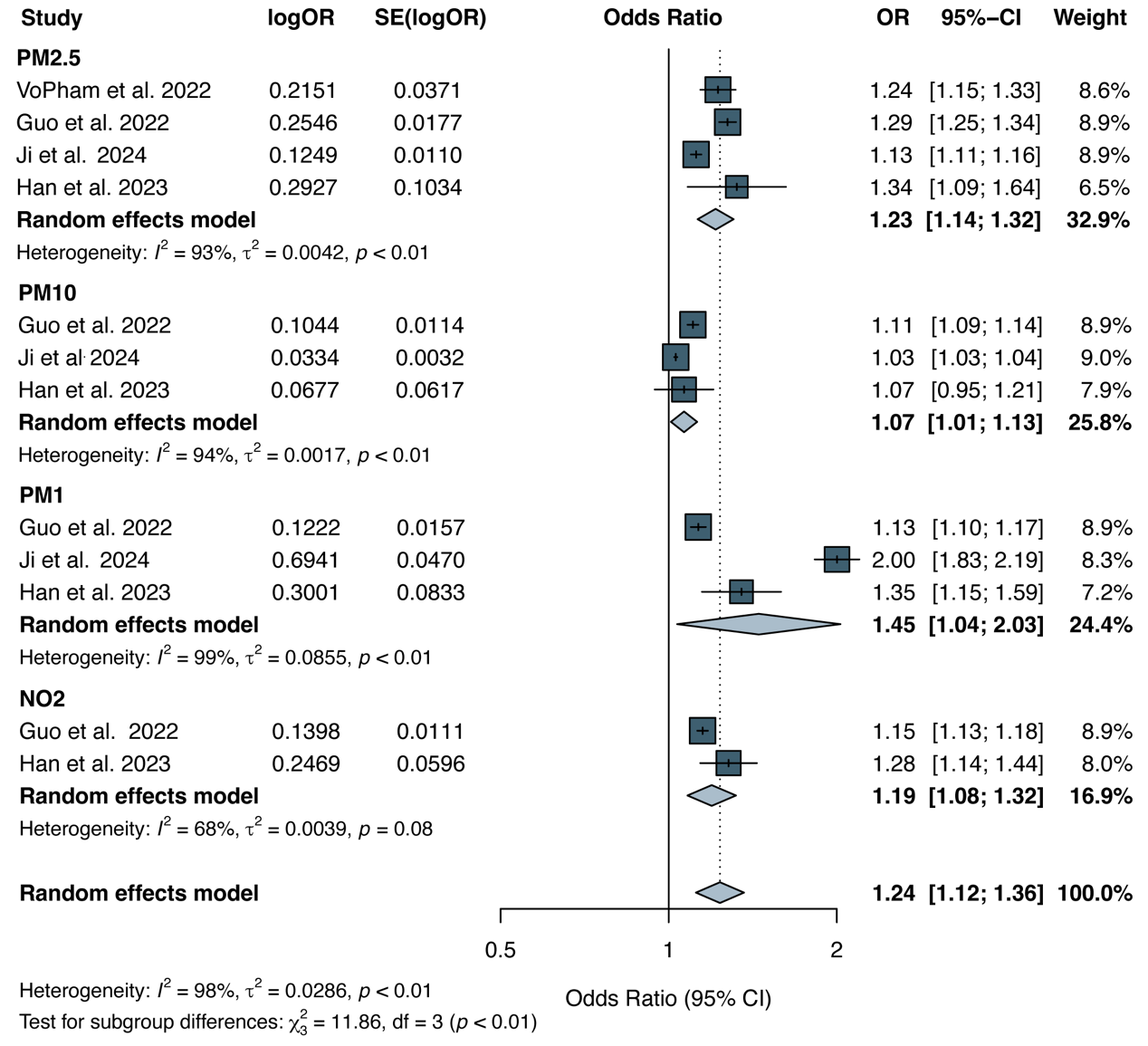
**Figure 2. Forest plot for the association between air pollutants exposure and SLD**

Figure 2. This figure shows pooled odds ratios (ORs) and 95% confidence intervals (CIs) for PM2.5, PM10, PM1, and NO₂ in relation to SLD risk, based on random-effects models. All pollutants showed statistically significant positive associations: PM2.5 (OR = 1.23), PM10 (OR = 1.07), PM1 (OR = 1.45), and NO₂ (OR = 1.19). Substantial heterogeneity was observed for PM2.5 (I² = 93%, τ² = 0.0042), PM10 (I² = 94%, τ² = 0.0017), and PM1 (I² = 99%, τ² = 0.0855), with moderate heterogeneity for NO₂ (I² = 68%, τ² = 0.0039). The overall pooled effect was significant (OR = 1.24, 95% CI: 1.12–1.36), with high heterogeneity across pollutants (I² = 98%, τ² = 0.0286, p < 0.01).

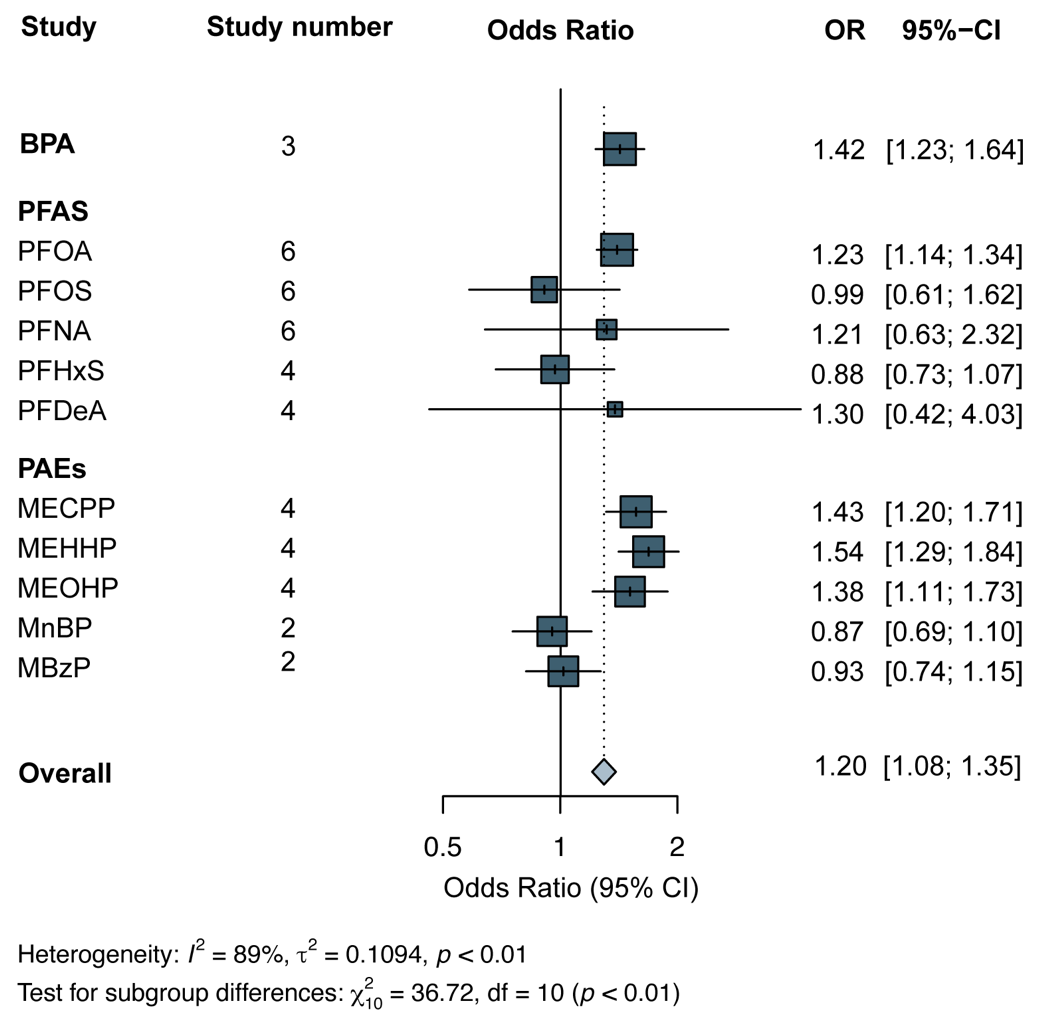
**Figure 3. Forest plot ORs(95%CI) for the association between EDCs exposure and SLD**

Figure 3. This forest plot summarizes the associations between EDCs exposure with the risk of steatotic liver disease (SLD). Pooled odds ratios (ORs) and 95% confidence intervals (CIs) were calculated using a random-effects model. Each square represents the point estimate of the OR for a given chemical, with the size proportional to the study weight; horizontal lines represent the 95% CI. The diamond indicates the overall pooled effect size across all included studies. Significant positive associations with SLD risk were observed for BPA, PFOA, MECPP, MEHHP, and MEOHP. Overall, exposure to these EDCs was associated with increased odds of SLD (OR = 1.20; 95% CI: 1.08–1.35). Substantial between-study heterogeneity was present (I² = 89%, τ² = 0.1094, *p* < 0.01).

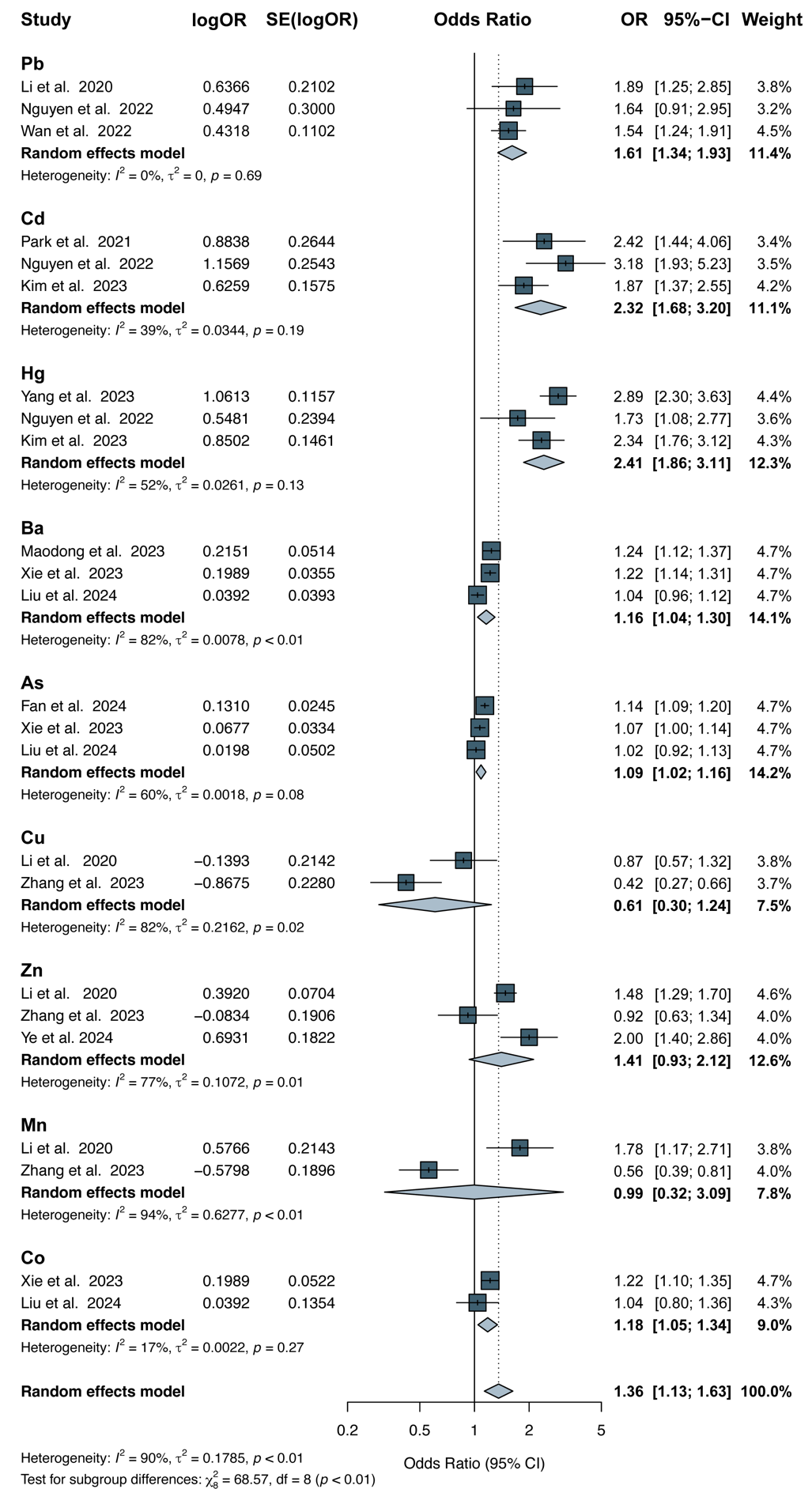
**Figure 4. Forest plot ORs(95%CI) for the association between heavy metals exposure and SLD**

Figure 4. This forest plot summarizes pooled odds ratios (ORs) and 95% confidence intervals (CIs) for nine heavy metals using random-effects models. Significant associations with increased SLD risk were observed for Pb (1.61), Cd (2.32), Hg (2.41), Ba (1.16), As (1.09), and Co (1.18). No significant associations were found for Cu(OR = 0.61, 95% CI: 0.30–1.24), Zn(OR = 1.41, 95% CI: 0.93–2.12), and Mn(OR = 0.99, 95% CI: 0.32–3.09).Substantial heterogeneity was detected for several metals, especially Mn (I² = 94%, τ² = 0.6277), Ba (I² = 82%, τ² = 0.0078), Cu (I² = 82%, τ² = 0.2162), and Zn (I² = 77%, τ² = 0.1072). The overall pooled estimate showed a significant association (OR = 1.36, 95% CI: 1.13–1.63), with high heterogeneity across metals (I² = 90%, τ² = 0.1785, p < 0.01).
